# Integrated Multi-Optosis Model for Pan-Cancer Candidate Biomarker and Therapy Target Discovery

**DOI:** 10.1101/2025.03.06.25323523

**Authors:** Emanuell Rodrigues de Souza, Higor Almeida Cordeiro Nogueira, Ronaldo da Silva Francisco, Ana Beatriz Garcia, Enrique Medina-Acosta

## Abstract

Regulated cell death (RCD) is essential for maintaining tissue homeostasis and controlling cellular stress responses, which are crucial in both cancer suppression, progression, and treatment response across multiple tumor types. RCD processes function in interconnected and overlapping networks. Although few individual RCD forms have been extensively studied, there is a lack of comprehensive approaches integrating multiple RCD forms, limiting the potential for holistic biomarker discovery. Our study addresses this gap by integrating 25 RCD forms into a multi-optosis model that correlates multi-omic and phenotypic data across 33 cancer types using a multilayer approach, enabling the discovery of candidate biomarkers with genome-wide significance. The model was developed using multi-omic data, including tumor and non-tumor tissue samples, from The Cancer Genome Atlas (TCGA) and the Genotype-Tissue Expression (GTEx) databases, accessed via UCSCXena and UCSCXenaShiny. 9,185 tumor samples from TCGA and 7,429 non-tumor tissue samples from GTEx were analyzed. We queried 5,913 genes associated with 25 RCD forms encompassing 62,090 transcript isoforms, 882 mature miRNAs, and 239 cancer-associated proteins and protein modifications. Seven omic features were examined: mutations, copy number variations (CNV), CpG methylation, protein array, mRNA, miRNA, and transcript isoform expression, all correlated with seven clinical phenotypic outcomes: tumor mutation burden (TMB), microsatellite instability (MSI), tumor stemness (TSM), hazard ratio, prognostic survival, tumor microenvironment (TME), and tumor immune cell infiltration (TIL). Over 27 million pairwise correlations were performed between these phenotypic features and multi-omic data. Our findings reveal 44,641 multi-omic signatures, comprising both unique signatures specific to individual RCD forms and overlapping signatures across multiple RCD types, highlighting significant correlations between the omic features and phenotypic outcomes. Each signature received a structured alphanumeric identifier, encoding its biological context, including multi-omic and phenotypic correlations and cancer specificity, providing a systematic approach to categorizing and tracing these associations. Apoptosis-related genes were prevalent across most multi-omic signatures, reaffirming the pivotal role of apoptosis partners among diverse RCD pathways in cancer. Our analysis revealed a prevalent occurrence of isoform-specific signatures, where transcript isoforms originating from the same gene exhibited distinct phenotypic correlations. This finding highlights the intricate roles of alternative splicing and promoter usage in regulating cancer. For instance, in multi-transcript genes like *MAPK10*, specific isoforms were associated with unique phenotypic outcomes, emphasizing the importance of isoform-level resolution for understanding cancer progression and therapeutic response. Notably, rare exceptions were observed in multi-transcript genes such as *COL1A1* and *UMOD*, where all isoforms consistently correlated with stemness indices in a cancer-type-specific manner, showing a coordinated regulatory function at the gene level rather than the isoform level. This finding underscores the potential for specific genes to exert unified effects on tumor biology, which may be crucial for maintaining stemness profiles across cancer types. Significantly, 879 of the identified RCD multi-omic signatures include chimeric antigen targets currently under clinical evaluation, emphasizing the translational potential of these multi-omic signatures in immunotherapy and precision oncology. This integrative framework, accessible through the *CancerRCDShiny* tool (https://cancerrcdshiny.shinyapps.io/cancerrcdshiny/), provides a powerful resource for advancing candidate biomarker discovery and identifying actionable therapeutic targets across cancer types with specific applications in immunotherapy.

## 1. Introduction

Regulated cell death (RCD) represents a highly controlled cellular process crucial for development, tissue homeostasis, and cellular stress responses [1]. This process removes damaged, unnecessary, or potentially harmful cells, supporting organismal function and survival. RCD is essential in cancer research, playing dual roles in tumor suppression, progression and treatment resistance [2, 3].

RCD involves a complex network of signals and mechanisms from various cell death processes rather than functioning through a single, isolated pathway [4, 5]. The cell death processes are categorized into types, referred to as RCD forms, each playing distinct yet sometimes overlapping roles [6]. The RCD forms include apoptosis [7], necroptosis [8], pyroptosis [9], ferroptosis [10], autophagy [11], cuproptosis [12], mitotic catastrophe [13], parthanatos [14], immunogenic cell death [15], autosis [16], NETosis [17], disulfidptosis [18], alkaliptosis [19], lysosome-dependent cell death [20], entosis [21], anoikis [22], oxeiptosis [23], paraptosis [24], cellular senescence [25], mitoptosis [26], erebosis [27], efferocytosis [28], mitochondrial permeability transition [29], methuosis [30], an d necrosis [31]. A summary of the operational definitions of the RCD forms is provided in Figure 1, Dataset S1A.

**Figure 1.**
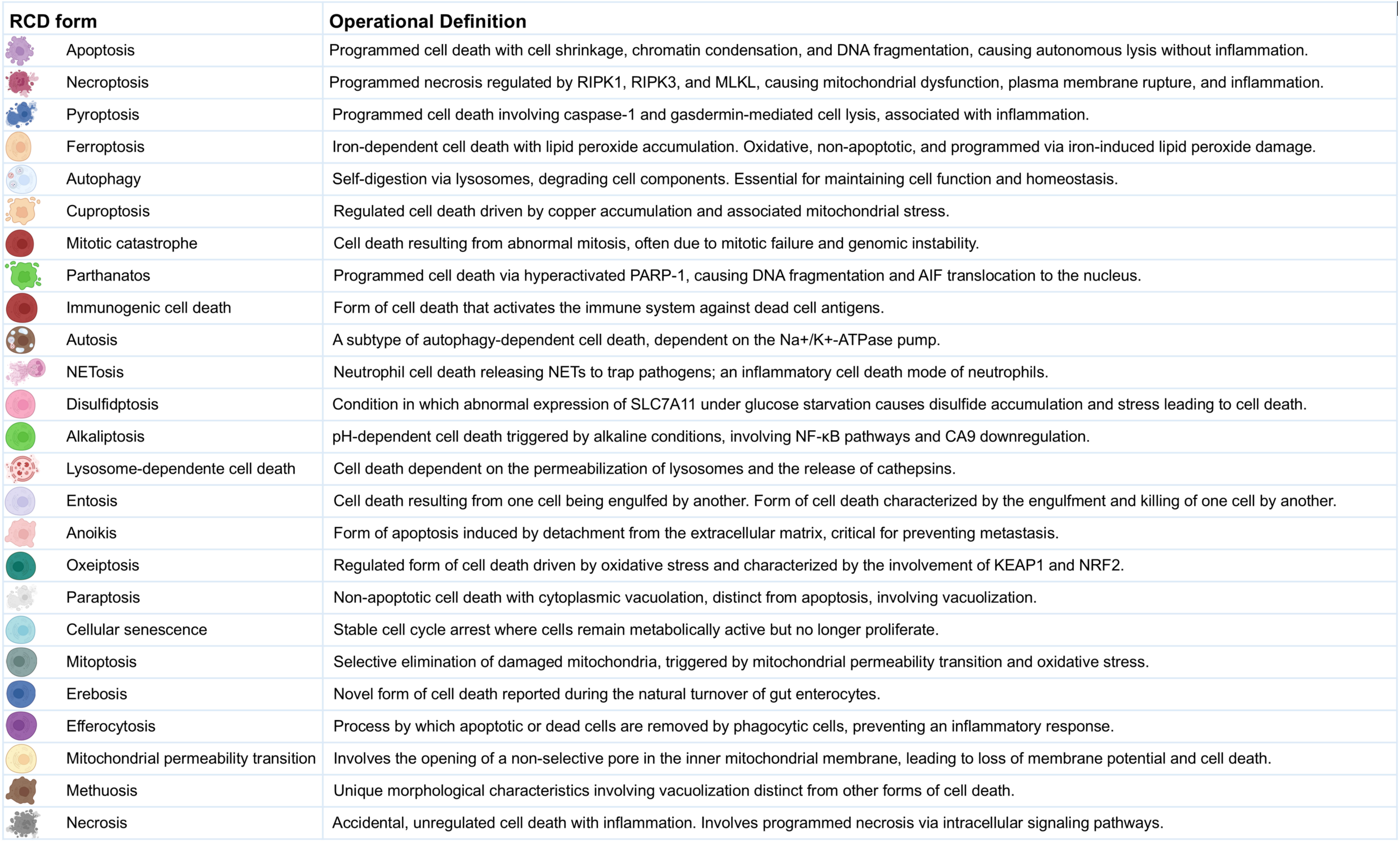
Operational Definitions of Regulated Cell Death Forms. This figure provides detailed operational definitions for the 25 regulated cell death (RCD) forms in the multi-optosis model. Each cell death process is characterized by specific biochemical and morphological features based on the Nomenclature of Cell Death 2018 [4], with additional definitions derived from original research and reviews in 6,603 PDFs (corpus A).

Most studies on RCD in cancer are confined to a death form [32–35]. Multi-optosis, a growing concept describing the crosstalk between different RCD pathways, highlights the complexity of RCD in cancer. This interconnectedness can be exploited for therapeutic strategies that simultaneously induce multiple forms of cell death. Integrating various forms of RCD into explorative strategies to discover biomarkers has ranged from 3-optosis to 15-optosis models in a restricted number of cancer types [36–40].

PANoptosis, a 3-optosis model, describes a unique inflammatory RCD pathway, a coordinated and often simultaneous confluence of features from pyroptosis, apoptosis, and necroptosis [37]. It is thought to play a role in various physiological processes and diseases, including cancer [41–45]. Research on the prognostic value of PANoptosis-related gene signatures in cancer is ballooning. In 2024 alone, the 3-optosis model has been assessed in a variety of cancers, including lung adenocarcinoma [46], breast cancer [47], pancreatic adenocarcinoma [48], hepatocellular carcinoma [44], colon adenocarcinoma [49], gastric cancer [49], head and neck squamous cell carcinoma [50], glioma [51], acute myeloid leukemia [52], thyroid cancer [53], and cutaneous melanoma [54]. Some models are mixed by including aging-associated and extrusion death-associated genes [36].

A 12-optosis model, including apoptosis, necroptosis, pyroptosis, ferroptosis, cuproptosis, entosis, NETosis, parthanatos, lysosome-dependent cell death, autophagy-dependent cell death, alkaliptosis, and oxeiptosis, was evaluated post-surgery in triple-negative breast cancer patients [38]. A 13-optosis model, including disulfidptosis, was assessed for lung carcinoma [39]. Recently, a 15-optosis model was assessed in postoperative bladder cancer patients [40]. This model includes pyroptosis, ferroptosis, necroptosis, autophagy, immunologic cell death, entosis, cuproptosis, parthanatos, lysosome-dependent cell death, intrinsic and extrinsic apoptosis, necrosis, anoikis, apoptosis-like morphology and necrosis-like morphology. The study identified a 13 gene-based cell death signature (*SFRP1*, *CDO1*, *HGF*, *SETD7*, *IRAK3*, *STEAP4*, *CD22*, *C4A*, *VIM*, *TUBB6*, *MFN2*, *FOXO3*, and *YAP1*).

Notably, the 13 genes contribute uniquely to the signature, each with distinct biological functions and associations with immune, tumor microenvironment, and clinical features, rather than sharing correlations across all phenotypic or genomic aspects to provide an overall prognostic score related to cell death in bladder cancer.

The discovery of molecular markers associated with RCD forms can serve as prognostic or predictive biomarkers, guiding treatment decisions and monitoring therapeutic responses [55]. Targeting specific RCD forms can improve the effectiveness of current therapies. For example, in patients with chronic lymphocytic leukemia and acute myeloid leukemia who have relapsed or refractory disease, BH3 mimetics such as Venetoclax (ABT-199), designed to mimic the activity of BH3-only proteins, can sensitize cancer cells to apoptosis by inhibiting anti-apoptotic BCL-2 family proteins [56–58].

Despite the diverse RCD forms, cancer cells often evade these processes through various mechanisms, including those involving cancer stem cells, which are the foundation of the disease [59]. This evasion leads to unchecked proliferation and tumor development [60]. Many standard cancer treatments, including chemotherapy and radiation, aim to induce RCD in cancer cells. However, resistance to these treatments frequently arises from defects in RCD pathways. Mutations in genes regulating apoptosis, such as *TP53* and *BCL2*, are prevalent in various cancers and contribute to resistance to cell death and increased malignancy [61, 62]. Mutations in genes critical for the execution of apoptosis, such as *CASP3* and *CASP9*, have been associated with various cancers, leading to reduced efficacy in chemotherapy and radiation treatments [63]. Mutations can inactivate apoptotic pathways or alter the expression of regulatory proteins like BCL-2 family members, contributing to multidrug resistance in cancer cells [64].

Research on identifying potential markers and therapeutic targets based on RCD forms in cancer often faces shortcomings. Most studies are limited to a single form of RCD [32–35], a specific type of cancer [38, 47, 65, 66], or one kind of association (e.g., mRNA expression versus T cell infiltrates and overall survival) [45, 46, 67]. Studies often overlook the biological significance of whether correlations are positive or negative, the perturbances in gene expression compared to non-tumor samples, or rank the importance of gene signatures based on non-adjusted p-values rather than on a genome-wide scale [68–70]. Many reported gene expression signatures exhibit low correlation scores and limited clinical utility, raising questions about their effectiveness and reliability [32, 71, 72].

Unlike studies that assume uniform behavior of RCD-related genes across cancers, our approach acknowledges that each cancer type has its unique molecular and biological context. Thus, a gene that induces cell death in one cancer might help another cancer evade treatment. An example is *TP53*, commonly known to induce apoptosis in many cancer types. Still, it has been found to promote survival in some contexts depending on the cellular environment and specific mutations present [61]. We thus recognize the non-uniformity in the involvement and roles of RCD-related genes across different RCD forms and cancer types. This non-uniformity means that the activities and effects of these genes can vary widely between different RCD forms and cancer types. By analyzing each gene and cancer type individually, we can understand these differences and identify multi-omic signatures that accurately capture the specific ways RCD-related genes contribute to each cancer. We believe this approach will lead to more precise biomarkers and better-targeted therapies.

Building upon the concept of multi-optosis—describing the intricate crosstalk between distinct RCD pathways—our model integrates 25 forms of RCD into a comprehensive framework (Figure 1; Dataset S1A) to enhance the identification of candidate biomarkers and potential therapeutic targets with genome-wide significance across multiple cancer types. The model provides a holistic view of RCD by analyzing multi-omic and phenotypic features as interconnected entities to understand their combined impact on cancer rather than studying each form independently. The identified signatures integrate clinically meaningful associations between multi-omic and phenotypic variables across 33 cancer types from The Cancer Genome Atlas (TCGA) Pan-Cancer analysis project [73], accessible through the UCSCXena portal [74] and UCSCXena Shiny portal [75, 76]. To facilitate data exploration and analysis, we developed two user-friendly tools: the *CancerRCDShiny* web browser (https://cancerrcdshiny.shinyapps.io/cancerrcdshiny/) and the Cancer Regulated Cell Death Data Analyst (https://chatgpt.com/g/g-8etzMPrtt-cancer-programmed-cell-death-data-analyst). These tools enable efficient extraction, analysis, and visualization of RCD data in cancer and related signatures, supporting a more effective interpretation of relevant data and enhancing the utility and impact of our findings.

## 2. Material and Methods

### 2.1. Multi-optosis Model Specificities

The multi-optosis model integrates 25 forms of RCD (Figure 1, Dataset S1A). Operational definitions of twenty forms of RCD followed the recommendations of the Nomenclature Committee on Cell Death 2018 [4]; RCD operational definitions not provided in the review by Galluzzi et al., 2018 were based on original research and reviews included 6,603 manually curated, free-access full-text PDF documents (Corpus A, Supplementary Material File S1). We extracted, processed, and analyzed data on various forms of RCD and their associations with cancer by using the PDF Ai Drive Tool (https://myaidrive.com/), which uses advanced Large Language Models (LLMs) and natural language processing (NLP) techniques to extract and contextually analyze data accurately. PDF AI Drive uses six AI models to summarize and extract structured information from PDF documents. The models are Claude 3 Haiku, Claude 3.5 Sonnet, Claude 3 Opus, CommandR+, Gemini 1.5 Flash and GPT-4o OpenAI (latest). GPT-4o provided us with the most detailed outputs.

A multi-optosis inventory of 5,913 genes was compiled by querying each RCD form term in the NCBI Gene database using a Boolean approach [77] (Dataset S1B). The information was then programmatically extracted in R using the NCBI ‘*Entrez*’ package. This approach solely reflects terms related to RCD forms and does not imply direct functional or causative involvement.

### 2.2. Correlation Analysis Between Multi-omic and Phenotypic Variables in 33 Cancer Types

We conducted a comprehensive computational analysis correlating multi-omic variables with phenotypic outcomes from the TCGA Pan-Cancer analysis project [73], using primary datasets sourced from the UCSC Xena portal [74], including the TCGA Pan-Cancer Atlas [73]. Secondary datasets were obtained from the UCSC XenaShiny portal [75, 76], including the GTEx dataset for non-tumor tissue comparisons [78].

The multi-omic feature included RNA-Seq transcriptomics (mRNA expression, transcript isoform expression, and miRNA expression), CpG methylation (450K array), CNV (gistic2 thresholded), mutations (SNP and INDEL; MC3 public version), and reverse-phase protein expression array (TCGA RPPA microarray) [79, 80]. The microarray comprises 258 protein and modification probes relative to 210 genes, of which 239 are term-based associated with RCD forms (Dataset S1C). miRNA gene symbols were converted to precursor miRNA identifiers (IDs) using ‘*BioMart*’ [81], and the precursor IDs were converted to mature miRNAs (Dataset S1D) using the ‘*miRBaseConverter*’ R package to analyze miRNA. Gene symbols were converted to transcript IDs (Dataset S1E) using the ‘*BioMart R*’ package.

The phenotypic features included the patient’s indexes for tumor mutation burden (TMB), microsatellite instability (MSI), tumor stemness (TSM), hazard ratio (HRC), prognostic survival metrics (SMC), tumor microenvironment (TME) and tumor immune infiltrates (TIC) indexes. The analysis was performed in R, using functions and customized source code based on the UCSC XenaShiny package [75, 76]. These tools enabled us to programmatically execute multiple iterative analyses between multi-omic and phenotype features spanning 33 cancer types (n=9,185 samples, Dataset S1F). To identify the most significant correlations, we used genome-wide significance (Holm-Bonferroni adjusted p (*padj*-value) < 5×10^-8) as the threshold for the association between the multi-omic and the phenotypic features (TMB, MSI, STM). The genome-wide threshold minimizes false positives because of multiple comparisons and provides a stringent criterion for identifying significant associations across the genome.

For the comparison of mRNA expression between tumor and non-tumor tissues, including primary-tissue-matched samples from the GTEx project (n=7,429 samples, Dataset S1F), we use the Wilcoxon test [78]. This non-parametric test was selected to handle potential deviations from normality in the expression data, ensuring robust comparative analysis. TCGA versus GTEx tissue RNA-Seq expression profiles were classified as unchanged expression, underexpressed, overexpressed or with no data. Unchanged expression includes genes with a *padj*-value ≥ 0.05. Genes are classified as overexpressed or underexpressed if they have a *padj*-value < 0.05. Overexpressed genes show higher median expression in tumor tissue, while underexpressed genes show lower median expression in tumor tissue, both compared to non-tumor tissue.

We performed hazard ratio analysis using the Cox proportional hazards regression model to assess the prognostic significance of the association between omic variables and patient survival outcomes, providing hazard ratios that refer to the relative risk of events occurring at any given point in time.

Multi-omic feature with consistent correlations, showing the same direction in tumor versus non-tumor expression, and Cox hazard ratio were used to create signatures. These signatures were then evaluated individually by summing the values of the constituent features (e.g., member 1 + member 2 + … + member n). The prognostic significance of the constructed signatures was evaluated using Cox proportional hazards analysis for four survival metrics: Disease-Specific Survival (DSS), Disease-Free Interval (DFI), Progression-Free Interval (PFI), and Overall Survival (OS). Kaplan-Meier survival curves were generated for each metric, and log-rank tests applied to compare survival distributions across patient groups, determining the statistical significance of observed differences. Together, these survival analyses offer a comprehensive view of patient outcomes and provide valuable insights into the effectiveness of cancer treatments [82]. The survival metrics are defined: DSS specifically measures survival without death attributed to the cancer being studied. It provides a more focused measure of treatment effectiveness on the targeted disease. DFI assesses the period after treatment during which the patient remains free from any signs or symptoms of cancer. It is useful in evaluating the efficacy of therapies. PFI measures the duration in which the cancer does not progress or get worse. OS is a critical endpoint in cancer clinical trials, measuring the time from randomization or diagnosis to death from any cause. It is the most definitive endpoint, reflecting the ultimate impact of the treatment on patient survival [83].

### 2.3. Classification of Signatures According to the Tumor Microenvironment Profile

We used CIBERSORT (Cell-type Identification By Estimating Relative Subsets Of RNA Transcripts) [84] and xCell [85] deconvoluted bulk gene expression data from UCSCXenaShiny [75, 76] to estimate correlations of the multi-omic gene-signature feature and the cellular composition of complex tissues based on 29 predefined immune cell signature subsets, including B cells (naïve, memory, plasma, class-switched memory), T cells (CD8+, CD4+ naïve, CD4+ memory resting, CD4+ memory activated, CD4+ Th1, CD4+ Th2, follicular helper, regulatory Tregs, gamma delta), NK cells (resting and activated), monocytes, macrophages (M0, M1, M2), myeloid dendritic cells (resting and activated), activated mast cells, eosinophils, neutrophils, cancer-associated fibroblasts, common lymphoid progenitor, endothelial cell, granulocyte-monocyte progenitor, and hematopoietic stem cell.

We categorized the signatures as anti-tumoral, pro-tumoral, or dual regarding tumor progression. This classification was based on the Spearman correlation coefficients between mRNA, miRNA, isoform RNA-Seq or protein expression of the signature database and the RNA-Seq expression profiles of the 29 specific cell infiltrate types representative of the tumor microenvironment profile (Dataset S1G). We used the categorizations hot, cold and variable for the involvement of cell infiltrates based on evidence from the literature (Dataset S1G).

In this system, the signs and magnitudes of the correlation coefficients provide insights into different tumor microenvironment scenarios (See Figure S1 for the categorization framework of tumor microenvironments and immune phenotypes across multiple scenarios). A positive correlation with a cell type shows a higher presence of that cell type in the tumor microenvironment for signatures that are overexpressed in a tumor type. Conversely, for underexpressed signatures, a positive correlation with a cell type shows a lower presence of that cell type. For overexpressed signatures exhibiting a negative correlation, the correlation sign also shows a lower presence of that cell type. Similarly, underexpressed signatures with a negative correlation show a higher presence of that cell type. For signatures whose expression profiles are unaltered between tumor and non-tumor tissues, a positive correlation indicates the presence of cell infiltrates, while a negative correlation indicates their absence.

We combined the correlation coefficients for all cell types to classify the signatures according to the tumor microenvironment, considering their signs. Signatures with the highest combined magnitude for anti-tumoral cell types were classified as anti-tumoral. Similarly, signatures with the highest combined correlations for pro-tumoral cell types were classified as pro-tumoral, and signatures with the highest combined correlations for dual microenvironment cell types were classified as dual. Detailed methodology is provided in Supplementary Material File S1 (Methodology 1).

### 2.4. Classification of Signatures According to the Tumor Immune Phenotype

The tumor immune phenotype, classified as hot, cold or variable based on immune cell infiltration, guides therapeutic interventions and identifies patients resistant to immunotherapies [86, 87]. Hot tumors exhibit high levels of cytotoxic T cells (NK and CD8+) and M1 macrophage signatures, while cold tumors show low T cell infiltration, a predominance of M2 macrophages, and immunosuppressive cells. Variable tumors have intermediate characteristics (Dataset S1H). Examples include melanoma and lung cancer as hot and prostate and pancreatic cancers as cold. Immune checkpoint inhibitors are more effective in hot tumors [86, 87]. Strategies to convert cold and variable tumors to hot ones, such as nanomedicines and combination therapies, are under development [88].

We used a classification method analogous to immunohistochemistry as a proxy to quantify tumor lymphocyte infiltration using RNA-Seq indexes [86, 87, 89], enabling categorization as "hot", "cold" or "variable". This allows automated categorization of signatures as "hot", "cold" or "variable" in R, enhancing understanding of tumor immunological characteristics and potential responses to immunotherapies. "Hot" tumors correlate positively with cytotoxic T cells and M1 macrophages, while "cold" tumors show low correlations with these cells but high correlations with M2 macrophages and Tregs. "Variable" tumors exhibit intermediate correlations (Dataset S1H).

For classification, we used Spearman correlation coefficients and p-value significance between RNA-Seq-based expression profiles of signatures and immune cell profiles (T CD8+, NK, M1/M2 macrophages, and Tregs) [86, 87]. In ambiguous cases, we applied a differentiated weighting criterion, prioritizing CD8+ T cells and NK cells because of their importance in classifying "hot" tumors and predicting immunotherapy responses. Detailed methodology is provided in Supplementary Material File S1 (Methodology 2).

### 2.5. Multi-optosis and Multi-omic Signature Nomenclature

The signature nomenclature system provides a structured alphanumeric identifier that categorizes signatures derived from multi-omic Pan-Cancer analysis. This system links multi-omic feature of target genes with phenotypic characteristics across 33 cancer types, ensuring high precision and clarity in data organization and retrieval. The signature identifier follows an eleven-component structure: **CTAB-GSI.GFC.PFC.SCS.TNC.HRC.SMC.TMC.TIC.RCD** (e.g., KIRP-107.3.2.N.1.44.44.1.1.2) (Figure 2).

**Figure 2.**
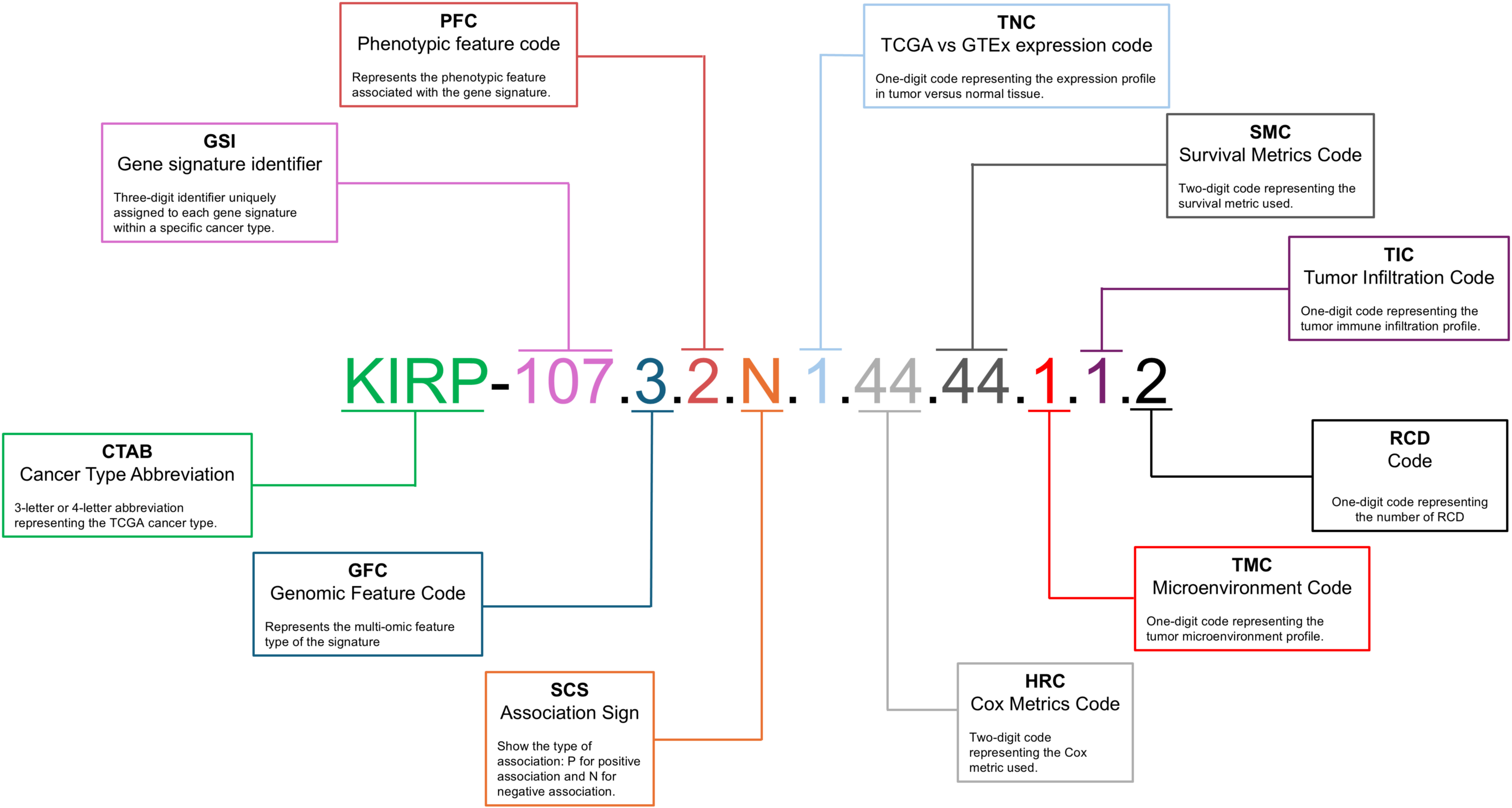
Multi-omic Signature Nomenclature and Coding System. This figure details the nomenclature and coding system used for multi-omic signatures in the multi-optosis model. Each signature is uniquely identified by a series of codes that represent different attributes: the cancer type abbreviation is a three- or four-letter abbreviation denoting the TCGA cancer type (e.g., KIRP for kidney renal papillary cell carcinoma); the phenotypic feature code is a one-digit code showing the specific phenotypic feature associated with the signature; the genomic feature code is a one-digit code representing the multi-omic feature; the signature identifier is a unique three-digit number assigned to each signature within a specific cancer type; the correlation sign shows the type of association, with ’P’ for positive and ’N’ for negative; the TCGA vs GTEx expression code is a one-digit code showing the gene expression profile in tumor tissue compared to non-tumor tissue; the Cox metrics code is a two-digit code representing the Cox proportional hazards metric used in the analysis; the survival metrics code is a two-digit code showing the specific survival metric applied; and the tumor infiltration code is a one-digit code representing the tumor immune infiltration profile. An example of a multi-omic signature identifier, such as KIRP-107.3.2.N.1.44.44.1.1.2, illustrates how these codes combine to form a comprehensive identifier for each signature. This standardized coding system enables precise classification and analysis of signatures in cancer research.

Each component is defined as:

**CTAB** refers to a 3- or 4-letter abbreviation representing the cancer type from the TCGA database (e.g., KIRP for kidney renal papillary cell carcinoma; see Dataset S1I for cancer type abbreviations).

**GSI** is a 1- to 4-digit identifier (e.g., 107) unique to each signature within a cancer type.

**GFC** represents the multi-omic feature type of the signature: 1 for Protein expression, 2 for Mutations, 3 for CNV, 4 for miRNA expression, 5 for Transcript expression, 6 for mRNA expression, and 7 for CpG Methylation.

**PFC** denotes the phenotypic feature linked to the signature: 1 for TMB, 2 for MSI, and 3 for TSM.

**SCS** shows the Spearman Correlation Sign: P for positive and N for negative correlations.

**TNC** represents tumor vs. non-tumor tissue expression: 0 for no data, 1 for unchanged expression, 2 for underexpressed, and 3 for overexpressed.

**HRC** stands for Hazard Ratio Code, represented as the alphanumeric array 1N2N3N4N. This shorthand notation encodes the significance levels of multiple survival metrics. The digits 1 to 4 correspond to the survival metrics: DSS, DFI, PFI, and OS, respectively. The letter N denotes the hazard effect, classified as A (no effect), B (risky), or C (protective).

**SMC** is the Kaplan-Meier survival distribution across patient groups. It also follows the array 1N2N3N4N, where the digits 1 to 4 correspond to survival metrics: DSS, DFI, PFI, and OS, respectively. However, the categorization of the letters A, B, C, and D across multi-omic feature reflects distinct classifications based on specific criteria. The letter A is used universally for all omic layers (Protein, Mutation, CNV, miRNA, Transcript, mRNA, and Methylation) when the category is "NS" (Not Significant).

The letter B varies according to the omic layer. For Protein, miRNA, Transcript, mRNA, and Methylation, it corresponds to the category "High". For the Mutation feature, B represents "MT" (Mutant), while for the CNV feature, B refers to "Deleted." Similarly, the letter C also differs by omic layer. For Protein, miRNA, Transcript, mRNA, and Methylation, C corresponds to the category "Low." For the Mutation feature, it represents "WT" (Wild Type), and for CNV, it reflects the "Duplicated" status.

The letter D is used explicitly for the CNV feature and represents the category "Deleted/Duplicated," where both deletion and duplication events are involved.

There are 128 combinations of the 1N2N3N4N array for hazard values and survival metrics. Each array combination is reassigned to a specific numerical identifier ranging from 0 to 127 (Dataset S1J). For instance, 1A2A3A4A (no effect for DSS, DFI, PFI, and OS) is reclassified to the identifier 0. In contrast, 1A2A3A4B (no effect for DSS, DFI, and PFI, yet “risky” for OS) is reclassified as to the identifier 1, accordingly.

**TMC** refers to the Tumor Microenvironment Code: 1 for anti-tumoral, 2 for dual, 3 for pro-tumoral, and 4 for no significant data.

**TIC** is the Tumor Lymphocyte Infiltrate Code, which defines immune cell infiltration: 1 for "hot", 2 for "variable", 3 for "cold", and 4 for no significant data.

**RCD** is a 1- to 2-digit code representing the number of RCD forms linked to the signature.

### 2.6. Signature Rank Method

We developed the *Cancer Multi-optosis Multi-omic Signature Rank Calculator in R* to evaluate how effectively a signature provides valuable, actionable insights to improve patient care or inform clinical decisions—its clinical meaningfulness potential—within our Pan-Cancer multi-optosis and multi-omic model. This system ranks candidate biomarker signatures by integrating multi-omic and phenotypic identifiers. Each component within a signature is assigned an integer rank based on its importance in predicting patient outcomes, such as survival prognosis [90] and immunotherapy potential. The immunotherapy potential is assessed using TME and TIC identifiers, applying the concepts of immune "hotness" and "coldness," which reflect the level of immune infiltration in tumors [86, 87]. A rank is assigned to each signature component through a mapping function in R, which attributes integer values to multi-omic and phenotypic identifiers. The final rank for each signature is obtained by summing the ranks of its individual components. Detailed criteria for assigning ranking values are provided in Supplementary Material File S1 (Methodology 3).

### 2.7. Drug-Gene Interaction Analysis

To identify potential therapeutic targets, we conducted a comprehensive cross-referencing analysis of gene components from the top-ranked multi-modular and clinically meaningful signatures. The gene members of these signatures were queried against the Drug–Gene Interaction Database (DGIdb 5) [91], which integrates drug-gene interaction and druggability data from multiple sources, facilitating the exploration of potential pharmacological interventions.

To construct the drug-gene interaction network, we retrieved curated interaction data from DGIdb 5.0, excluding undefined or unknown interaction types to ensure meaningful associations. The dataset was processed in R using the ‘*tidyvers*’ suite, which included data cleaning, removal of redundant entries, and standardization of gene and drug names. A bipartite network was generated, where genes (from top-ranked multi-omic RCD signatures) formed one node type, and drugs (categorized by interaction type) formed the other node type. The edges in the network represent drug-gene interaction relationships, as defined by DGIdb. Network visualization was performed using ‘*igraph*’ and *‘ggraph’* for static rendering, with the Fruchterman-Reingold force-directed layout applied to optimize node distribution and improve clarity.

### 2.8. Validation Using the Independent PRECOG Cancer Database

To validate the prognostic value of the selected mRNA-specific signatures associated with risk, protection, and poor prognosis, we used the PRECOG (PREdiction of Clinical Outcomes from Genomic Profiles) database [92]. PRECOG is a curated resource that provides a standardized meta-analysis framework to generate prognostic meta-Z scores, which quantify the strength and direction of the association between gene expression and overall survival (OS) across multiple cancer types. The database integrates transcriptomic data from publicly available datasets, encompassing 28 cancer types independent of TCGA but equivalent to 24 TCGA cancer types (Dataset S1I). Meta-Z scores were extracted from the PRECOG repository (https://precog.stanford.edu) for each gene within the 126 signatures selected for their association with risk or protection in all survival metrics and with anti-tumoral, pro-tumoral or dual microenvironment cell profiles, and hot, cold or variable immune infiltrates (Dataset S1K). To validate significant poorer or better prognosis associations, the validation process relied on stringent statistical thresholds (|Meta-Z| > 3.09 or < -3.09, p < 0.001). This validation set of signatures represents only 11 TCGA cancer type (ACC, BLCA, BRCA, CESC, HNSC, KIRP, LGG, LUAD, LUSC, PRAD, STAD). For single-gene signatures, the corresponding meta-Z score was retrieved for each cancer type. For multi-gene signatures, each gene was queried individually, and the median meta-Z score across all genes was computed to derive the final signature-level score. To identify cancer-specific prognostic associations, we compared the direction of association between PRECOG meta-Z scores and our gene signatures, refining the selection of relevant cancer-specific signatures. Positive meta-Z scores show a poor prognosis, while negative scores suggest a favorable prognosis.

### 2.9. PDF-Ai-Assisted Evidence of Involvement of Signature Members in the Multi-Optosis Model

A drawback of most multi-omic studies aimed at discovering biomarkers in cancer is the lack of cross-referencing with databases. Flat lists of genes with limited features are often reported [5, 69, 93, 94], restricting understanding of their potential applications. We implemented a PDF generative artificial intelligence-based (PDF-Ai) strategy to provide evidence-based support for the involvement of the identified signature members. The strategy cross-references signature members with structured information from the scientific literature. This approach uses LLMs within a ChatGPT-based PDF-AI analysis tool to extract relevant data directly from the PDF corpus A, ensuring robustness and reproducibility. The method involves several key supervised, executable sequential tasks that focus on identifying mentions of gene members of the signatures, associated RCD forms, and cancer types (see Supplementary Material File S1 – Methodology 4). The last step involves validating the cross-referenced data through manual review and automated checks to ensure data integrity and reliability. Any discrepancies were resolved manually to maintain the robustness of the dataset. By implementing this PDF-AI strategy, the applicability of findings is enhanced through a user-friendly data analysis tool. The structured tabular output information was compiled to create the *Cancer Regulated Cell Death Data Analyst* (https://chatgpt.com/g/g-8etzMPrtt-cancer-programmed-cell-death-data-analyst), a user-friendly, publicly accessible GTP-based chat software engineer for extracting, analyzing, and visualizing RCD data in cancer and signature members. This tool allows Chat-GPT registered users to access and interpret the relevant data efficiently, enhancing the applicability and impact of our findings.

A detailed inventory of established immunotherapy targets and their presence within the multi-omic RCD signature repertoire is presented in Dataset S1L. The relevance and representation of these targets were assessed by cross-referencing with a curated corpus (Corpus B) of 642 manually selected PDF articles using PDF AI extraction (Supplementary Material File S1).

The PDF corpora were compiled using the NCBI *pubmed* R package and the RIS identifiers were used to download free-text using EndNote^TM^ citation software (https://endnote.com/).

## 3. Results

The construction and analysis of the multi-optosis model, depicted in the workflow (Figure 3), provide a comprehensive framework that integrates 25 distinct forms of RCD (Figure 1). This model is founded on a core gene set of 5,913 RCD term-based gene symbols (Dataset S1B). The broader RCD gene inventory includes 62,090 transcripts, spanning both primary and alternative isoforms, 882 mature miRNAs (representing both 5p and 3p strands), and 239 proteins known to be associated with cancer, including post-translational modifications. These elements form the backbone of our investigation, offering extensive coverage of RCD-related genes across cancer types.

**Figure 3.**
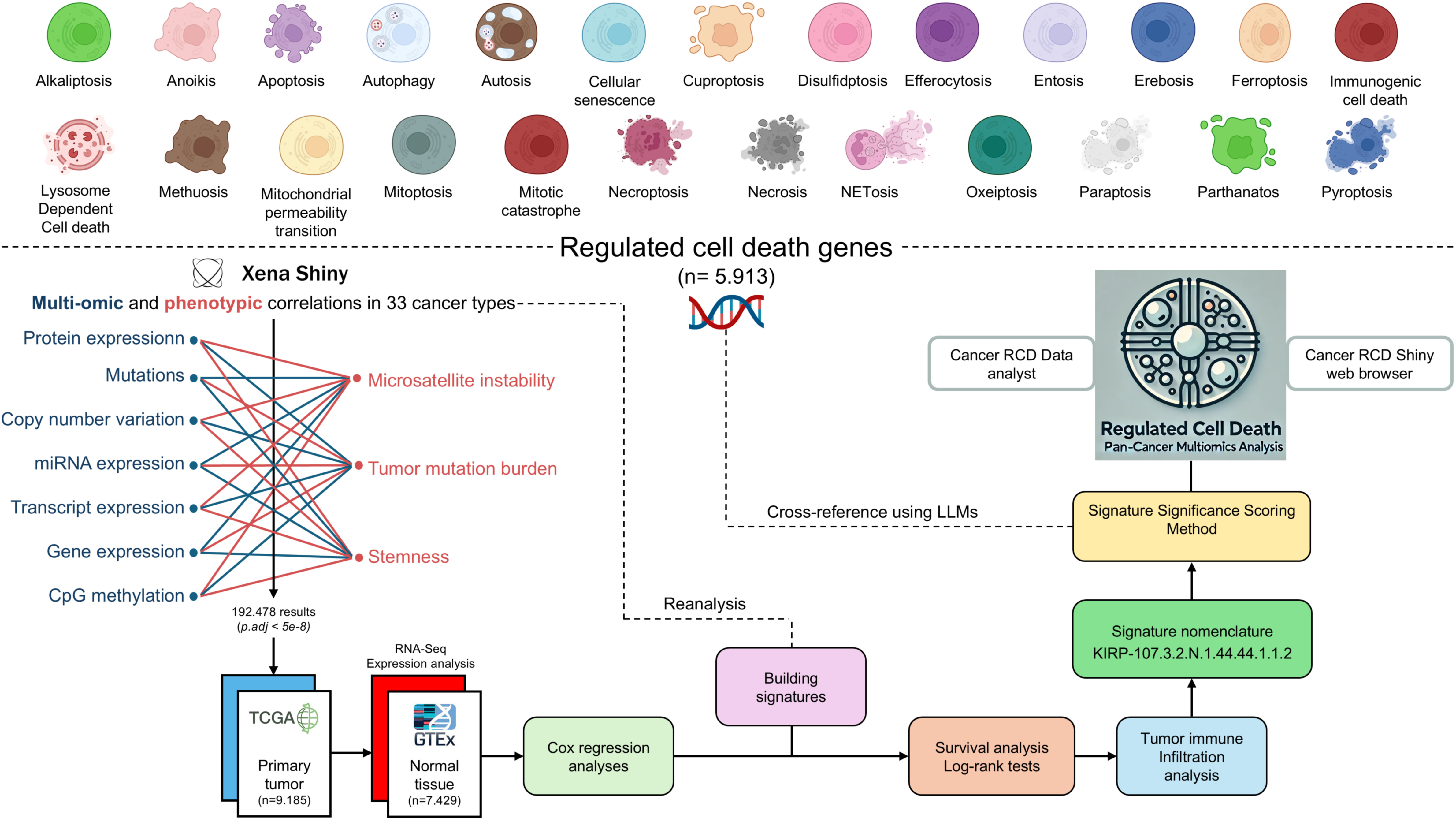
Workflow of the Multi-Optosis Model Analysis. This workflow illustrates the detailed process for constructing and analyzing a multi-optosis model focusing on 25 regulated cell death (RCD) mechanisms. The process begins with identifying 5,913 RCD-related genes using the NCBI ‘*Entrez’* function in R. Multi-omic and phenotypic data from TCGA Pan-Cancer are then integrated using the ‘Get Xenà R script. Expression and correlation analyses are conducted with a stringent p-value threshold (< 5e-8) using the ‘main’ R script, then consolidating all results into a single data frame. The hazard ratio is assessed for four survival metrics: overall survival (OS), disease-specific survival (DSS), disease-free interval (DFI), and progression-free interval (PFI) using Cox proportional hazards models and log-rank tests. Tumors are classified into “hot”, variable, or “cold” categories based on immune infiltration profiles. Each signature is assigned a unique nomenclature, and significance scoring is applied. The roles and involvement of gene members in various RCD forms in cancer are investigated. Finally, cross-referencing and visualization are enabled through the *CancerRCDShiny* web browser and the LLM-based *Cancer Regulated Cell Death Data Analyst* tool, allowing for interactive exploration and visualization of the findings. This structured approach integrates computational and statistical methods to enhance understanding of regulated cell death mechanisms in cancer.

Approximately 40% (n=2,403) of all genes in the inventory are involved in two or more forms of RCD (Dataset S1B). Genes exclusively associated with apoptosis account for approximately 42% (n=2,511) of the target genes, showing no term-based association with other RCD forms. The RCD forms with the fewest associated genes are alkaliptosis, lysosome-dependent cell death, and methuosis (Dataset S1M).

Notably, 422 genes in the inventory are established Cancer Gene Census Tier 1 driver genes (n=584, 72.3%) in COSMIC (Catalogue Of Somatic Mutations In Cancer) [95] and other databases [96]. These include oncogenes, tumor suppressors, and fusion genes, all linked to at least one RCD form (Dataset S1B). Driver genes such as *TP53*, *AKT1*, *MTOR*, *CD274*, *PTEN*, and *STAT3* are linked to at least eight RCD forms. Among these, *TP53* stands out as the most prominent driver gene, being associated with 12 distinct forms of RCD: anoikis, apoptosis, autophagy, cellular senescence, entosis, ferroptosis, mitochondrial permeability transition, mitotic catastrophe, necroptosis, pyroptosis, necrosis, and autosis. However, several non-driver genes, such as *SIRT3*, *CXCL8*, *NFKB1*, *STING1*, and *TNF*, are noteworthy for their presence across at least eight RCD forms (Dataset S1B).

The multi-optosis model integrates multi-omic and phenotypic reiterative correlations estimated from the TCGA Pan-Cancer secondary database [74–76], using R coding based on functionalities from the UCSCXenaShiny [75, 76]. Correlation analyses were performed between the seven omic features with seven phenotypic and clinical variables in 33 cancer types. For each gene target, survival metrics were assessed using Cox proportional hazards models. Unique, single- and multi-gene signatures were built according to feature commonalities, and their prognosis values of the signatures were evaluated using the log-rank test in four survival metrics. Each signature was then queried for significant correlations with the expression profiles indicative of immune and nonimmune cell infiltrates to determine their association values with the tumor microenvironment. We performed 27,238,756 pair associations between multi-omic, phenotypic, risk, survival and cell immune infiltration features.

The multi-omic and phenotypic features associated with each gene member in the signatures are compiled into an extensive integrative database (Dataset S2) comprising 44,641 multi-omic signatures across 32 cancer types. None of the target genes achieved genome-wide significance with phenotypic variables in Lymphoid Neoplasm Diffuse Large B-cell Lymphoma (DLBC).

The number of elements per signature ranged from 1 to 2,052 (mean = 4.3; median = 1; Q1 = 1; Q3 = 2; P90 = 6, meaning that only 10% of signatures contain over 6 elements; Dataset S2). Importantly, for the multi-member signatures, all the components share the association features, the RCD type(s), and the statistical significance level. The maximum number of member elements per omic feature is: 2,052 (Transcript), 487 (Mutation), 477, (mRNA), 423 (Methylation), 124 (CNV), 58 (miRNA) and 4 (protein) (Figure S2).

The distribution of multi-omic signatures across omic features and cancer types is represented in Figure 4. This accumulated histogram provides insight into the proportional presence of each omic feature within different cancer types, with the absolute accumulated counts depicted per feature.

**Figure 4.**
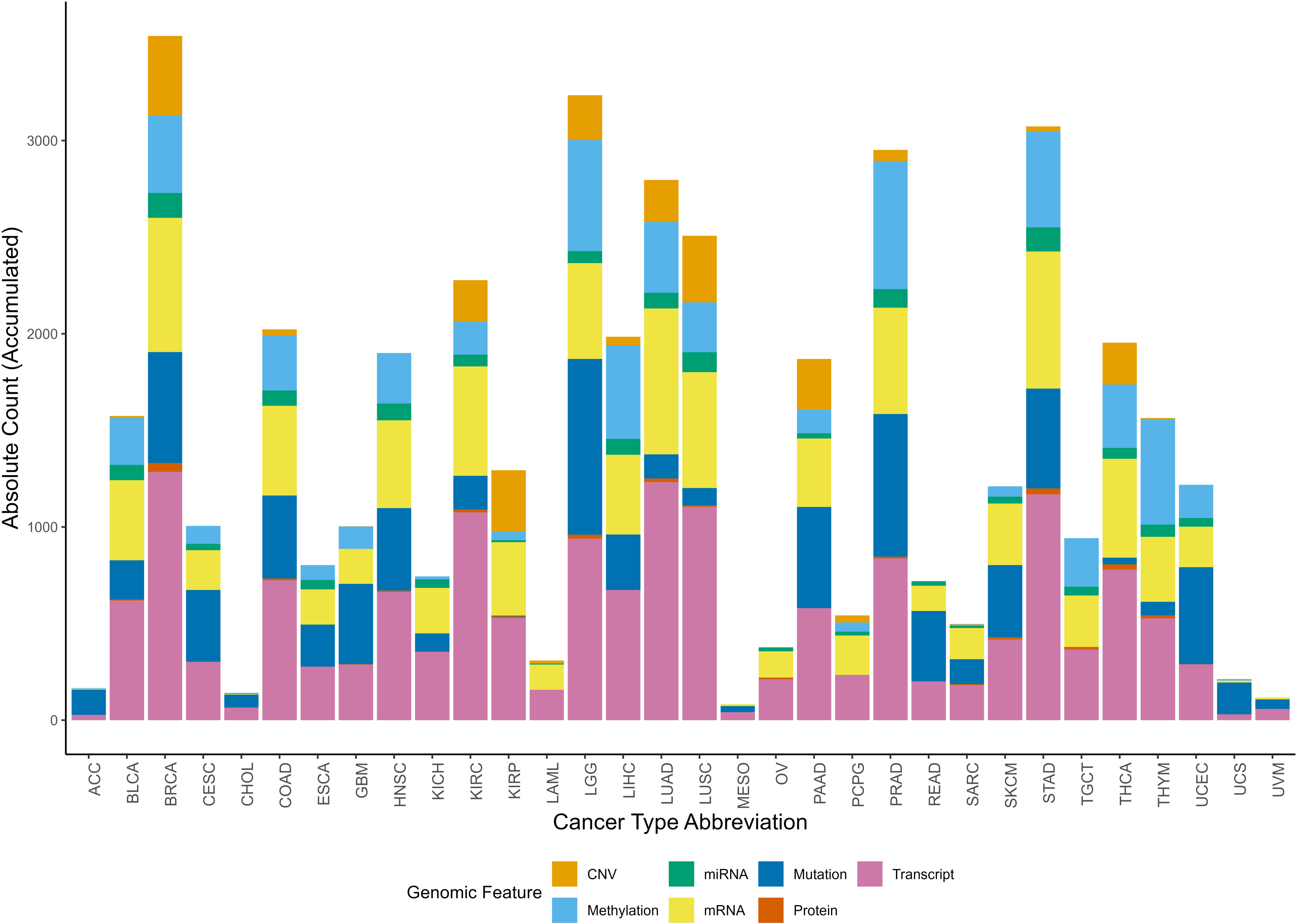
Accumulated histogram illustrating the distribution of multi-omic signatures by multi-omic feature across various cancer types. Each bar represents a unique Cancer Type Abbreviation, with colors depicting the relative proportions of signatures across multi-omic feature. The height of each bar shows the absolute accumulated count of signatures for each multi-omic feature within each cancer type. The Okabe-Ito color-blind friendly palette has been applied to enhance accessibility for all viewers.

The top-ranked cancer types by the number of signatures for each omic feature reveal specific molecular patterns (Figure 4). Breast Cancer (BRCA) has the highest number of signatures associated with CNV, protein expression, transcript, and miRNA, with absolute counts of 413, 45, 1,286, and 129, respectively. Prostate Cancer (PRAD) ranks the highest in methylation-associated signatures, totaling 663, while LGG (Lower Grade Glioma) has the greatest number of mutation-linked signatures, with 910. Lung Adenocarcinoma (LUAD) exhibits a high frequency of mRNA-associated signatures, totaling 755.

Of the 5,913 target genes, 5,777 (97.7%) reached a significant correlation and are therefore included as elements in the signature database. Of the remaining genes, 101 did not achieve significance, and 35 lacked data in the Xena database. Most of the signatures include at least one apoptosis-related gene (34,500; 77.2%). This rate is expected, since 4,812 (81,4%) target genes are associated with apoptosis (Dataset S2).

Among transcript isoform signatures, the ten most frequently occurring genes were *EFEMP2*, *ABI3BP*, *TPM1*, *ELN*, *FN1*, *COL1A1*, *DCN*, *PDLIM7*, *TCF4*, and *COL1A2*, each appearing in 69 to 91 signatures. Collectively, these genes are associated with anoikis, apoptosis, autophagy, cellular senescence, necrosis, and pyroptosis (Dataset S1N).

The identifier KIRP-107.3.2.N.1.44.44.1.1.2 exemplifies the nomenclature system used throughout, as shown in Figure 2. KIRP represents the Cancer Type Abbreviation (CTAB) for Kidney Renal Papillary Cell Carcinoma, and 107 is the Gene Signature Identifier (GSI), showing the 107th signature identified for this cancer type. The Genomic Feature Code (GFC) is 3, corresponding to CNV, while the Phenotypic Feature Code (PFC) is 2, indicating MSI. The Spearman Correlation Sign (SCS) is N, signifying a negative correlation. The TCGA vs. GTEx Expression Code (TNC) is 1, meaning the gene expression is unchanged in tumor tissue compared to non-tumor tissue. The Hazard Ratio Code (HRC) is 44, based on the combination 1B2B3B4B, which shows a risk effect by all survival metrics (DSS, DFI, PFI, and OS). The Survival Metrics Code (SMC) is also 44, derived from the combination 1B2B3B4B, reflecting specific prognostic implications across all four survival outcomes. TMC is 1, indicating a correlation with an anti-tumoral environment immune profile. TIC is 1, showing an association with “hot” profiling of immune cell infiltration. Finally, RCD is 2, signifying that the gene members are associated with two RCD forms, namely apoptosis and necrosis.

The commonalities of the signatures can be explored and analyzed purposefully or guided. Here, we exemplified the downstream analysis in two ways. The first is selecting signatures whose elements capture the highest impact rank in given omic-phenotype associations. The members of such signatures can pertain to different RCD forms (RCD Multi-Modular signatures). The second is selecting signatures that are RCD form-specific.

### 3.1. Exploring Signatures with RCD Multi-Modular Elements

30,877 signatures exhibit multi-modular involvement in RCD, where each gene component within a signature is involved in the same RCD forms. Details of these signatures are available in Dataset S2. A negative correlation between multi-omic and phenotypic features was observed in most signatures (n=17,069). Most multi-modular signatures were overexpressed in tumor tissues compared to non-tumor tissues (n=13,898; Dataset S2). Selected examples of RCD multi-modular signatures are shown in Table 1.

**Table 1.** Top-ranked multi-modular RCD signatures with comprehensive multi-omic representation.

| Rank | Nomenclature | Signature | Elements | Omic feature | RCD count | RCD forms |
| --- | --- | --- | --- | --- | --- | --- |
| 48 | KIRP-1086.1.3.P.3.44.34.1.1.5 | P62LCKLIGAND | 1 | Protein | 5 | Apoptosis, Autophagy, Ferroptosis, Parthanatos, Autosis |
| 48 | KIRC-169.2.1.P.2.71.45.1.1.2 | (CPEB4 + NF2) | 2 | Mutation | 2 | Apoptosis, Ferroptosis |
| 47 | KIRP-419.3.2.N.1.44.114.1.1.2 | (SLC16A1 + SNHG3) | 2 | CNV | 2 | Apoptosis, Autophagy |
| 47 | KIRC-168.2.2.P.2.71.45.1.1.2 | (CPEB4 + NF2) | 2 | Mutation | 2 | Apoptosis, Ferroptosis |
| 47 | KIRP-107.3.2.N.1.44.44.1.1.2 | (CXCL10 + TNFRSF4) | 2 | CNV | 2 | Apoptosis, Necrosis |
| 46 | CESC-215.5.3.N.2.44.44.1.1.3 | (ENST00000511732 + ENST00000471344 + ENST00000559488) | 3 | Transcript | 3 | Apoptosis, Autophagy, Necrosis |
| 46 | KIRP-927.3.2.N.3.15.125.1.1.3 | GBP5 | 1 | CNV | 3 | Apoptosis, Pyroptosis, Necrosis |
| 45 | CESC-283.6.3.N.2.44.44.1.1.3 | (ITGB3 + POSTN) | 2 | mRNA | 3 | Apoptosis, Autophagy, Necrosis |
| 44 | BRCA-2207.5.3.N.3.93.95.1.1.3 | ENST00000518797 | 1 | Transcript | 3 | Apoptosis, Autophagy, Necrosis |
| 44 | BRCA-1496.1.3.P.3.71.71.1.1.2 | CASPASE7CLEAVEDD198 | 1 | Protein | 2 | Apoptosis, Autophagy |
| 44 | CESC-332.6.3.N.2.44.44.1.1.2 | ADAMTS12 | 1 | mRNA | 2 | Apoptosis, Necrosis |
| 43 | BRCA-1629.2.1.P.3.7.44.1.1.2 | CXCR6 | 1 | Mutation | 2 | Apoptosis, Autophagy |
| 42 | SKCM-711.5.3.N.3.71.71.1.1.5 | ENST00000378588 | 1 | Transcript | 5 | Apoptosis, Ferroptosis, NETosis, Parthanatos, Necrosis |
| 42 | CESC-420.6.3.N.2.15.44.1.1.3 | COL1A1 | 1 | mRNA | 3 | Apoptosis, Autophagy, Necrosis |
| 40 | HNSC-1855.4.3.P.3.71.64.1.1.4 | hsa-miR-142-3p | 1 | miRNA | 4 | Apoptosis, Autophagy, Ferroptosis, Necrosis |
| 36 | PRAD-521.5.3.N.2.26.20.1.1.6 | (ENST00000355622 + ENST00000394487) | 2 | Transcript | 6 | Apoptosis, Autophagy, Ferroptosis, Necroptosis, Pyroptosis, Necrosis |
| 35 | BRCA-2459.7.3.N.2.7.94.1.1.2 | FHIT | 1 | Methylation | 2 | Apoptosis, Autophagy |
| 33 | BLCA-576.7.3.N.3.20.5.1.1.6 | AIM2 | 1 | Methylation | 6 | Apoptosis, Autophagy, Cellular senescence, Ferroptosis, Pyroptosis, Necrosis |
| 32 | LUSC-933.4.3.N.2.62.30.1.1.4 | (`hsa-miR-145-3p` +<br>`hsa-miR-145-5p`) | 2 | miRNA | 4 | Anoikis, Apoptosis, Autophagy,<br>Necrosis |
| 31 | BRCA-1824.5.3.N.2.1.1.1.1.6 | ENST00000321556 | 1 | Transcript | 6 | Apoptosis, Autophagy, Cellular<br>senescence, Ferroptosis,<br>Pyroptosis, Mitoptosis |

### 3.2. Exploring signatures with RCD-specific elements

About 13,764 (30.83%) signatures comprise gene elements categorized in only one RCD type. These signatures encompass 20 of 25 different RCD types. Because 81,4% of genes in the inventory are term-based associated with apoptosis, we identified a large number of apoptosis-specific signatures (n = 5,793; 42%) (Dataset S2).

We applied a sequential ranking strategy to identify the most representative signatures that prioritized both performance and comprehensive representation. For each unique RCD form, the most informative signature was selected based on the highest rank value, reflecting the overall importance of the signature. Where multiple signatures shared the same ranking value, ties were resolved by considering the highest value in additional variables in the following order: the number of gene components in the signature, TIC, TMC, SMC, and HRC. This ensured that ties were broken systematically based on biological relevance. We checked whether each omic feature was present in the final selection to represent all unique omic feature comprehensively. If any were missing, the highest-ranked signature for the missing omic feature was added, following the same tie-breaking hierarchy. This method allowed us to generate a ranked list of signatures that reflected their importance and ensured balanced coverage of RCD forms and multi-omic feature. The top-ranked signatures by comprehensive RCD type-specific and multi-omic feature representation are presented in Table 2.

**Table 2.**
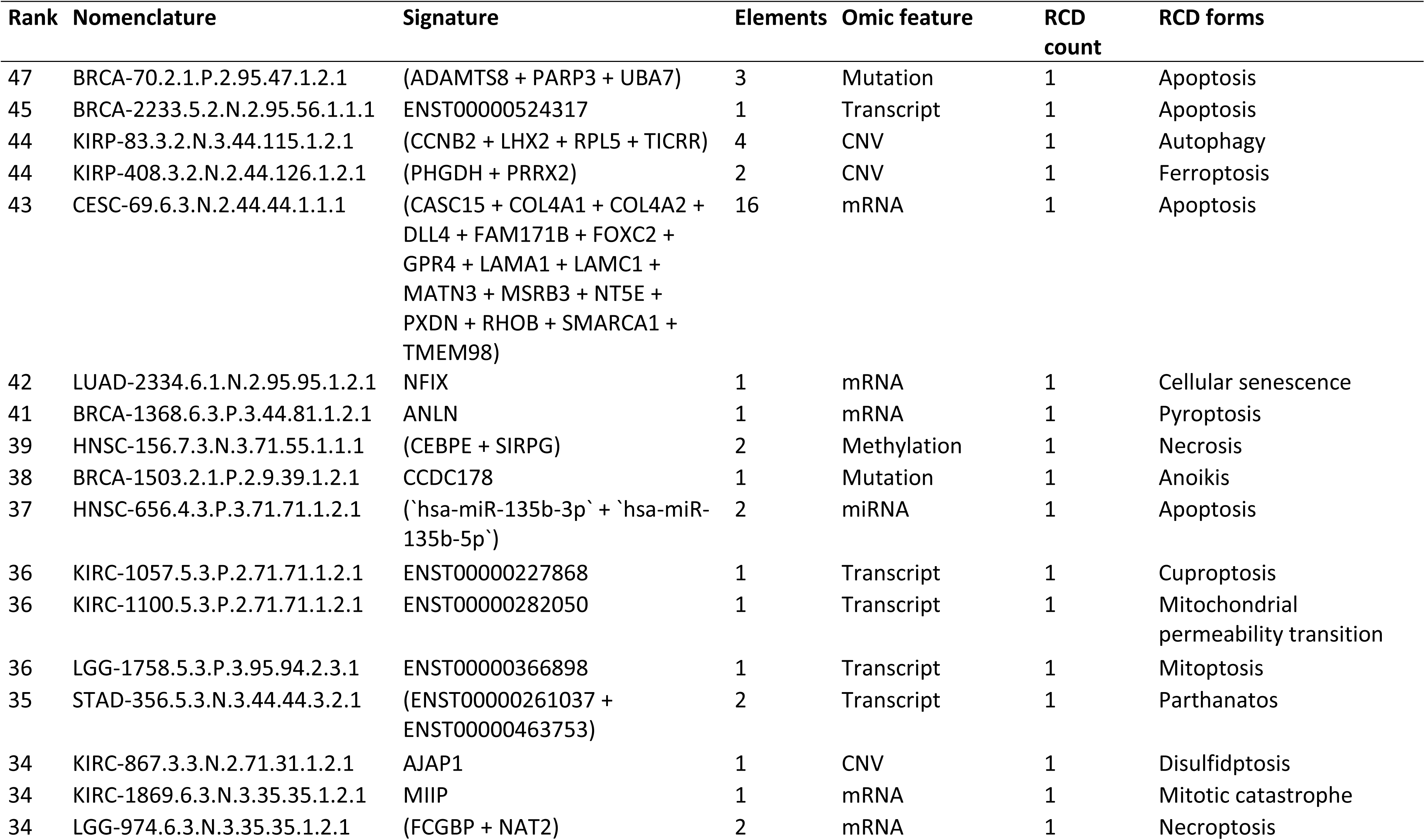
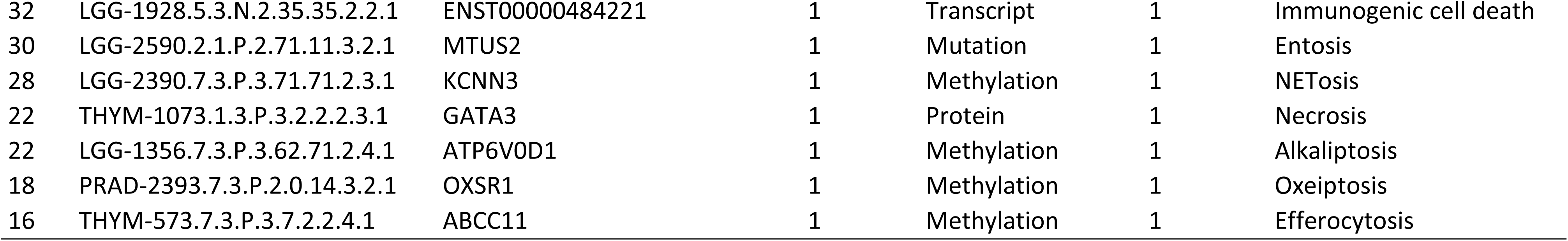
Top-ranked RCD type-specific signatures with comprehensive multi-omic representation.

We next illustrate the clinical meaningfulness potential of the signature database by providing a signature for each omic feature selected from the top-ranked signatures (Table 3).

**Table 3.**
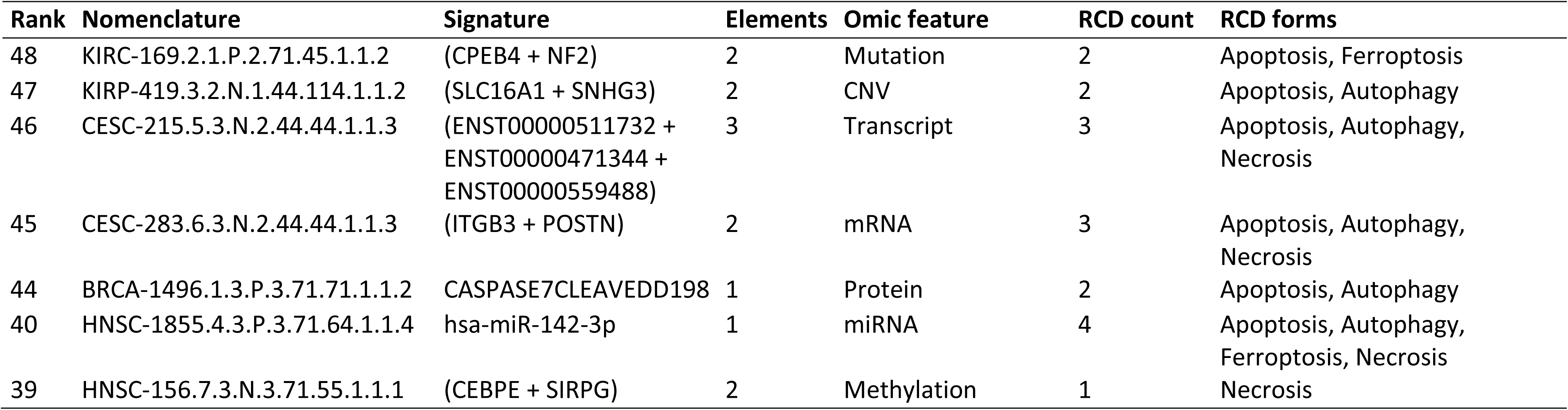
Seven top-ranked signatures by multi-omic feature.

### 3.3. mRNA-specific signatures

We identified 10,096 mRNA-specific signatures (22.6% of the full dataset, Dataset S3), each comprising a range of gene elements per signature from 1 to 477 (mean = 3,8; median = 1; Q1 = 1; Q3 = 2; P90 = 5). Of these, 7,278 signatures (72.1%) were negatively correlated with phenotypic features. Among these, 6,842 signatures (94.1%) showed a correlation with TSM; Within this group, 2,479 (36.2%) signatures were associated with increased risk, while 1,709 (24.9%) were protective based on at least one survival metric.

Among the mRNA-specific signatures, 3,864 (38.3%) were associated with anti-tumoral transcriptional profiles, 2,101 (20.8%) with pro-tumoral profiles, and 2,750 (27.2%) with dual microenvironment profiles, reflecting diverse roles in tumor progression. Based on their correlation with immune cell infiltration profiles, the mRNA-specific signatures were categorized as “hot” (n=273; 2.7%), showing robust immune cell presence, “cold” (n=781; 7.7%), reflecting minimal immune infiltration, and “variable” (n=1,540; 15.2%), denoting an intermediate or mixed immune environment.

The identifier CESC-283.6.3.N.2.44.44.1.1.3 exemplifies a mRNA-specific signature comprising two gene members: *ITGB3* and *POSTN*, associated with apoptosis, autophagy and necrosis (Table 3). These genes play diverse roles in RCD, cell survival, and migration across various cell types, contributing to cancer progression, immune modulation, and cellular stress responses. As part of the same signature, each gene consistently shares correlation signs across all phenotypic features in patients with cervical squamous cell carcinoma and endocervical adenocarcinoma (CESC) (Figure 5). Specifically, mRNA expression levels of these genes exhibit a negative correlation with TSM (Figure 5A), show lower expression in tumor samples relative to non-tumor tissue TSM (Figure 5B), and correlate with risk across all survival metrics (Figure 5C-5F). Elevated expression of these genes is associated with poor prognosis across all survival metrics (Figure 5I-5J). In contrast, their expression profiles correlate with an anti-tumor transcriptional profile within the tumor microenvironment and a “hot” immune infiltrate transcriptional profile (Figure 5K).

**Figure 5.**
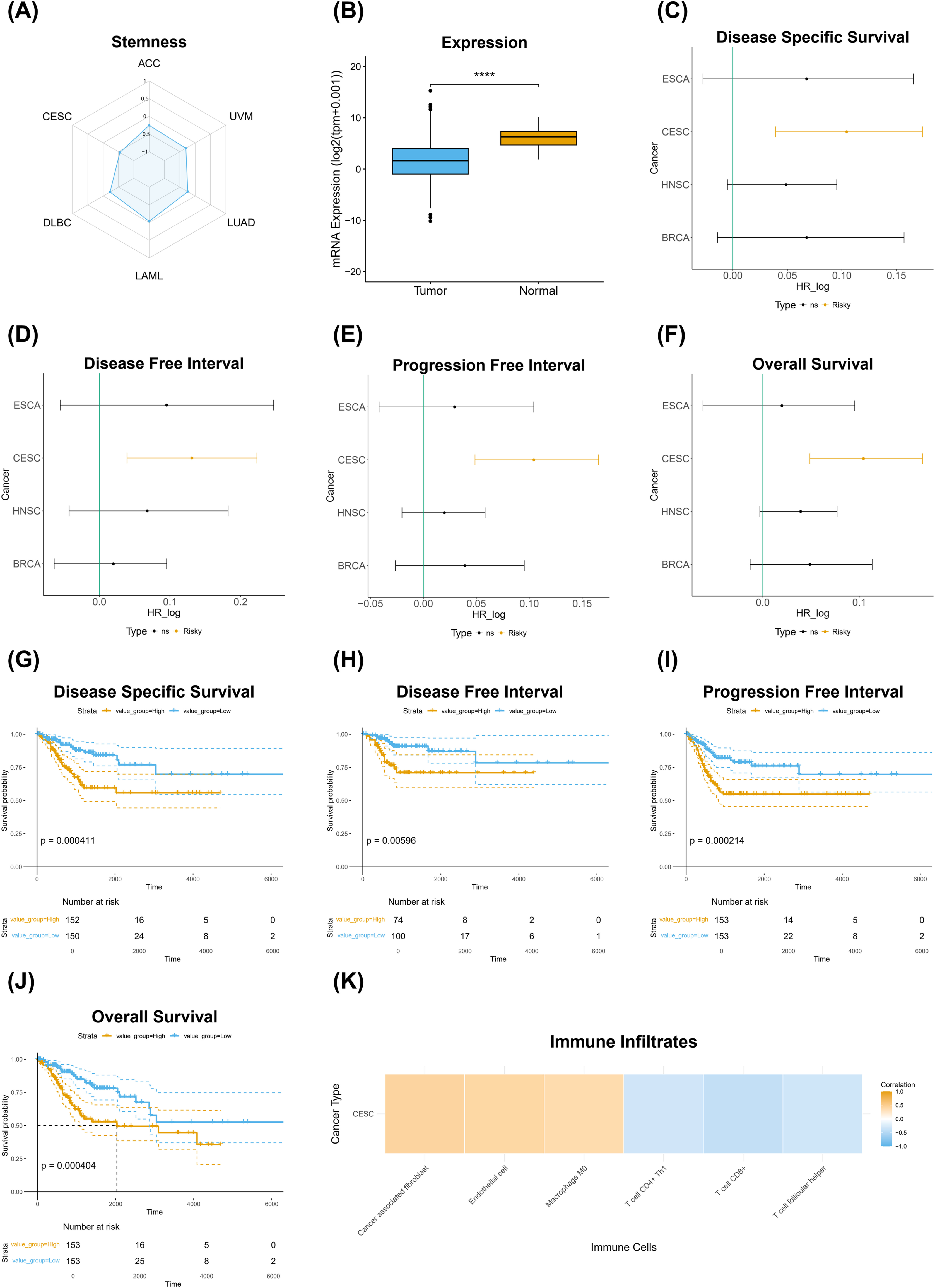
Phenotypic associations and prognostic significance of the mRNA signature CESC-283.6.3.N.2.44.44.1.1.3 in cervical squamous cell carcinoma and endocervical adenocarcinoma (CESC). Panel (A) shows a radar plot illustrating the negative correlation between mRNA signature expression and TSM across multiple cancer types. Panel (B) demonstrates significantly lower mRNA signature expression in tumor samples compared to normal tissue (****p < 0.0001). Panels (C-F) present hazard ratio (HR) analyses evaluating the prognostic associations of the mRNA signature with clinical outcomes across various cancer types, including (C) Disease-Specific Survival, (D) Disease-Free Interval, (E) Progression-Free Interval, and (F) Overall Survival, where a positive log HR indicates a risk effect of the mRNA signature. Panels (G-J) display Kaplan-Meier survival curves for CESC patients stratified by high and low mRNA signature expression, with significant survival outcomes for (G) Disease-Specific Survival (p = 0.000411), (H) Disease-Free Interval (p = 0.00596), (I) Progression-Free Interval (p = 0.000214), and (J) Overall Survival (p = 0.000404). Panel (K) illustrates the correlation between the mRNA signature and immune cell infiltration in CESC, highlighting associations with various immune cell types.

### 3.4. Transcript-level gene signatures

Given that many gene loci express multiple transcripts through alternative splicing and promoter usage, we hypothesize specific transcripts retain the correlation observed in the mRNA analysis. This suggests that individual transcript expression offers more precise insights into cancer progression and therapy response. By analyzing these specific transcripts, we aim to identify transcript-specific signatures that could serve as accurate prognostic and diagnostic markers, enhancing our understanding of the molecular mechanisms and heterogeneity in cancer phenotypes.

We identified 16,244 transcript-specific signatures, with each signature containing between 1 and 2,052 transcript elements (mean = 5.9; median = 1; Q3 = 3 and P90 = 8) (Dataset S4). The mean number of transcript members per signature was 3.9 (range 1 to 49) for signatures that were associated with risk and 4.1 (range 1 to 76) with protection (Dataset S4). Approximately 62.8% (n=10,207) were associated with risk or protection in at least one survival metric. From those, we identified 605 (5.9%) signatures associated with risk across all patient survival metrics and 270 (2.7%) signatures with protective association in all four patient survival metrics. Most signatures ascribed correlations between transcript expression and tumor stemness (86% for risk and 92% for protective signatures). Transcript signature overexpression was the feature most frequently associated with risk (54.5%), whereas underexpression was mainly associated with protection (47%). Example: CESC-215.5.3.N.2.44.44.1.1.3 refers to the transcript expression (ENST00000511732 + ENST00000471344 + ENST00000559488), which negatively correlated with stemness in CESC patients (Figure S3A). There was significantly lower transcript signature expression in tumor samples compared to normal tissue (Figure S3B). Transcript overexpression is associated with risk in DSS (Figure S3C), DFI (Figure S3D), PFI (Figure S3E) and OS (Figure S3F), survival metrics. Transcript signature overexpression was associated with poor prognosis in all survival metrics (Figure S3G-S5J). Transcript signature expression correlated with an anti-tumor transcriptional profile within the tumor microenvironment and a “hot” immune infiltrate transcriptional profile (Figure 3K).

An interesting observation in multi-transcript genes is worth noting; first, the highest number of transcripts per gene that correlated with a phenotype in a cancer type was 19, and limited to the *CD36* (19 out of 24 transcripts), *ABI3BP* (19/29), and *TCF4* (19/93) genes. Second, correlations with all transcript isoforms per gene were extremely rare. Examples are *COL1A1* (known cancer driver gene) with its 13 isoforms negatively correlated with stemness in the multi-element signature HNSC-308.5.3.N.3.0.0.3.2.3 with 46 member elements, and *UMOD* with its 12 transcripts also negatively associated with stemness in the multi-element signature KICH-117.5.3.N.2.0.0.2.4.3 with 61 member elements (Figure 6). Thus, for those signatures, all the *COL1A1- and UMOD-*specific transcripts consistently retained the correlation with stemness. Hence, for those genes, the entire gene loci, rather than individual isoforms, uniformly contribute to the observed phenotype, highlighting a coordinated regulatory role of these genes in maintaining the correlation with stemness. The uniformity across all isoforms within a gene is an uncommon and significant finding, underscoring the comprehensive influence of these genes on the stemness phenotype.

**Figure 6.**
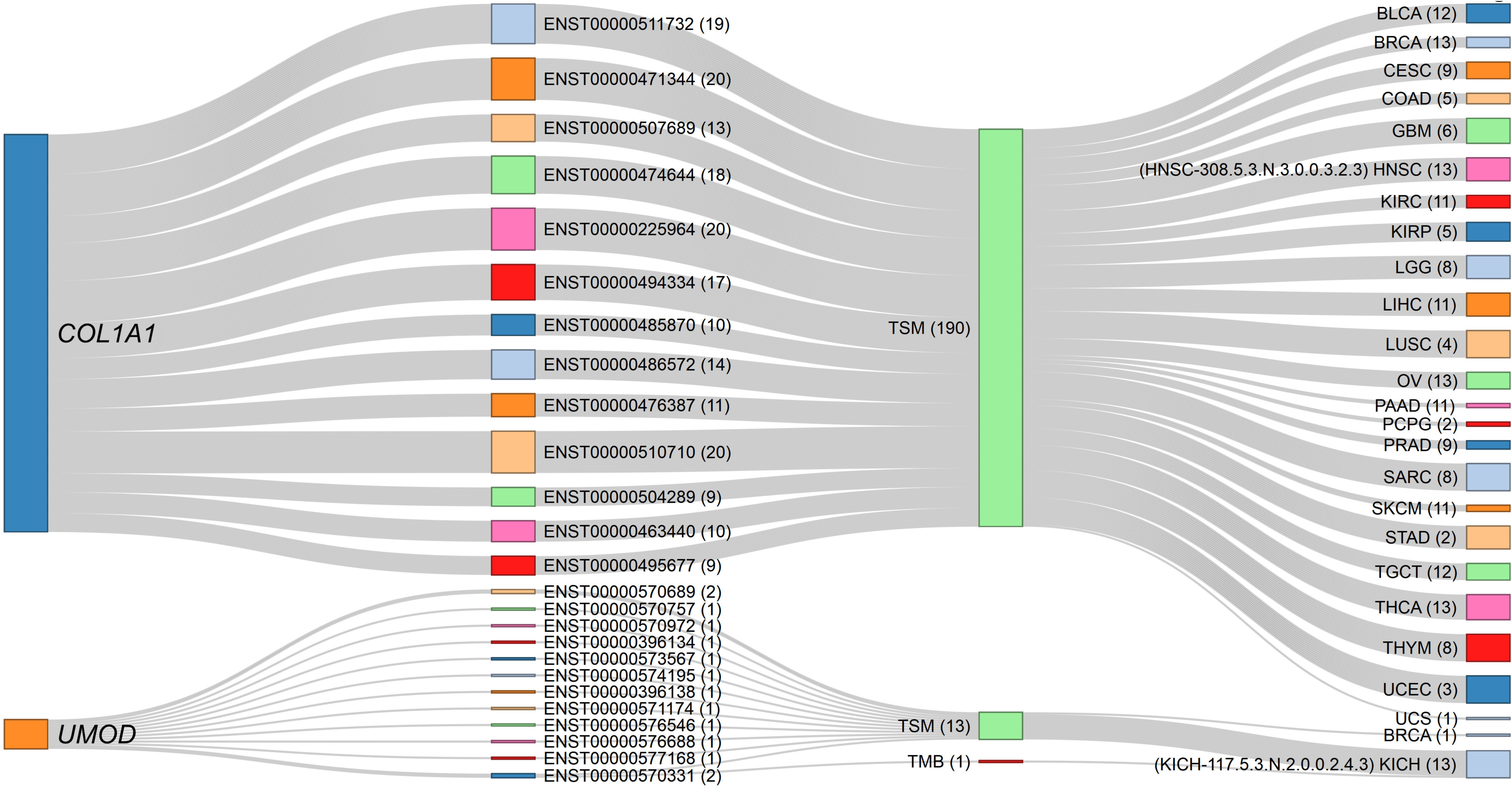
Sankey diagram depicting the negative correlations of *COL1A1* and *UMOD* gene isoforms with stemness across specific cancer signatures. The source nodes represent the *COL1A1* and *UMOD* gene loci, each linked to their respective transcript isoforms identified in the dataset. The numbers in parentheses indicate the number of connection strokes. All 13 *COL1A1* isoforms consistently exhibit negative associations with TSM in the HNSC-308.5.3.N.3.0.0.3.2.3 signature (comprising 46 elements), while all 12 *UMOD* isoforms similarly show negative correlations with TSM in the KICH-117.5.3.N.2.0.0.2.4.3 signature (comprising 61 elements). The thickness of the stroke connection lines represents the frequency of correlations between nodes (genes, transcripts, phenotypes, and cancer types), emphasizing the uniform contribution of each gene’s isoforms to the observed phenotype. This consistent transcript-level correlation across all isoforms of *COL1A1* and *UMOD* suggests a coordinated regulatory function in modulating TSM within these cancer contexts. The corresponding dynamic network diagram is available in Supplementary Material File S1 (Figure S12).

In contrast, for most multi-transcript RCD genes, the correlations were transcript isoform-specific rather than involving the entire gene locus transcript repertoire. Isoform-specific signatures refer to the unique associations of transcript variants from a single gene locus with distinct clinical and phenotypic outcomes. These signatures enable the identification of specific transcript variants that contribute to cancer progression, prognosis, and therapeutic response. Specifically, for the *MAPK10* gene, which has 192 known transcripts, our analysis revealed that only 24 transcripts showed significant correlations with metrics such as TSM, TMB, or MSI across 17 cancer types, appearing in up to 47 different signature identifiers (Figure 7, Dataset S1O). The remaining 180 transcripts from this locus showed no meaningful association. The highest number of *MAPK10* transcript members per signature was 12, observed in LUAD-350.5.3.N.2.0.0.1.4.2. Notably, distinct *MAPK10* transcript isoforms were associated with divergent phenotypes across cancer types. For example, ENST00000486985 expression was positively correlated with MSI in lung squamous cell carcinoma (LUSC) patients (LUSC-1549.5.2.P.1.4.0.4.4.2). In contrast, ENST00000502302 was negatively correlated with TMB in lung adenocarcinoma (LUAD) patients (LUAD-1824.5.1.N.1.0.0.3.4.2). Similarly, ENST00000395169 exhibited a protective role correlating with favorable outcomes in LGG (LGG-1814.5.3.P.3.93.72.2.3.2), whereas ENST00000395160, a different isoform from the same locus, was associated with risk, by four survival metrics, in stomach adenocarcinoma (STAD-1718.5.3.N.1.44.0.3.4.2). These isoform-specific correlations underscore the heterogeneity within the *MAPK10* gene locus, where distinct transcripts contribute variably to cancer progression, phenotypic features, and therapeutic responses across cancer types.

**Figure 7.**
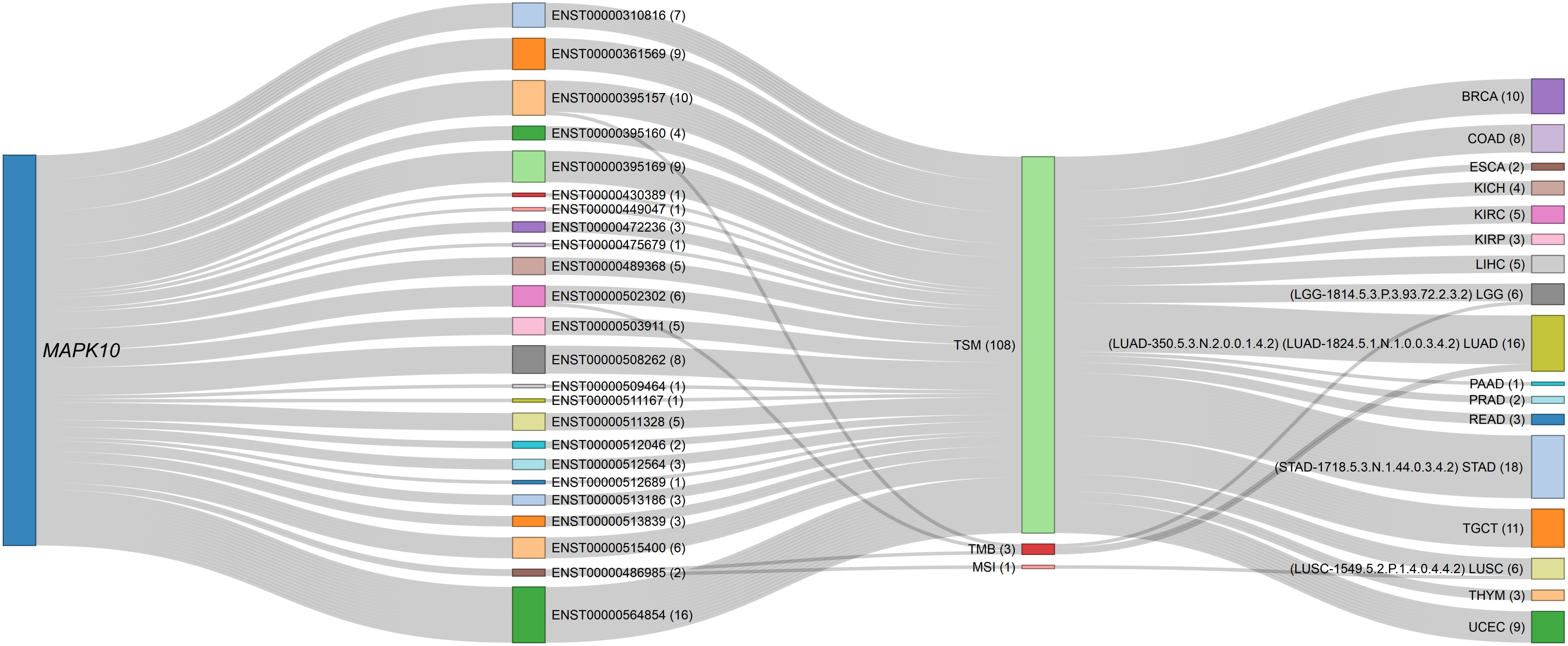
Sankey diagram illustrating transcript-specific associations of the *MAPK10* gene across various phenotypes and cancer types. The *MAPK10* gene locus appears as the source node, connected to its specific transcript isoforms identified in the dataset. Each transcript is further linked to phenotypic profiles (e.g., TSM, TMB, MSI) and mapped to cancer types such as BRCA, COAD, and GBM. The numbers in parentheses indicate the number of connection strokes. The thickness of each link represents the frequency of correlation between *MAPK10* transcripts and the respective phenotypes or cancer types, highlighting both transcript-specific and phenotype-driven associations within multi-transcript gene correlations. For example, the transcript ENST00000486985 (signature identifier: LUSC-1549.5.2.P.1.4.0.4.4.2) shows a positive correlation with MSI in patients with LUSC, while the isoform ENST00000502302 (LUAD-1824.5.1.N.1.0.0.3.4.2) demonstrates a negative correlation with TMB in LUAD patients. The corresponding interactive proportional node dynamic network is available in Supplementary material file S1 (Figure S13).

Table 4 summarizes the transcript-specific correlations of the *MAPK10* gene with cancer types, phenotypic characteristics, and prognostic outcomes, as detailed above. Each transcript is linked to a unique signature identifier, highlighting its distinct role in cancer progression, phenotypic features, and therapeutic relevance.

**Table 4.** Examples of Transcript-Specific Correlations of *MAPK10* with Cancer Types, Phenotypic Features, and Prognostic Outcomes.

| Signature Identifier | Transcript ID | Cancer Type | Phenotypic Correlation | Correlation Direction | Comment |
| --- | --- | --- | --- | --- | --- |
| LGG-1814.5.3.P.3.93.72.2.3.2 | ENST00000395169 | LGG (Lower-Grade Glioma) | Favorable outcomes | Protective | Correlated with better survival outcomes. |
| STAD-1718.5.3.N.1.44.0.3.4.2 | ENST00000395160 | STAD (Stomach Adenocarcinoma) | Poor prognosis | Risk | Linked to worse survival outcomes. |
| LUSC-1549.5.2.P.1.4.0.4.4.2 | ENST00000486985 | LUSC (Lung Squamous Cell Carcinoma) | Microsatellite Instability (MSI) | Positive | Transcript positively correlated with MSI phenotype. |
| LUAD-1824.5.1.N.1.0.0.3.4.2 | ENST00000502302 | LUAD (Lung Adenocarcinoma) | Tumor Mutational Burden (TMB) | Negative | Transcript negatively correlated with high TMB, a hallmark of poor prognosis in LUAD. |

### 3.5. miRNA-Specific Signatures

Our study identified 1,470 miRNA-specific signatures, each composed of 1 to 58 elements (mean = 2.2; median = 1; Q3 = 2; P90 = 4). Of these, 954 (64.9%) contain a single miRNA element. Among the miRNA-specific signatures, 786 (53.5%) correlated with risk or protection in at least one survival metric. Of these, 41 (5.2%) correlated with risk and 16 (2%) with protection in all four metrics of survival. (Dataset S5). The miRNA signatures correlated with distinct tumor microenvironment profiles, including anti-tumoral, pro-tumoral, and variable conditions. We highlight the signature HNSC-1855.4.3.P.3.71.64.1.1.4, which corresponds to hsa-miR-142-3p, the mature form of *MIR142* in head and neck squamous cell carcinoma (HNSC) patients. Cross-referencing public datasets revealed *MIR142* is involved in four RCD forms — apoptosis, autophagy, ferroptosis and necrosis — emphasizing its critical role in hematopoiesis, immune regulation, and cancer progression by modulating various target genes involved in T cell differentiation, inflammation, and tumorigenesis.

hsa-miR-142-3p expression shows a positive correlation with TSM (Figure S4A). It is overexpressed in HNSC tumors as compared with non-tumor tissues (Figure S4B). While hsa-miR-142-3p overexpression was associated with protection in DSS (Figure S4C), PFI (Figure S4E) and OS (Figure S4F), the underexpression was associated with poorer prognosis, as reflected in DSS (Figure S4G), PFI (Figure S4I), and OS (Figure S4J). Furthermore, hsa-miR-142-3p expression was linked to an anti-tumoral profile in the tumor microenvironment, characterized by a “hot” immune infiltrate, indicative of active immune engagement (Figure S4K).

### 3.6. Gene-Specific CpG Methylation Signatures

We identified 6,109 gene-specific CpG methylation signatures, with element counts ranging from 1 to 423 per signature (mean = 3.2; median = 1; Q1 =1; Q3 =2; P90 = 5), of which, 4,246 (69.5%) contain a single CpG Methylation-specific member. The majority (n = 5,350; 87.6%) was associated with TSM. Of these, 192 (3.59 %) were linked to increased risk, while 60 (1.12 %) were protective in all four metrics of survival (Dataset S6). These signatures were further stratified based on their correlation with tumor microenvironment profiles, showing anti-tumoral (n = 98; 38.9 %), pro-tumoral (n = 42; 16.7%), and dual (n = 54; 21.4%) characteristics. The methylation signatures associated with TSM were classified according to their association with immune cell infiltration profiles, showing “hot” (n = 6; 2.4%), “cold” (n = 16; 6.4%), or variable (n = 25; 9.9%) immune phenotypes.

For instance, the signature HNSC-156.7.3.N.3.71.55.1.1.1 demonstrates a negative correlation between CpG methylation at the *CEBPE* and *SIRPG* loci and TSM in HNSC patients (Figure S5A). *CEBPE* and *SIRPG* mRNA expression levels were higher in in tumor than in non-tumor samples (Figure S5B). *CEBPE* and *SIRPG* mRNA expression levels are associated with protection in DSS (Figure S5C), PFI (Figure S5E), and OS (Figure S5F). High methylation levels at *CEBPE* and *SIRPG* are linked to a poorer prognosis in all survival metrics (Figure S5G-S5J). Furthermore, *CEBPE* and *SIRPG* mRNA expression correlates with an anti-tumor microenvironment transcriptional profile and is linked to a “hot” immune infiltration profile in HNSC patients (Figure S5K).

### 3.7. Protein-Specific Signatures

We identified 258 protein-specific signatures, each containing between 1 and 4 elements (mean = 1.1; median = 1; Q1 = 1; Q3 = 1; P90 =1). Of these, the majority (254; 98.5%) exhibited a correlation with TSM, with 153 (60.2%) showing a positive correlation and 101 (39.8%) displaying a negative correlation. Among these, 7 (2.76 %) were associated with increased risk, while 1 (0.4%) was linked to protective effects in all four metrics of survival (Dataset S7). Furthermore, 47 (18.22%) protein-specific signatures correlated with anti-tumoral profiles and 147 (57%) with dual tumor microenvironment profiles. Protein signatures also correlated with immune phenotypes categorized as “hot” (4; 1.3%), “cold” (11; 4.26%), or “variable” (17; 6.6%). Example: BRCA-1496.1.3.P.3.71.71.1.1.2 refers to the expression of the CASPASE7CLEAVEDD198 protein modification, which positively correlated with stemness in BRCA patients (Figure S6A). There was significantly higher mRNA expression for the gene encoding the signature element in tumor samples compared to normal tissue (Figure S6B). Protein overexpression is protective in DSS (Figure S6C), PFI (Figure S6E) and OS (Figure S6F), survival metrics. Low protein expression was associated with poor prognosis in the same survival metrics (Figure S6G, S6I, S6J). Moreover, CASPASE7CLEAVEDD198 expression correlated with anti-tumoral microenvironment and “hot” immune infiltration profiles (Figure S6K).

### 3.8. Mutation-Specific Signatures

We identified 8,022 mutation-specific signatures, each comprising 1 and 487 elements (mean = 3.6; median = 1; Q1 =1; Q3 = 1; P90 = 5), 5,464 (68.1%) of which comprised one element. The majority showed a positive correlation with TMB (5,880; 73.3%) and MSI (2,136; 26.6%), while a small subset (3; 0.04%) with TSM (Dataset S8).

The TMB-associated signatures were linked to risk (n=229; 3.9%), protection (n=96; 1.63 %), “cold” immune cell profiles (n=437; 5.45%), “hot” immune profiles (n = 200; 3.4%), “variable” immune profiles (n= 627; 10.7%), pro-tumoral (n= 687; 11.7%), anti-tumoral (1,426; 24.3%) and dual tumor microenvironment profiles (n=1,501; 25.5%) (Dataset S8). For example, signature KIRC-169.2.1.P.2.71.45.1.1.2, which features the mutation commonalities of *CPEB4* and *NF2* genes, is positively associated with TMB in KIRC patients (Figure S7A). mRNA expression of the signature element was significantly higher in tumor versus non-tumor samples (Figure S7B). mRNA expression of those genes was a protective factor in DSS (Figure S7C), PFI (Figure S7E) and OS (Figure S7F). Mutations in those genes were associated with poor prognosis in all metrics (Figure S7G-S7J). mRNA expression of the signature elements correlated with anti-tumoral microenvironment and “hot” immune infiltration profiles (Figure S7K).

### 3.9. CNV-Specific Signatures

We identified 2,442 CNV-specific signatures, each comprising 1 and 124 elements (mean = 2.4; median = 1; Q1 = 1; Q3 = 2; P90 = 4), 675 (27.6%) of which comprise >1 element. Most of the CNV-specific signatures (1,313; 53.8%) associated with TSM (Dataset S9). Among these, 915 (69.9%) exhibited a negative correlation, while 398 (30.3%) demonstrated a positive correlation. Among the CNV signatures that correlated with TSM, 54 (4.1%) were linked to risk or protection in all four metrics of survival. A portion of these signatures correlated with anti-tumoral (n = 24; 44.4%), pro-tumoral (n = 8; 14.8%) and dual expression profiles (n = 19; 35.2%). These signatures were associated with tumor immune infiltration, characterized as “cold” (n = 11; 20.4%), “hot” (n = 3; 5.6 %) or “variable” (n = 9; 16.7%).

For example, signature KIRP-107.3.2.N.1.44.44.1.1.2, comprising *CXCL10* and *TNFRSF4*, showed CNV negatively correlated with MSI in KIRP (Figure S8A). mRNA expression of the CNV signature constituents was unchanged between tumors and non-tumor samples (Figure S8B). mRNA overexpression of these genes was associated with risk factor in DSS (Figure S8C), DFI (Figure S8D), PFI (Figure S8E) and OS (Figure S8F). Patients with CNV deletions exhibited poor prognosis across all survival metrics (Figure S8G-S8J). Furthermore, mRNA expression of *CXCL10* and *TNFRSF4* was associated with anti-tumoral and “hot” immune microenvironment profiles (Figure S8K).

Table 5 provides a consolidated overview of the classification and distribution of multi-omic signatures, including mRNA, transcript, miRNA, CpG methylation, CNV, mutation, and protein, according to their hazard-risk assessment (risky, protective, or poor prognostic signatures) and their correlation with tumor microenvironment and immune phenotype profiles. These profiles are further categorized based on anti-tumoral, pro-tumoral, or dual microenvironment classifications and immune phenotypes as “hot,” “cold,” or variable. This summary underscores the complexity of prognostic and therapeutic insights derived from distinct multi-omic feature, enabling a deeper understanding of their contextual relevance in cancer research and facilitating the discovery of new biomarkers and therapeutic targets for improved patient outcomes. By integrating diverse molecular features, we highlight the differential associations of multi-omic signatures with tumor prognosis and therapeutic informativeness, defined as the clinical relevance of biomarkers in guiding treatment decisions.

**Table 5.** Classification and Distribution of Multi-omic Signatures by Clinical Outcomes, Hazard Risk, and Therapeutic Informativeness.

| Omic feature | Hazard risk |  |  |  |  | Kaplan-Meier |  |  |  |  | Therapeutic informativeness |  |  |  |  |  |  |  |  |
| --- | --- | --- | --- | --- | --- | --- | --- | --- | --- | --- | --- | --- | --- | --- | --- | --- | --- | --- | --- |
|  | Risky |  |  |  |  | Protective |  |  |  |  | Poorer prognosis |  |  |  |  | TME |  | TIC |  |
|  | Total | DSS | DFI | PFI | OS | DSS | DFI | PFI | OS | DSS | DFI | PFI | OS | Anti-tumoral | Pro-tumoral | Dual | Hot | Cold | Variable |
| mRNA | 10096 | 2372 | 1136 | 2207 | 2400 | 1414 | 728 | 1344 | 1151 | 2734 | 1405 | 2630 | 2733 | 3864 | 2101 | 2750 | 273 | 781 | 1540 |
| Transcript | 16244 | 3737 | 1987 | 3527 | 3724 | 2291 | 1393 | 2231 | 1968 | 4485 | 2535 | 4352 | 4428 | 4176 | 4222 | 6360 | 368 | 968 | 2006 |
| miRNA | 1470 | 291 | 121 | 249 | 248 | 190 | 98 | 161 | 185 | 328 | 165 | 301 | 310 | 572 | 175 | 537 | 35 | 37 | 141 |
| Mutation | 8022 | 1900 | 957 | 1850 | 1802 | 987 | 558 | 1106 | 1022 | 722 | 312 | 1165 | 679 | 2003 | 1000 | 2175 | 251 | 437 | 913 |
| CNV | 2442 | 591 | 323 | 519 | 562 | 371 | 160 | 350 | 259 | 879 | 492 | 827 | 717 | 724 | 316 | 746 | 74 | 148 | 403 |
| Methylation | 6109 | 1255 | 605 | 1241 | 1289 | 730 | 410 | 722 | 618 | 1211 | 753 | 1327 | 1460 | 1632 | 850 | 1708 | 197 | 419 | 711 |
| Protein | 258 | 37 | 14 | 39 | 37 | 18 | 4 | 22 | 27 | 43 | 20 | 46 | 49 | 47 | 0 | 147 | 4 | 11 | 17 |
| Total | 44641 | 10183 | 5143 | 9632 | 10062 | 6001 | 3351 | 5936 | 5230 | 10402 | 5682 | 10648 | 10376 | 13018 | 8664 | 14423 | 1202 | 2801 | 5731 |

### 3.10. Clinically meaningful signatures

By integrating diverse molecular features, we identified 167 clinically meaningful signatures across five omic features: Transcript, mRNA, CNV, Methylation, and Mutation. These signatures were derived from analyses of 11 cancer types, including STAD, PRAD, LUSC, LUAD, LGG, KIRP, KIRC, HNSC, CESC, BRCA, and ACC (Dataset S1P). The selection process focused on signatures with significant associations with hazard risks and prognostic metrics across all four survival outcomes: DSS, DFI, PFI and OS. These signatures also showed robust correlations with immune infiltration profiles, which were categorized into anti-tumoral, pro-tumoral, or dual microenvironment roles, and immune phenotypes classified as “hot,” “cold,” or “variable”. Among these, the top most clinically significant signatures are presented in Table 6.

**Table 6.**
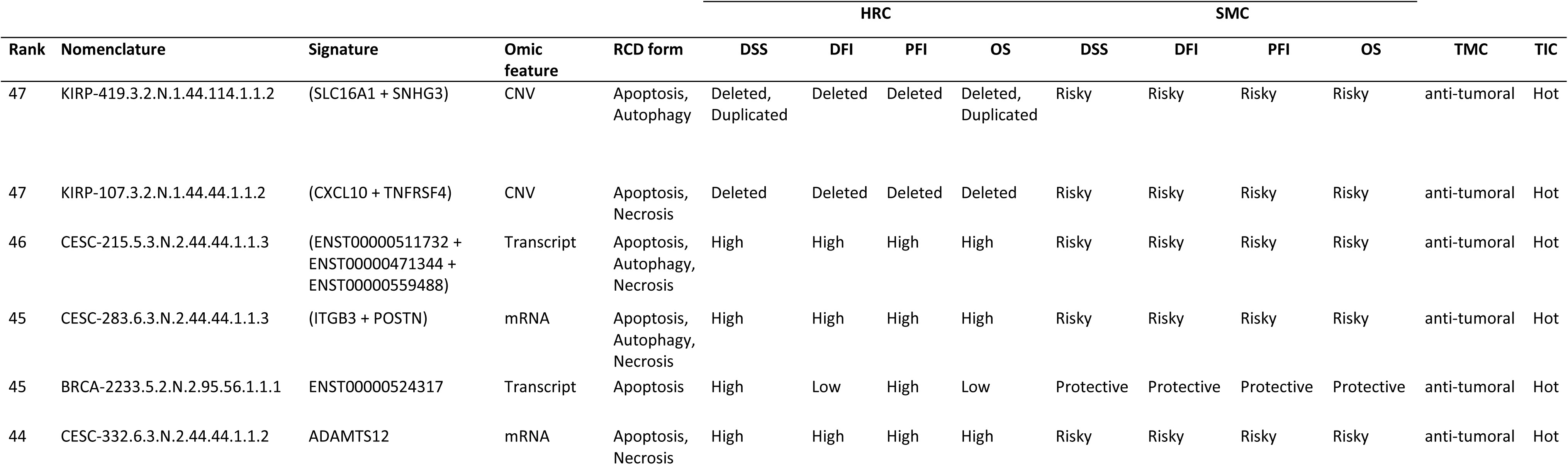
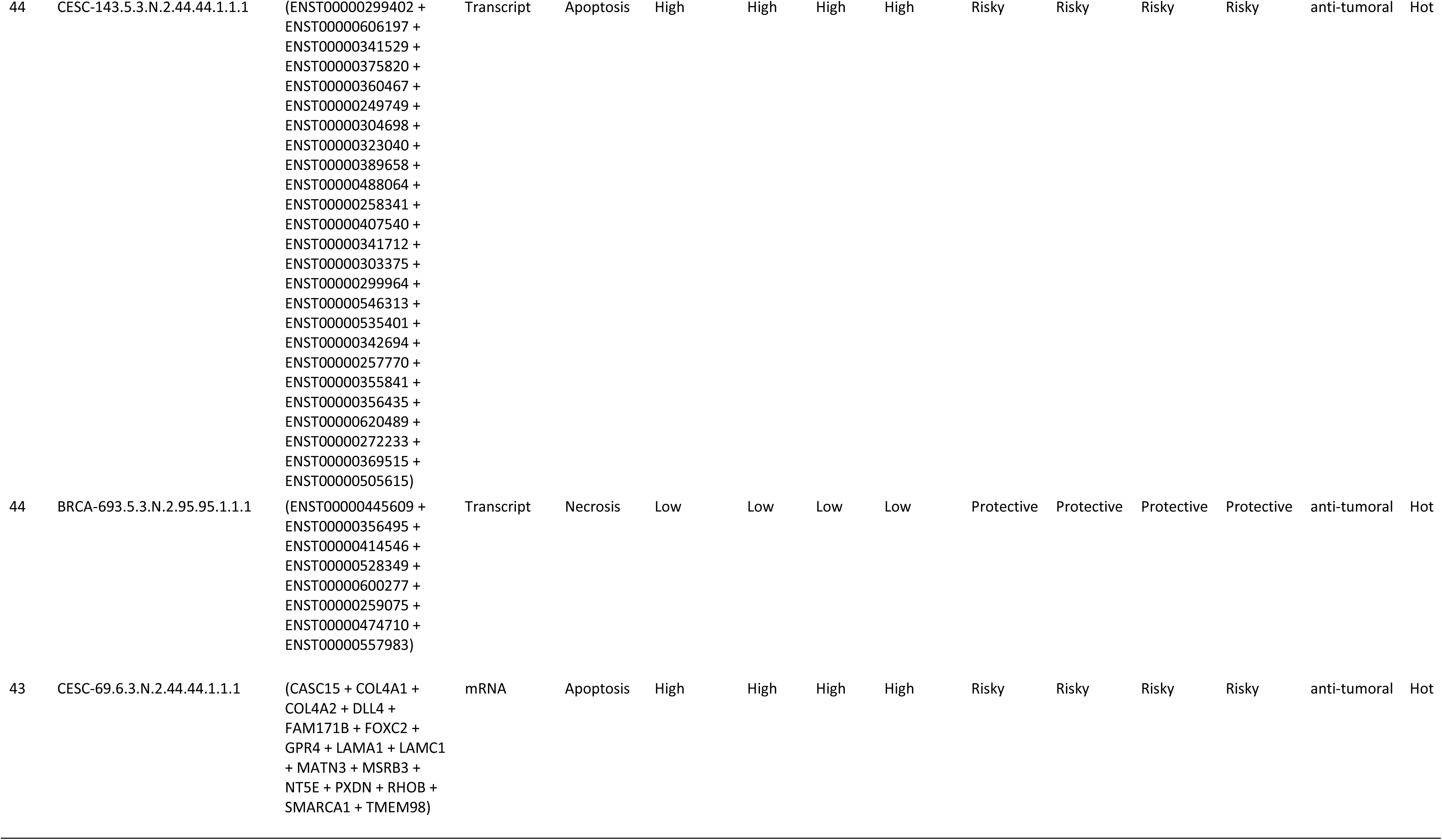
Top most clinically meaningful signatures.

### 3.11. Identification of Potential Therapeutic Targets Through Known Drug-Gene Interactions in Top-Ranked Gene Signatures

To identify potential therapeutic targets, we analyzed the gene components of the leading multi-modular signatures (Table 1), RCD type-specific signatures (Table 2), multi-omic feature signatures (Table 3) and top clinically meaningful signatures (Table 6). Collectively, these top 45 signatures (Dataset S1Q) encompass 84 distinct genes (Dataset S1R). By inputting this list into the DGIdb [91], we found that 27 of the 84 genes are associated with 146 known drug interactions, as detailed in Dataset S1S. Notably, 59.6% (n=87) of these interactions involve drug inhibitors. The genes with the highest number of drug interactions include *APBB1*, *NAT2*, *ITGB3*, *RHOB*, *TLR4*, *ATP5F1A*, *TNFRSF4*, *GATA3*, *PARP3*, *RPL5* (Figure S11).

#### 3.1.2. Independent Validation of Prognostic Signatures Using PRECOG

To ensure the robustness and generalizability of our findings, we assessed the prognostic value of 126 top, clinical meaningful, mRNA-specific signatures (Dataset S1K) using the independent PRECOG database. Of the 126 signatures selected for their association with risk, protection, and poor prognosis—as well as their links to anti-tumoral, pro-tumoral, or dual microenvironment cell profiles and immune infiltrates—we successfully validated 73 signatures in five PRECOG cancer types (Lung cancer ADENO, Breast cancer, Brain cancer Glioma, Brain cancer Astrocytoma, and Prostate cancer). These PRECOG cancer types are equivalent to the TCGA cancer types LUAD, BRCA, LGG, and PRAD, in which these signatures were initially identified (Figure 9). This validation underscores the clinical relevance of these signatures and their potential utility in diverse patient populations.

**Figure 8.**
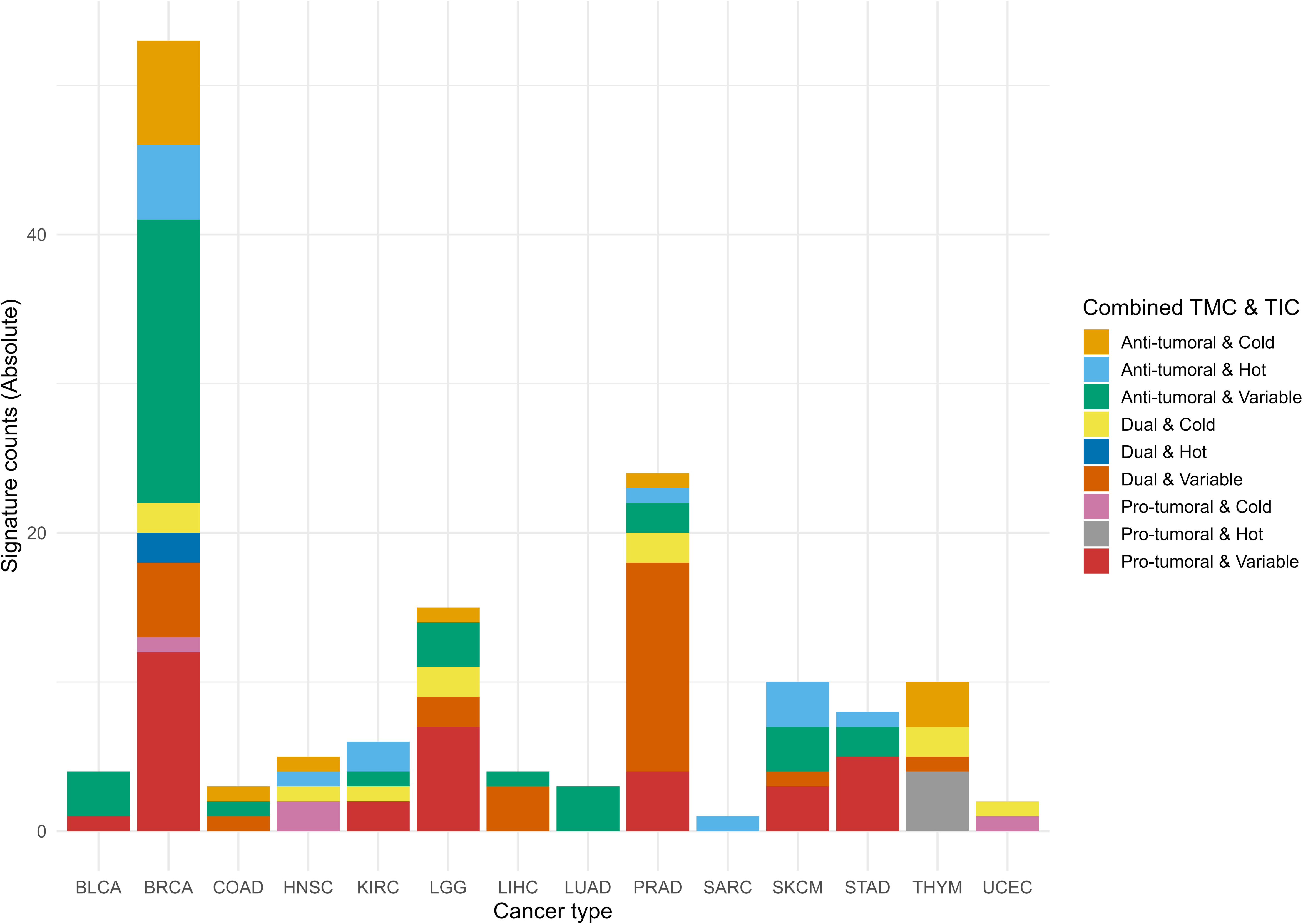
Accumulated histogram illustrating the distribution of mutation-specific signatures by meaningful immunotherapy potential across cancer types. The histogram shows the absolute counts of signatures associated with the combined Tumor Microenvironment Code (TMC) and Tumor Lymphocyte Infiltrate Code (TIC) ranks. Each bar represents a specific cancer type (CTAB), and segments within the bars show the distribution of combined ranks categorized as Anti-tumoral & Hot, Dual & Variable, and Pro-tumoral & Cold, among others. The colors correspond to the Combined TMC & TIC ranks, mapped using the Okabe-Ito color palette extended for color-blind friendliness. Data were processed and summarized from multi-omic analyses of mutation-associated signatures with the potential for immunotherapy (Dataset S1W).

**Figure 9.**
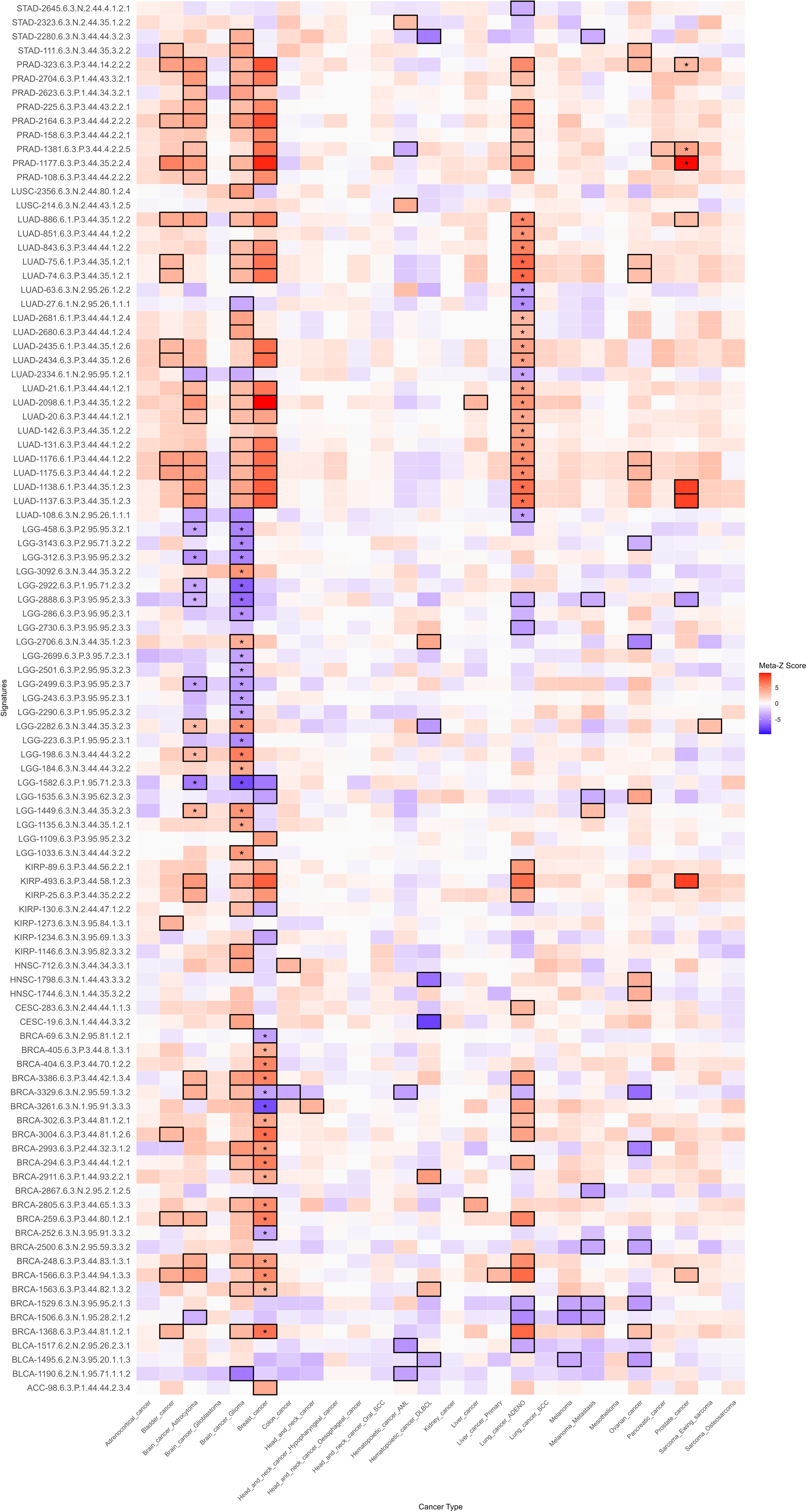
Heatmap of prognostic meta-Z scores from the PRECOG independent database. This heatmap illustrates the association between mRNA-specific signatures (y-axis) and PRECOG cancer types (x-axis). Median meta-Z scores were computed based on overall survival (OS) metrics. Cells are color-coded: blue denotes a favorable prognosis (negative meta-Z scores), red indicates a poor prognosis (positive meta-Z scores), and gray represents neutral or non-significant values. Black-bordered cells highlight statistically significant associations (|Meta-Z| > 3.09 or < -3.09, p < 0.001). The asterisk within the black-bordered cells marks signatures whose prognostic values were validated in both direction and strength in TCGA-equivalent PRECOG cancer types.

### 3.13. *CancerRCDShiny*: Exploring Multi-omic Signatures in RCD for Cancer Research

We implemented *CancerRCDShiny* (https://cancerrcdshiny.shinyapps.io/cancerrcdshiny/), a tool designed to facilitate the exploration and analysis of signatures associated with RCD forms in cancer. This R Shiny app is tailored for researchers and clinicians aiming to uncover the molecular underpinnings of cancer through the lens of cell death processes. *CancerRCDShiny* integrates a robust database encompassing 25 distinct RCD forms and 32 cancer types, allowing users to delve into the intricate relationships between signatures and cancer phenotypes. The app employs rigorous genome-wide significance filters to identify the most relevant signatures, ensuring access to high-confidence data for thorough analysis. Users can explore multiple gene features and phenotypic attributes, providing a comprehensive view of the genetic landscape associated with RCD in cancer. The app offers a user-friendly interface with dynamic visualization tools, allowing users to navigate data easily, create custom plots, and generate detailed reports. Researchers can tailor their queries to specific RCD forms, cancer types, or omic features, facilitating targeted investigations. *CancerRCDShiny* is an essential resource for precision oncology, empowering researchers to uncover novel insights and advance cancer research.

*CancerRCDShiny* also contains an *RCD Multi-omic Signature Identifier Interpreter* function that deciphers the complex nomenclature of the signatures. This function allows the user to paste any RCD signature identifier from the database and download the interpreted identifier in text format.

### 3.14. Performance of the Cancer Regulated Cell Death Data Analyst tool

The Cancer Regulated Cell Death Data Analyst is a specialized GPT-based software tool designed to extract and process information from various file formats, generating structured tabular outputs in .csv format to address specific research queries related to regulated cell death (RCD) in cancer. It offers robust capabilities, including automated data cleaning, integration with external databases, and natural language processing (NLP) techniques for extracting insights from unstructured text. The tool supports interactive dashboards for real-time visualization, functional annotation and enrichment analysis, predictive modeling using machine learning, and customizable reporting. Additional features include secure user authentication, data encryption, API access for seamless integration with other software tools, collaborative functionalities for team-based analysis, version control for data and workflows, and educational resources. It also provides advanced R code suggestions for in-depth analysis, such as data visualization through plots and images, tailored specifically for RCD research. A built-in feedback mechanism ensures continuous improvement, while enhanced plotting and imaging capabilities further refine data interpretation. The tool can be accessed at URL: https://chatgpt.com/g/g-8etzMPrtt-cancer-regulated-cell-death-data-analyst.

## 4. Discussion

### 4.1 Holistic Approach and Context-Specific Analysis

Our multi-optosis model is integrative and holistic, querying 5,913 genes associated with RCD, encompassing 62,090 transcripts, 882 matured miRNAs and 239 cancer-associated proteins and protein modifications (for 193 genes) from 25 distinct RCD forms. The model assumes non-uniformity in the activity and effects of the RCD gene components across different cancer types. Each cancer type is analyzed separately, ensuring that the unique biological contexts and specific molecular mechanisms of each cancer type are thoroughly considered. When querying target genes, we treat the RCD gene inventory as a whole; however, each gene is conceptually tagged to one or more RCD forms. This approach allows us to account for each cancer type’s unique biological contexts and specific molecular mechanisms, ensuring a comprehensive understanding of the associations between RCD gene partners in cancer progression and treatment resistance. By combining these elements, our model uncovers new biomarkers and therapeutic target candidates, opening avenues for more effective cancer treatments.

The signature database developed in this study offers a valuable resource for advancing cancer research and treatment through multiple RCD signaling pathways. The signature identifiers are enriched with meaningful information encoded in the nomenclature system, unveiling hidden correlations between multi-omic and phenotypic features. Our rank-scoring system integrates multiple critical factors to assess each signature’s overall significance and correlation, providing preliminary solid evidence for their prognostic value. This method offers a comprehensive framework for evaluating signatures in cancer research. Our method ensures a balanced and accurate assessment of each signature’s relevance by considering multiple factors, including cancer type, survival metrics, and multi-omic and phenotypic features. The scoring of signatures can facilitate the prioritization of signatures for further investigation, ultimately accelerating the discovery of actionable insights and improving patient outcomes.

Currently, no signature identifier system in the literature incorporates multi-omics features as comprehensively as in our model. Most studies on signatures related to RCD and cancer list signatures based on a single type of association [40, 97–108]. Our model’s novel integration of multi-omics features and multiple phenotypic attributes provides a more holistic and informative framework for understanding cancer biology. The uniqueness of our integrated multi-omic, multi-feature signature discovery approach significantly enhances the potential of key biomarkers and therapeutic targets.

However, we recognize the necessity of thorough validation across independent datasets to confirm the reliability of these signatures in clinical settings. Therefore, validation in independent datasets is required to establish our findings’ clinical applicability fully.

### 4.2 Interpretation of Multi-Omic and Phenotype Correlations and Their Signs

The signatures identified through specific multi-omic and phenotype correlations are candidate proxies for diagnosis, prognosis, or therapeutic response. Integrating positive and negative correlations into our analysis provides a more comprehensive understanding of the signatures associated with various cancer phenotypes. This thorough approach allows for the identification of potential oncogenes and tumor suppressors, which can pave the way for the creation of more tailored and effective cancer treatments. For instance, positive correlations between gene expression and tumor stemness could indicate aggressive cancer phenotypes, metastasis, and therapy resistance. Examples include overexpression of the pluripotency- and apoptosis-related *POU5F1* (*OCT4*) (i.e., TGCT-15.6.3.P.3.0.0.2.4.2), *SOX2* (i.e., LUSC-2293.6.3.P.3.62.30.1.4.3), and *NANOG* (i.e., TGCT-4.6.3.P.3.0.0.2.4.1) genes in various cancer types associated with stemness and poor prognosis [109–117].

Negative correlations provide equally critical insights. A negative correlation between gene CNV and a particular phenotype (i.e., stemness) could indicate genes that suppress aggressive traits or resistance mechanisms. For example, *TP53* deletion/duplication was correlated with poor prognosis in Liver hepatocellular carcinoma (LICH) patients (LIHC-1867.3.3.N.3.0.126.1.4.12). *TP53* plays a crucial role in DNA repair and apoptosis [118].

The link between *TP53* CNV changes and adverse outcomes in LIHC suggests its potential as a marker for high-risk patients. Loss of *TP53* function can weaken DNA repair and apoptosis, facilitating tumor progression. This underscores *TP53*’s role in restraining tumor aggressiveness and highlights it as a potential therapeutic target. Exploring similar negatively correlated genes can reveal critical mechanisms in cancer suppression and inform targeted therapies.

Identifying genes that are negatively correlated with aggressive tumor features can highlight potential tumor suppressors or biomarkers of less aggressive disease. For example, reduced levels of E-cadherin (*CDH1*) are associated with increased invasiveness and metastasis in several cancers [119]. We identified 16 signatures with *CDH1*, which is overexpressed in ten cancer types (PAAD, COAD, BRCA, LGG, HNSC, THYM, STAD, READ, GBM, and PRAD). In head and neck squamous cell carcinoma (HNSC) patients, *CDH1* mRNA expression was found to be negatively correlated with MSI (HNSC-814.6.2.N.3.0.0.2.2.3) (Dataset S3). Notably, *CDH1* mutations in HNSC patients are rare (Dataset S1T) compared to *TP53* mutations (Dataset S1U and Figure S9).

This negative correlation suggests that higher *CDH1* expression may contribute to tumor stability and cohesion, consistent with its role as a tumor suppressor and adhesion molecule. In HNSC, where high MSI frequently correlates with aggressive behavior and poor prognosis, elevated *CDH1* expression may help preserve cellular integrity, potentially limiting the tumor’s invasive capacity. This aligns with *CDH1*’s function in stabilizing cell-cell interactions and opposing epithelial-mesenchymal transition, a process often heightened in MSI-high tumors [119, 120].

The presence of solo *CDH1* signatures across different cancer types further underscores *CDH1*’s potential as a marker of epithelial integrity and reduced invasiveness, especially in tumors with a preserved epithelial phenotype. Using *CDH1* as an indicator of cellular cohesion could improve patient stratification, identifying patients who may benefit from therapies focused on maintaining cell adhesion and curbing invasion-related pathways. This finding supports the need for further research into *CDH1*’s role in tumor stability and its broader potential as a biomarker across cancer types.

### 4.3. Prognostic and Diagnostic Potential of Transcript-Specific Signatures

The transcript-level correlations observed for *MAPK10* underscore the importance of distinguishing specific isoforms in cancer research and clinical applications. While *MAPK10* as a gene locus shows variability across cancer types, individual transcripts reveal distinct associations with phenotypic features and patient outcomes. This specificity highlights the potential for isoform-level resolution to refine prognostic tools and therapeutic strategies. For instance, identifying protective or risk-associated transcripts can improve the accuracy of patient stratification, enabling more personalized treatment plans.

The distinct roles of *MAPK10* isoforms in tumor progression and microenvironment interactions also emphasize the need for targeted therapeutic approaches. By isolating isoforms associated with pro-tumoral or anti-tumoral phenotypes, therapies could be designed to selectively modulate these transcripts, maximizing treatment efficacy while minimizing off-target effects. This approach could be precious in cancers where *MAPK10* isoforms contribute differentially to immune infiltrates, such as "cold" or "hot" tumors, potentially guiding the selection of immunotherapy strategies.

The heterogeneity within *MAPK10* reinforces the importance of transcriptomics in understanding cancer biology. Whole-gene analyses may overlook critical isoform-specific contributions that drive tumor behavior and therapeutic response. As such, incorporating transcript-level data into clinical workflows could enhance diagnostic precision, prognosis accuracy, and the development of isoform-targeted therapies, representing a significant advancement in precision oncology.

### 4.4. Application and Translational Potential in Clinical Settings

Cancer therapies, including immunotherapy, aim to eliminate cancer cells, with their success often influenced by genes that regulate cell death. Most RCD-associated genes play either a pro-RCD or an anti-RCD role. However, depending on the cancer context, specific RCD-associated genes can promote or inhibit cell death, affecting their suitability as therapeutic targets. It is also known that some genes exhibit dual roles, acting as pro-RCD or anti-RCD agents based on the cancer type. For example, *SLC7A11*, which codes for a component of a sodium-independent, anionic amino acid transport system specific for cysteine and glutamate, promotes resistance to ferroptosis in gliomas (e.g. LGG-2956.3.3.N.3.0.0.2.4.4) but inhibits ferroptosis in endometrial carcinoma (e.g. UCEC-1106.2.1.P.3.2.0.2.4.4) [115, 117, 121] (Dataset S1V).

In cancer treatment, genes that promote RCD are often considered desirable targets because they facilitate the elimination of cancer cells. Conversely, genes that inhibit RCD can contribute to therapy resistance, making them challenging targets in specific cancers. Customizing therapeutic strategies based on the gene’s role in RCD within the specific cancer type can optimize treatment outcomes. Therapies should be aligned with whether a gene’s function is to promote or inhibit cell death, ensuring that the approach enhances the effectiveness of the treatment.

The RCD signature database holds significant promise for practical application in preclinical settings, offering valuable tools for patient stratification, personalized treatment plans, prognostic applications, and therapeutic decision-making. These signatures may enable categorizing patients based on their molecular profiles, leading to more tailored and effective treatment strategies.

We identified 148 gene signatures with somatic mutations positively correlated with TMB and with immunotherapeutic potential by their association with immune infiltrate profiles in fourteen cancer types (Dataset S1W). The distribution of these mutation-specific signatures by cancer type and their immunotherapeutic potential is shown in Figure 8. Mutations in these genes are likely sources of neoantigens, as high TMB produces more immunogenic mutations [122]. This connection suggests the mutation-specific signatures could identify neoantigen targets for personalized therapies, such as cancer vaccines or T cell-based treatments.

The practical application of our findings lies in stratifying patients by using the signatures as prognostic tools to guide therapeutic decisions based on the cancer’s molecular profile [123]. To bring our multi-omic signature database into clinical practice, it is essential to conduct rigorous clinical trials that validate both its efficacy and reliability. This involves evaluating the predictive power of the signatures across diverse patient cohorts and confirming reproducibility in different clinical settings [123]. Notably, cross-referencing the gene components in the database with existing literature reveals that some signatures or their members have already been evaluated in previous preclinical studies, which highlights the translational potential of our findings, bridging preclinical insights with clinical applications. Out of the 150 widely recognized immunological targets in cancer research, 91 (60,7%) are included in the signatures identified in this study (Dataset S1L).

### 4.5. Cases of clinically validated RCD multi-omic signatures

We identified 879 multi-omic signatures (Dataset S10) that contain at least one gene member from 27 out of 29 genes whose protein products are classified as chimeric antigen receptor (CAR) targets and are currently under investigation in clinical trials as identified by Clinicaltrials.gov [124].

We exemplify the translational impact of the RCD multi-omic signature database with two cases in which members of the multi-optosis signatures have been clinically validated in independent studies. The first case is *CD274* (a driver gene that encodes for PD-L1) [125]. The finding that *CD274* is involved in eight RCD processes (apoptosis, autophagy, cuproptosis, efferocytosis, ferroptosis, necroptosis, pyroptosis, and necrosis) expands our understanding of the multifaceted roles of PD-L1 in cancer biology (Dataset S1B). This broad involvement suggests that PD-L1 may influence tumor progression and response to therapy through multiple pathways, not just immune evasion. This knowledge can lead to targeted and effective therapeutic strategies that address these various pathways.

The positive correlation between *CD274* mutations and TMB in GBM-410.2.1.P.3.35.0.4.4.8, LGG-1442.2.1.P.3.35.0.3.4.8, and PAAD-773.2.1.P.3.42.0.2.4.8 suggests that higher TMB, often associated with better responses to immunotherapy, is linked to the occurrence of *CD274* mutations (Figure S10, Dataset S1X). Given the low frequency of *CD274* somatic mutations in those cancer types (0.4%; 4 mutations in 998 patients, Dataset S1X), as compared to *TP53*, a prominent driver cancer gene (Dataset S1Y), the findings highlight that even rare mutations can have significant clinical implications. Patients with high TMB are more likely to have neoantigens that enhance the immune response, making them better candidates for immunotherapy. This correlation can guide the selection of patients for immune checkpoint inhibitor therapies, potentially leading to better clinical responses.

The identification of *CD274* mutations as a risk factor in at least one survival metric in patients with GBM, LGG, and PAAD (Figure S9) aligns with previously reported associations [126–128]. It reinforces the role of PD-L1 as a critical biomarker for patient stratification in these cancers. Clinicians can assess PD-L1 expression levels to identify patients more likely to benefit from PD-L1/PD-1 checkpoint blockade therapies, thus personalizing treatment plans and improving outcomes.

Understanding the diverse roles of *CD274* in various RCD processes (apoptosis, autophagy, cuproptosis, efferocytosis, ferroptosis, necroptosis, pyroptosis, and necrosis) (Dataset S1B) and its positive association with TMB can inform the development of combination therapies. For instance, combining PD-L1 inhibitors with agents targeting specific RCD pathways (e.g., ferroptosis inducers or necroptosis inhibitors) could enhance therapeutic efficacy by simultaneously disrupting multiple tumor survival mechanisms.

The second case example is the *AXL* receptor tyrosine kinase, which plays critical roles in cellular functions such as growth, migration, aggregation, and anti-inflammation in multiple cell types [129], and it is term-based associated with apoptosis, efferocytosis, necroptosis, and necrosis in various cancer types (Dataset S1B). Our findings show that *AXL* mRNA expression is negatively correlated with stemness in PAAD-718.6.3.N.3.44.43.3.4.4, LGG-1367.6.3.N.3.35.35.2.4.4, and STAD-1167.6.3.N.3.35.1.3.2.4 patients (Dataset S3). Specifically, these signatures indicate that high *AXL* expression is linked to decreased stemness in these cancers. Conversely, in PAAD and STAD patients, *AXL* mutations show a positive correlation with TMB (Dataset S3). The frequency of *AXL* somatic mutations in those cancer types is low (5.5%; 34 mutations in 616 patients, Dataset S1Z) compared to *TP53* (Figure S9, Dataset S1AA). Thus, adverting that even infrequent mutations can have significant clinical implications. Notably, the overexpression of *AXL* in these three cancer types is a risk factor across three to four metrics of patient survival (Dataset S3).

Cross-referencing shows that anti-human monoclonal antibodies targeting the AXL receptor tyrosine kinase inhibit AXL activity effectively, limiting the proliferation and migration of pancreatic cancer cells *in vitro* and *in vivo* [130]. This evidence suggests a promising approach for immunotherapy in PAAD, LGG, and STAD patients, underscoring the potential for these signatures to inform innovative therapeutic strategies involving anti-AXL antibodies and small molecule AXL kinase inhibitors. Dysregulated *AXL* expression in STAD is highlighted as a promising therapeutic target, further supporting the relevance and potential impact of targeting AXL in gastrointestinal cancers [131].

### 4.7. Validation in PRECOG cancer types

While 73 of the 126 mRNA-specific signatures were successfully validated in PRECOG cancer types equivalent to TCGA (Lung adenocarcinoma, Breast cancer, Glioma, Astrocytoma, and Prostate cancer), the incomplete validation rate (58.4%) highlights important biological and technical considerations. Several factors may explain why not all signatures showed consistent prognostic value across the independent PRECOG dataset: (1) lack of equivalent TCGA vs PRECOG cancer type (example: CESC - Cervical squamous cell carcinoma and endocervical adenocarcinoma; n = 5 signatures); (2) gene absent in PRECOG (example: ADAMTS9-AS1 in PRAD-1064.6.3.N.2.95.26.1.2.1); (3) the validation process relied on stringent statistical thresholds (|Meta-Z| > 3.09 or < -3.09, p < 0.001) to identify significant poorer or better prognosis, respectively. Signatures with weaker but still biologically relevant effects may not have met these thresholds in PRECOG, leading to their exclusion from the validated set. (4) Some signatures may exhibit cancer-specific prognostic value, meaning they are highly relevant in certain cancer types but not others. While PRECOG includes cancer types equivalent to TCGA, the absence of certain subtypes or including additional subtypes in PRECOG could explain why some signatures were not validated. (5) For multi-gene signatures, the median meta-Z score was computed across all genes, which may dilute the contribution of individual genes with strong prognostic effects. This aggregation approach could cause the loss of signal for signatures where only a subset of genes drives the prognostic association.

Despite these challenges, the validation of 73 signatures in PRECOG underscores their robustness and clinical relevance across independent datasets. The partial validation rate highlights the complexity of translating gene expression signatures into universally applicable prognostic tools and emphasizes the need for further refinement and context-specific validation in future studies.

### 4.8. Advanced Tools for RCD Data Analysis

Although existing resources provide valuable insights, they have limitations that our model addresses. Four comprehensive and interactive online tools are currently available to support research on RCD in cancer: RCD map [5], FerrDb [94], HAMdb [93], XDeathDB [69] and RCDdb [132]. The first appears to have inactive hyperlinks. FerrDb is dedicated to ferroptosis regulators and disease associations. It categorizes regulators into genes (drivers, suppressors, markers, unclassified) and substances (pure and mixtures like iron, erastin, and herbal extracts). These are further classified as inducers or inhibitors. FerrDb includes seven curated datasets. HAMdb is a database of autophagy modulators and their disease links, containing 796 proteins, 841 chemicals, and 132 miRNAs. It helps identify new modulators, drug candidates, and therapeutic targets through a user-friendly interface for easy searching and browsing, advancing autophagy research in cancer and other diseases. XDeathDB gathers information about a 12-optosis model that includes intrinsic apoptosis, autosis, efferocytosis, ferroptosis, immunogenic cell death, lysosomal cell death, mitotic cell death, mitochondrial permeability transition, necroptosis, parthanatos, and pyroptosis. It integrates big data for cell death gene-disease associations, gene-cell death pathway associations, pathway-cell death mode associations, and cell death-cell death associations derived from literature reviews and public databases. RCDdb features over 3,000 literature-derived annotations covering 1,850 RCD-associated genes linked to 15 RCD forms (apoptosis, pyroptosis, necroptosis, autophagy-dependent cell death, entotic cell death, NETotic cell death, parthanatos, MPT-driven necrosis, immunogenic cell death, lysosome-dependent cell death, ferroptosis, alkaliptosis, oxeiptosis, cuproptosis, and disulfidptosis). It integrates data on diseases, drugs, pathways, proteins, and gene expression and provides advanced visualization tools and three analytical modules to enable users to identify and study RCD-related features. The RCDdb is the first comprehensive, manually curated database focused on annotating and analyzing the 15 known RCD forms.

Despite their comprehensive scopes, FerrDb, HAMdb, XDeathDB and RCDdb do not index outputs by significance, making it challenging to prioritize critical associations, which can hinder effective data utilization and research prioritization. We developed the *CancerRCDShiny* web browser and the *Cancer Regulated Cell Death Data Analyst* tools to address these gaps. These new tools enhance the utility and impact of our findings, making them more accessible and actionable for researchers and clinicians. Their integrative and user-friendly design facilitates efficient extraction, analysis, and visualization of RCD data in cancer, ultimately advancing our understanding and treatment of cancer through more precise biomarkers and targeted therapies.

## 5. Shortcomings and Limitations

This study has shortcomings and limitations that should be considered when selecting impactful signatures. First, the gene inventory is an ongoing effort, which means some genes reported in various studies may have been omitted since our catalog is primarily based on the NCBI Gene database. Despite our cross-referencing, relying on a single database means the catalog may not comprehensively include all genes associated with RCD forms reported in the literature.

Second, using a stringent genome-wide significance threshold (*padj*-value < 5×10^-8) while minimizing false positives may reduce sensitivity, especially in smaller datasets or those with lower signal-to-noise ratios. Users should consider the specific context of their dataset and study design when applying this threshold. A less stringent threshold might enhance sensitivity in particular scenarios while maintaining the stringent threshold is crucial in larger datasets to control false discovery rates. We have made our source code publicly available, enabling researchers to fine-tune the significance threshold.

Third, the signatures identified in this study are related to primary tumor samples. Therefore, the impact values of these signatures in recurrent tumors, metastatic tumors, and primary blood-derived cancers were not addressed in this study. Future studies should expand the analysis to these other tumor types to provide a more comprehensive understanding of the signatures’ roles across different cancer stages and contexts.

Fourth, the corpora of PDFs comprise only free-access full-text files. This limitation may cause a biased dataset, as some relevant studies published in subscription-based journals were not included. Cross-referencing gene targets and gene components of signatures might miss critical information available in those restricted-access publications. Future research should incorporate a broader range of manually curated sources to ensure greater accuracy and depth in the findings.

Lastly, although this study identifies biomarkers with potential immunotherapeutic applications, it does not incorporate AI-driven drug discovery or molecular docking methods to identify or validate therapeutic compounds. Such approaches could refine our ability to screen for specific inhibitors or activators targeting RCD-related pathways and enhance the translational relevance of our findings. Future research should aim to integrate AI and docking-based platforms into the *CancerRCDShiny* tool to support the discovery of novel drugs targeting the multi-omic signatures identified in this study.

## 6. Strengths

The strengths of this research include a comprehensive exploration of 25 forms of RCD in cancer, integrating multi-omic data to identify signatures associated with RCD. A scoring method was developed to assess the significance of these signatures. A PDF-Ai-based strategy was also implemented to provide evidence-based support for signature involvement. The CancerRCDShiny tool facilitates data exploration and analysis, enhancing the overall effectiveness of this research.

## 7. Concluding remarks

This study introduces the multi-optosis framework as a novel, integrative approach for investigating RCD mechanisms in cancer. By incorporating 25 distinct forms of RCD, the model transcends traditional, single-pathway analyses, offering a holistic view of the intricate crosstalk between RCD pathways. This framework advances our understanding of cancer progression and treatment resistance while providing a robust platform for identifying genome-wide biomarkers and actionable therapeutic targets. Notably, the multi-optosis model lays the foundation for clinical applications, such as stratifying patients based on RCD-related phenotypes and designing therapies that target multiple RCD pathways for enhanced efficacy.

We developed a signature database enriched with a systematic nomenclature that reveals hidden correlations between multi-omic and phenotypic features. Our ranking method ensures a balanced and comprehensive assessment of signature relevance by integrating survival metrics and tumor immune infiltration profiles. This prioritization speeds up the discovery of actionable insights and supports the development of personalized therapeutic strategies to improve patient outcomes.

Practical applications of our findings are facilitated by user-friendly tools such as CancerRCDShiny and the Cancer Regulated Cell Death Data Analyst. These tools enable researchers and clinicians to explore RCD multi-omic signatures efficiently, leveraging dynamic visualization and customizable reporting capabilities to enhance data interpretation.

By addressing the complexity and heterogeneity of cancer biology, the multi-optosis framework provides a detailed understanding of RCD gene associations in cancer progression and resistance. This integrative approach paves the way for identifying candidate biomarkers and therapeutic targets, driving the development of more effective cancer treatments.

Together, the multi-optosis model and its associated tools represent a significant advancement in cancer biomarker discovery and translational research, offering invaluable resources for personalized cancer therapies and improved clinical outcomes.

## Supporting information

https://github.com/CancerRCD/Supplementary-Material

## Data Availability

All data produced in the present work are contained in the manuscript.
All source codes for these analyses are in the supporting source code repository available at GitHub URL: https://github.com/CancerRCD.

## 8. Data availability

All source codes for these analyses are in the supporting source code repository available at GitHub URL: https://github.com/CancerRCD.

## 9. Ethics statement

The study aimed to identify signatures for future development of prognostic markers in cancer genomics. The study did not involve physical or history examinations of subjects or laboratory testing for a genetic disease, as the information about the subjects’ DNA, protein, methylation, mutation, and clinical data were from public repositories.

## 11. Author contributions

ERS, HACN, ABG, EMA: Conceived and designed the experiments, conducted comprehensive computational analyses, crated the *Cancer Regulated Cell Death Data Analyst* tool, and revised the manuscript.

ABG: Performed PDF-AI-assisted NLP extraction.

ERS, HACN, EMA: Developed and validated the R codes.

ERS, HACN, EMA: Created the figures.

EMA: Drafted the manuscript and compiled the supplementary material.

HACN: Created the *CancerRCDShiny* app, created and maintain the GitHub repository RSFJ, EMA: Performed the PRECOG analysis

## 12. Acknowledgments

The authors thank Dr. Shixiang Wang (https://github.com/ShixiangWang) and Dr. Shensuo Li, (https://github.com/lishensuo), from the team of developers of the UCSCXena shiny application, for their helpful coding suggestions.

## 13. Footnotes

ERS and HACN are recipients of PIBIC/CNPq and FAPERJ undergraduate and graduate fellowships, respectively. The agencies had no role in the study design, data collection and analysis, decision to publish, or manuscript preparation.

## Web resources (Footnotes)

**BioMart**: https://www.ensembl.org/biomart/

**COSMIC**: https://cancer.sanger.ac.uk/cosmic

**DGIdb**: https://dgidb.org

**FerrDb**: http://www.zhounan.org/ferrdb/legacy/index.html

**GTEx Portal**: https://www.gtexportal.org/

**HAMdb**: http://hamdb.scbdd.com/

**KEGG**: https://www.genome.jp/kegg/

**PDF AiDrive**: https://myaidrive.com

**PRECOG:** https://precog.stanford.edu/

**R software package**: http://www.R-project.org

**RCDdb**: http://chenyclab.com/RCDdb/

**RCD map**: https://navicell.vincent-noel.fr/pages/maps_rcd.html

**UCSC Genome browser**: https://genome.ucsc.edu/

**UCSCXena**: https://xena.ucsc.edu/

**UCSCXenaShiny**: https://shiny.zhoulab.ac.cn/UCSCXenaShiny/#

**XDeathDB**: https://pcm2019.shinyapps.io/XDeathDB/

## 13 References

1. Newton, K., et al., Cell death. Cell, 2024. 187(2): p. 235–256.

2. Gong, L., et al., Regulated cell death in cancer: from pathogenesis to treatment. Chin Med J (Engl), 2023. 136(6): p. 653–665.

3. Koren, E. and Y. Fuchs, Modes of Regulated Cell Death in Cancer. Cancer Discov, 2021. 11(2): p. 245–265.

4. Galluzzi, L., et al., Molecular mechanisms of cell death: recommendations of the Nomenclature Committee on Cell Death 2018. Cell Death Differ, 2018. 25(3): p. 486–541.

5. Ravel, J.M., et al., Comprehensive Map of the Regulated Cell Death Signaling Network: A Powerful Analytical Tool for Studying Diseases. Cancers (Basel), 2020. 12(4): p. 990.

6. Peng, F., et al., Regulated cell death (RCD) in cancer: key pathways and targeted therapies. Signal Transduct Target Ther, 2022. 7(1): p. 286.

7. Elmore, S., Apoptosis: a review of programmed cell death. Toxicol Pathol, 2007. 35(4): p. 495–516.

8. Galluzzi, L., et al., Necroptosis: Mechanisms and Relevance to Disease. Annu Rev Pathol, 2017. 12(Volume 12, 2017): p. 103–130.

9. Jorgensen, I., M. Rayamajhi, and E.A. Miao, Programmed cell death as a defence against infection. Nat Rev Immunol, 2017. 17(3): p. 151–164.

10. Stockwell, B.R., et al., Ferroptosis: A Regulated Cell Death Nexus Linking Metabolism, Redox Biology, and Disease. Cell, 2017. 171(2): p. 273–285.

11. Debnath, J., N. Gammoh, and K.M. Ryan, Autophagy and autophagy-related pathways in cancer. Nat Rev Mol Cell Biol, 2023. 24(8): p. 560–575.

12. Feng, Y., et al., Cuproptosis: unveiling a new frontier in cancer biology and therapeutics. Cell Commun Signal, 2024. 22(1): p. 249.

13. Castedo, M., et al., Cell death by mitotic catastrophe: a molecular definition. Oncogene, 2004. 23(16): p. 2825–37.

14. Fatokun, A.A., V.L. Dawson, and T.M. Dawson, Parthanatos: mitochondrial-linked mechanisms and therapeutic opportunities. Br J Pharmacol, 2014. 171(8): p. 2000–16.

15. Choi, M., et al., Immunogenic cell death in cancer immunotherapy. BMB Rep, 2023. 56(5): p. 275–286.

16. Bai, L., et al., Autosis as a selective type of cell death. Front Cell Dev Biol, 2023. 11: p. 1164681.

17. Brinkmann, V., et al., Neutrophil extracellular traps kill bacteria. Science, 2004. 303(5663): p. 1532–5.

18. Zheng, P., et al., Disulfidptosis: a new target for metabolic cancer therapy. J Exp Clin Cancer Res, 2023. 42(1): p. 103.

19. Chen, F., et al., Mechanisms of alkaliptosis. Front Cell Dev Biol, 2023. 11: p. 1213995.

20. Aits, S. and M. Jaattela, Lysosomal cell death at a glance. J Cell Sci, 2013. 126(Pt 9): p. 1905–12.

21. Overholtzer, M., et al., A nonapoptotic cell death process, entosis, that occurs by cell-in-cell invasion. Cell, 2007. 131(5): p. 966–79.

22. Frisch, S.M. and H. Francis, Disruption of epithelial cell-matrix interactions induces apoptosis. J Cell Biol, 1994. 124(4): p. 619–26.

23. Holze, C., et al., Oxeiptosis, a ROS-induced caspase-independent apoptosis-like cell-death pathway. Nat Immunol, 2018. 19(2): p. 130–140.

24. Sperandio, S., I. de Belle, and D.E. Bredesen, An alternative, nonapoptotic form of programmed cell death. Proc Natl Acad Sci U S A, 2000. 97(26): p. 14376–81.

25. Campisi, J., Aging, cellular senescence, and cancer. Annu Rev Physiol, 2013. 75(Volume 75, 2013): p. 685–705.

26. Lyamzaev, K.G., D.A. Knorre, and B.V. Chernyak, Mitoptosis, Twenty Years After. Biochemistry (Mosc), 2020. 85(12): p. 1484–1498.

27. Ciesielski, H.M., et al., Erebosis, a new cell death mechanism during homeostatic turnover of gut enterocytes. PLoS Biol, 2022. 20(4): p. e3001586.

28. Qiu, H., et al., Efferocytosis: An accomplice of cancer immune escape. Biomed Pharmacother, 2023. 167: p. 115540.

29. Suh, D.H., et al., Mitochondrial permeability transition pore as a selective target for anti-cancer therapy. Front Oncol, 2013. 3: p. 41.

30. Maltese, W.A. and J.H. Overmeyer, Methuosis: nonapoptotic cell death associated with vacuolization of macropinosome and endosome compartments. Am J Pathol, 2014. 184(6): p. 1630–42.

31. Kim, E.H., S.W. Wong, and J. Martinez, Programmed Necrosis and Disease:We interrupt your regular programming to bring you necroinflammation. Cell Death Differ, 2019. 26(1): p. 25–40.

32. Liang, J.Y., et al., A Novel Ferroptosis-related Gene Signature for Overall Survival Prediction in Patients with Hepatocellular Carcinoma. Int J Biol Sci, 2020. 16(13): p. 2430–2441.

33. Zhang, Y., et al., Pyroptosis-Related Gene Signature Predicts Prognosis and Indicates Immune Microenvironment Infiltration in Glioma. Front Cell Dev Biol, 2022. 10: p. 862493.

34. Zhang, Z., et al., Cuproptosis-related gene signature stratifies lower-grade glioma patients and predicts immune characteristics. Front Genet, 2022. 13: p. 1036460.

35. Xu, L., et al., Identification of a necroptosis-related gene signature as a novel prognostic biomarker of cholangiocarcinoma. Front Immunol, 2023. 14: p. 1118816.

36. Su, W., et al., A unique regulated cell death-related classification regarding prognosis and immune landscapes in non-small cell lung cancer. Front Immunol, 2023. 14: p. 1075848.

37. Sun, X., et al., PANoptosis: Mechanisms, biology, and role in disease. Immunol Rev, 2024. 321(1): p. 246–262.

38. Zou, Y., et al., Leveraging diverse cell-death patterns to predict the prognosis and drug sensitivity of triple-negative breast cancer patients after surgery. Int J Surg, 2022. 107: p. 106936.

39. Wei, Q., et al., Molecular subtypes of lung adenocarcinoma patients for prognosis and therapeutic response prediction with machine learning on 13 programmed cell death patterns. J Cancer Res Clin Oncol, 2023. 149(13): p. 11351–11368.

40. Wang, Y. and Q. Zhang, Leveraging programmed cell death signature to predict clinical outcome and immunotherapy benefits in postoperative bladder cancer. Sci Rep, 2024. 14(1): p. 22976.

41. Samir, P., R.K.S. Malireddi, and T.D. Kanneganti, The PANoptosome: A Deadly Protein Complex Driving Pyroptosis, Apoptosis, and Necroptosis (PANoptosis). Front Cell Infect Microbiol, 2020. 10: p. 238.

42. Wang, Y. and T.D. Kanneganti, From pyroptosis, apoptosis and necroptosis to PANoptosis: A mechanistic compendium of programmed cell death pathways. Comput Struct Biotechnol J, 2021. 19: p. 4641–4657.

43. Shi, C., et al., PANoptosis: A Cell Death Characterized by Pyroptosis, Apoptosis, and Necroptosis. J Inflamm Res, 2023. 16: p. 1523–1532.

44. Zhang, Z., et al., Identification of PANoptosis-relevant subgroups to evaluate the prognosis and immune landscape of patients with liver hepatocellular carcinoma. Front Cell Dev Biol, 2023. 11: p. 1210456.

45. Zhu, J., et al., Identification of molecular subtypes based on PANoptosis-related genes and construction of a signature for predicting the prognosis and response to immunotherapy response in hepatocellular carcinoma. Front Immunol, 2023. 14: p. 1218661.

46. Han, C., et al., Machine learning reveals PANoptosis as a potential reporter and prognostic revealer of tumour microenvironment in lung adenocarcinoma. J Gene Med, 2024. 26(1): p. e3599.

47. Yu, T., et al., Identification of a PANoptosis-related Gene Signature for Predicting the Prognosis, Tumor Microenvironment and Therapy Response in Breast Cancer. J Cancer, 2024. 15(2): p. 428–443.

48. Zhao, Q., et al., PANoptosis-related long non-coding RNA signature to predict the prognosis and immune landscapes of pancreatic adenocarcinoma. Biochem Biophys Rep, 2024. 37: p. 101600.

49. Liu, Y., et al., PANoptosis-related genes function as efficient prognostic biomarkers in colon adenocarcinoma. Front Endocrinol (Lausanne), 2024. 15: p. 1344058.

50. Gao, F., et al., A PANoptosis pattern to predict prognosis and immunotherapy response in head and neck squamous cell carcinoma. Heliyon, 2024. 10(5): p. e27162.

51. Sun, F., et al., Identification of PANoptosis-related predictors for prognosis and tumor microenvironment by multiomics analysis in glioma. J Cancer, 2024. 15(9): p. 2486–2504.

52. Tang, L., et al., Machine Learning-Based Integrated Analysis of PANoptosis Patterns in Acute Myeloid Leukemia Reveals a Signature Predicting Survival and Immunotherapy. Int J Clin Pract, 2024. 2024: p. 5113990.

53. Xie, D., et al., Identification of PANoptosis-related genes as prognostic indicators of thyroid cancer. Heliyon, 2024. 10(11): p. e31707.

54. Zhong, L., et al., Molecular Subtypes Based on PANoptosis Genes and Characteristics of Immune Infiltration in Cutaneous Melanoma. Cell Mol Biol (Noisy-le-grand), 2023. 69(8): p. 1–8.

55. Zhou, Y., et al., Tumor biomarkers for diagnosis, prognosis and targeted therapy. Signal Transduct Target Ther, 2024. 9(1): p. 132.

56. Souers, A.J., et al., ABT-199, a potent and selective BCL-2 inhibitor, achieves antitumor activity while sparing platelets. Nat Med, 2013. 19(2): p. 202–8.

57. Roberts, A.W., et al., Targeting BCL2 with Venetoclax in Relapsed Chronic Lymphocytic Leukemia. N Engl J Med, 2016. 374(4): p. 311–22.

58. DiNardo, C.D., et al., Venetoclax combined with decitabine or azacitidine in treatment-naive, elderly patients with acute myeloid leukemia. Blood, 2019. 133(1): p. 7–17.

59. Hanahan, D. and R.A. Weinberg, Hallmarks of cancer: the next generation. Cell, 2011. 144(5): p. 646–74.

60. Castelli, V., et al., The Great Escape: The Power of Cancer Stem Cells to Evade Programmed Cell Death. Cancers (Basel), 2021. 13(2).

61. Aubrey, B.J., et al., How does p53 induce apoptosis and how does this relate to p53-mediated tumour suppression? Cell Death Differ, 2018. 25(1): p. 104–113.

62. Su, Y., et al., Current insights into the regulation of programmed cell death by TP53 mutation in cancer. Front Oncol, 2022. 12: p. 1023427.

63. Ghavami, S., et al., Apoptosis and cancer: mutations within caspase genes. J Med Genet, 2009. 46(8): p. 497–510.

64. Neophytou, C.M., et al., Apoptosis Deregulation and the Development of Cancer Multi-Drug Resistance. Cancers (Basel), 2021. 13(17).

65. Wei, Y., et al., Robust analysis of a novel PANoptosis-related prognostic gene signature model for hepatocellular carcinoma immune infiltration and therapeutic response. Sci Rep, 2023. 13(1): p. 14519.

66. Chen, G., et al., Construction of a machine learning-based artificial neural network for discriminating PANoptosis related subgroups to predict prognosis in low-grade gliomas. Sci Rep, 2022. 12(1): p. 22119.

67. Wang, X., et al., PANoptosis-based molecular clustering and prognostic signature predicts patient survival and immune landscape in colon cancer. Front Genet, 2022. 13: p. 955355.

68. Pan, B., et al., Non-Canonical Programmed Cell Death in Colon Cancer. Cancers (Basel), 2022. 14(14).

69. Gadepalli, V.S., et al., XDeathDB: a visualization platform for cell death molecular interactions. Cell Death Dis, 2021. 12(12): p. 1156.

70. Ye, Z., Y. Jiang, and J. Wu, A novel necroptosis-associated miRNA signature predicting prognosis of endometrial cancer and correlated with immune infiltration. Taiwan J Obstet Gynecol, 2023. 62(2): p. 291–298.

71. Pan, S., et al., Comprehensive Analysis of Programmed Cell Death Signature in the Prognosis, Tumor Microenvironment and Drug Sensitivity in Lung Adenocarcinoma. Front Genet, 2022. 13: p. 900159.

72. Wu, Z., et al., Identification and Validation of Ferroptosis-Related LncRNA Signatures as a Novel Prognostic Model for Colon Cancer. Front Immunol, 2021. 12: p. 783362.

73. Cancer Genome Atlas Research, N., et al., The Cancer Genome Atlas Pan-Cancer analysis project. Nat Genet, 2013. 45(10): p. 1113–20.

74. Goldman, M.J., et al., Visualizing and interpreting cancer genomics data via the Xena platform. Nat Biotechnol, 2020. 38(6): p. 675–678.

75. Wang, S., et al., UCSCXenaShiny: an R/CRAN package for interactive analysis of UCSC Xena data. Bioinformatics, 2022. 38(2): p. 527–529.

76. Li, S., et al., Facilitating integrative and personalized oncology omics analysis with UCSCXenaShiny. Commun Biol, 2024. 7(1): p. 1200.

77. Brown, G.R., et al., Gene: a gene-centered information resource at NCBI. Nucleic Acids Res, 2015. 43(Database issue): p. D36–42.

78. Consortium, G.T., The Genotype-Tissue Expression (GTEx) project. Nat Genet, 2013. 45(6): p. 580–5.

79. Akbani, R., et al., A pan-cancer proteomic perspective on The Cancer Genome Atlas. Nat Commun, 2014. 5: p. 3887.

80. Sanjai, C., et al., A comprehensive review on anticancer evaluation techniques. Bioorg Chem, 2024. 142: p. 106973.

81. Ren, H., et al., A cuproptosis-related gene expression signature predicting clinical prognosis and immune responses in intrahepatic cholangiocarcinoma detected by single-cell RNA sequence analysis. Cancer Cell Int, 2024. 24(1): p. 92.

82. Royle, K.L., et al., How is overall survival assessed in randomised clinical trials in cancer and are subsequent treatment lines considered? A systematic review. Trials, 2023. 24(1): p. 708.

83. Korn, R.L. and J.J. Crowley, Overview: progression-free survival as an endpoint in clinical trials with solid tumors. Clin Cancer Res, 2013. 19(10): p. 2607–12.

84. Newman, A.M., et al., Robust enumeration of cell subsets from tissue expression profiles. Nat Methods, 2015. 12(5): p. 453–7.

85. Aran, D., Z. Hu, and A.J. Butte, xCell: digitally portraying the tissue cellular heterogeneity landscape. Genome Biol, 2017. 18(1): p. 220.

86. Galon, J. and D. Bruni, Approaches to treat immune hot, altered and cold tumours with combination immunotherapies. Nat Rev Drug Discov, 2019. 18(3): p. 197–218.

87. Wang, L., et al., Hot and cold tumors: Immunological features and the therapeutic strategies. MedComm (2020), 2023. 4(5): p. e343.

88. Wang, M., et al., Therapeutic strategies to remodel immunologically cold tumors. Clin Transl Immunology, 2020. 9(12): p. e1226.

89. Sahu, A., et al., In vivo tumor immune microenvironment phenotypes correlate with inflammation and vasculature to predict immunotherapy response. Nat Commun, 2022. 13(1): p. 5312.

90. Liu, J., et al., An Integrated TCGA Pan-Cancer Clinical Data Resource to Drive High-Quality Survival Outcome Analytics. Cell, 2018. 173(2): p. 400–416 e11.

91. Cannon, M., et al., DGIdb 5.0: rebuilding the drug-gene interaction database for precision medicine and drug discovery platforms. Nucleic Acids Res, 2024. 52(D1): p. D1227–D1235.

92. Gentles, A.J., et al., The prognostic landscape of genes and infiltrating immune cells across human cancers. Nature Medicine, 2015. 21(8): p. 938–945.

93. Wang, N.N., et al., HAMdb: a database of human autophagy modulators with specific pathway and disease information. J Cheminform, 2018. 10(1): p. 34.

94. Zhou, N. and J. Bao, FerrDb: a manually curated resource for regulators and markers of ferroptosis and ferroptosis-disease associations. Database (Oxford), 2020. 2020.

95. Sondka, Z., et al., The COSMIC Cancer Gene Census: describing genetic dysfunction across all human cancers. Nat Rev Cancer, 2018. 18(11): p. 696–705.

96. Kinnersley, B., et al., Analysis of 10,478 cancer genomes identifies candidate driver genes and opportunities for precision oncology. Nat Genet, 2024. 56(9): p. 1868–1877.

97. Bao, Z.S., et al., Prognostic value of a nine-gene signature in glioma patients based on mRNA expression profiling. CNS Neurosci Ther, 2014. 20(2): p. 112–8.

98. Cai, Y., et al., Combined Use of Three Machine Learning Modeling Methods to Develop a Ten-Gene Signature for the Diagnosis of Ventilator-Associated Pneumonia. Med Sci Monit, 2020. 26: p. e919035.

99. Chen, H., et al., Identification of an autophagy-related gene signature for survival prediction in patients with cervical cancer. J Ovarian Res, 2020. 13(1): p. 131.

100. Chang, K., C. Yuan, and X. Liu, Ferroptosis-Related Gene Signature Accurately Predicts Survival Outcomes in Patients With Clear-Cell Renal Cell Carcinoma. Front Oncol, 2021. 11: p. 649347.

101. Modarres, P., et al., Meta-analysis of gene signatures and key pathways indicates suppression of JNK pathway as a regulator of chemo-resistance in AML. Sci Rep, 2021. 11(1): p. 12485.

102. Wan, R.J., et al., Ferroptosis-related gene signature predicts prognosis and immunotherapy in glioma. CNS Neurosci Ther, 2021. 27(8): p. 973–986.

103. Bian, Z., R. Fan, and L. Xie, A Novel Cuproptosis-Related Prognostic Gene Signature and Validation of Differential Expression in Clear Cell Renal Cell Carcinoma. Genes (Basel), 2022. 13(5): p. 851.

104. Li, C., et al., A seven-autophagy-related gene signature for predicting the prognosis of differentiated thyroid carcinoma. World J Surg Oncol, 2022. 20(1): p. 129.

105. Yao, X., et al., Exploration and validation of a novel ferroptosis-related gene signature predicting the prognosis of intrahepatic cholangiocarcinoma. Acta Biochim Biophys Sin (Shanghai), 2022. 54(9): p. 1376–1385.

106. Chen, L., et al., An Immunogenic Cell Death-Related Gene Signature Reflects Immune Landscape and Predicts Prognosis in Melanoma Independently of BRAF V600E Status. Biomed Res Int, 2023. 2023: p. 1189022.

107. Li, J., et al., The anoikis-related gene signature predicts survival and correlates with immune infiltration in osteosarcoma. Aging (Albany NY), 2024. 16(1): p. 665–684.

108. Zhou, X., et al., Identification of a necroptosis-related gene signature for making clinical predictions of the survival of patients with lung adenocarcinoma. PeerJ, 2024. 12: p. e16616.

109. Clemente-Perivan, S.I., et al., Role of Oct3/4 in Cervical Cancer Tumorigenesis. Front Oncol, 2020. 10: p. 247.

110. Chiou, S.H., et al., Coexpression of Oct4 and Nanog enhances malignancy in lung adenocarcinoma by inducing cancer stem cell-like properties and epithelial-mesenchymal transdifferentiation. Cancer Res, 2010. 70(24): p. 10433–44.

111. Wu, Y.C., et al., Chemotherapeutic sensitivity of testicular germ cell tumors under hypoxic conditions is negatively regulated by SENP1-controlled sumoylation of OCT4. Cancer Res, 2012. 72(19): p. 4963–73.

112. Gutekunst, M., et al., Cisplatin hypersensitivity of testicular germ cell tumors is determined by high constitutive Noxa levels mediated by Oct-4. Cancer Res, 2013. 73(5): p. 1460–9.

113. Upadhyay, V.A., et al., Putative stemness markers octamer-binding transcription factor 4, sex-determining region Y-box 2, and NANOG in non-small cell lung carcinoma: A clinicopathological association. J Cancer Res Ther, 2020. 16(4): p. 804–810.

114. Mehrzad, N., S. Irani, and M. Karimipoor, Expression of Sox2, Oct4, and Nanog in Human Lung Cancer Non-small-cell. Immunoregulation, 2022. 5(1): p. 49–58.

115. Fang, X., T. Zhang, and Z. Chen, Solute Carrier Family 7 Member 11 (SLC7A11) is a Potential Prognostic Biomarker in Uterine Corpus Endometrial Carcinoma. Int J Gen Med, 2023. 16: p. 481–497.

116. von Eyben, F.E., et al., Epigenetic Regulation of Driver Genes in Testicular Tumorigenesis. Int J Mol Sci, 2023. 24(4).

117. Zhu, H. and S. Xu, SOX4 inhibits ferroptosis and promotes proliferation of endometrial cancer cells via the p53/SLC7A11 signaling. J Obstet Gynaecol Res, 2024. 50(11): p. 2093–2106.

118. Blagih, J., M.D. Buck, and K.H. Vousden, p53, cancer and the immune response. J Cell Sci, 2020. 133(5).

119. Berx, G. and F. Van Roy, The E-cadherin/catenin complex: an important gatekeeper in breast cancer tumorigenesis and malignant progression. Breast Cancer Res, 2001. 3(5): p. 289–93.

120. Liu, H., et al., SPHK1 (sphingosine kinase 1) induces epithelial-mesenchymal transition by promoting the autophagy-linked lysosomal degradation of CDH1/E-cadherin in hepatoma cells. Autophagy, 2017. 13(5): p. 900–913.

121. Liu, Y., et al., Ferroptosis in Low-Grade Glioma: A New Marker for Diagnosis and Prognosis. Med Sci Monit, 2020. 26: p. e921947.

122. Zhang, X., et al., Neoantigen DNA vaccines are safe, feasible, and induce neoantigen-specific immune responses in triple-negative breast cancer patients. Genome Med, 2024. 16(1): p. 131.

123. Wang, D.R., X.L. Wu, and Y.L. Sun, Therapeutic targets and biomarkers of tumor immunotherapy: response versus non-response. Signal Transduct Target Ther, 2022. 7(1): p. 331.

124. Dannenfelser, R., et al., Discriminatory Power of Combinatorial Antigen Recognition in Cancer T Cell Therapies. Cell Syst, 2020. 11(3): p. 215–228 e5.

125. Topalian, S.L., et al., Safety, activity, and immune correlates of anti-PD-1 antibody in cancer. N Engl J Med, 2012. 366(26): p. 2443–54.

126. Zeng, Y.F., et al., The efficacy and safety of anti-PD-1/PD-L1 in treatment of glioma: a single-arm meta-analysis. Front Immunol, 2023. 14: p. 1168244.

127. Chen, R.Q., et al., The Prognostic and Therapeutic Value of PD-L1 in Glioma. Front Pharmacol, 2018. 9: p. 1503.

128. Feng, M., et al., PD-1/PD-L1 and immunotherapy for pancreatic cancer. Cancer Lett, 2017. 407: p. 57–65.

129. Goyette, M.A. and J.F. Cote, AXL Receptor Tyrosine Kinase as a Promising Therapeutic Target Directing Multiple Aspects of Cancer Progression and Metastasis. Cancers (Basel), 2022. 14(3).

130. Leconet, W., et al., Preclinical validation of AXL receptor as a target for antibody-based pancreatic cancer immunotherapy. Oncogene, 2014. 33(47): p. 5405–14.

131. Pidkovka, N. and A. Belkhiri, Altered expression of AXL receptor tyrosine kinase in gastrointestinal cancers: a promising therapeutic target. Front Oncol, 2023. 13: p. 1079041.

132. Wang, X., et al., RCDdb: A manually curated database and analysis platform for regulated cell death. Comput Struct Biotechnol J, 2024. 23: p. 3211–3221.

