## Supplementary material for "Integrated Multi-Optosis Model for Pan-Cancer Candidate Biomarker and Therapy Target Discovery": https://github.com/CancerRCD/Supplementary-Material

**CROSS-REFERENCING PDFs CORPUS A**

The PDF corpus was compiled by extracting PMID numbers from PubMed using both programmatic (Supplementary source code URL: https://github.com/CancerRCD.) and manual methods. Boolean queries were constructed to combine each of the 25 forms of regulated cell death (RCD) with terms "cancer," "tumor," "gene," "immunotherapy," "target," and "signature." The queries were restricted to studies on Homo sapiens and limited to free-access full-text articles.

For RCD forms that did not yield direct hits, relevant PMIDs were manually selected using specific terms for each RCD type along with "gene" or "signature."
The PMIDs were downloaded in a format compatible with EndNote (RIS format), and these citations were subsequently used to download the corresponding PDF files.
The content of these files was manually inspected to exclude irrelevant documents and any retracted publications. The final corpus consists of 6,603 document files.

Accessible from: https://myaidrive.com/3wJiT5RAvo34apuQZM37xK/89wn.folder.pdf (It requires sign-in account from myaidrive.com and OpenAi ChatGTP4o).

**CROSS-REFERENCING PDFs CORPUS B**

Accessible from: https://myaidrive.com/sNbwNifn9zL5XWyGDFXQG4/3iKL.folder.pdf (It requires account from myaidrive.com and OpenAi ChatGTP4o).

**CLASSIFICATION OF SIGNATURES ACCORDING TO THE TUMOR MICROENVIRONMENT**
**Code source file: cell_classification**
**Note: all source codes are in supplementary source files**

**METHODOLOGY 1**

For each signature, correlations with the presence of tumor microenvironment cells (See Dataset S1G) were calculated. The direction and magnitude of the correlations were considered:

- **Direct Correlation:** A positive correlation with overexpression (or negative with underexpression) indicates a greater presence of cells.
- **Inverse Correlation:** A negative correlation with overexpression (or positive with underexpression) indicates a lesser presence of cells.

**Scoring System**

Each correlation was added or subtracted based on the signature's expression and the cell classification:

- For signatures with overexpression, positive correlations were added and negative correlations were subtracted.
- For signatures with underexpression, negative correlations were added and positive correlations were subtracted.
- For signatures unchanged, positive correlations indicated the presence and negative correlations indicated the absence of cells.

**Determination of Final Classification**

Signatures were classified based on the total sum of adjusted correlations for each category:

- **Anti-tumoral:** Signatures with the highest total sum of adjusted correlations for anti-tumor cells.
- **Pro-tumoral:** Signatures with the highest total sum of adjusted correlations for pro-tumor cells.
- **Dual:** Signatures with the highest total sum of adjusted correlations for dual cells.

**Scoring Details**
Each signature was accompanied by a detailed scoring, including specific cells and their correlation values.

**CLASSIFICATION OF SIGNATURES ACCORDING TO THE TUMOR IMMUNE PROFILES CONCEPT FOR CLASSIFICATION OF TUMORS AS HOT, COLD, OR VARIABLE**
The classification of tumors as "hot," "variable," and "cold," introduced by Camus et al., is based on the interaction between tumor evasion and immunological coordination.

**Hot Tumors:**
- **Characteristics:** High infiltration of T cells, elevated expression of PD-L1, genomic instability, and pre-existing antitumor immune responses.
- **Microenvironment:** High inflammatory signature (M1) and a significant presence of cytotoxic cells such as NK cells, CD8 T cells, and dendritic cells.

**Cold Tumors:**
- **Characteristics:** Low infiltration of T cells and predominance of immunosuppressive pathways.
- **Microenvironment:** Low inflammatory signature (M2) and absence of CD8 T cells and NK cells.

**Variable Tumors:**
- **Characteristics:** Intermediate state between hot and cold tumors.

The difference between these types of tumors is primarily determined by the presence and activity of cytotoxic T cells within the tumor, along with specific immunological and molecular factors.
Currently, the terms "hot" and "cold" are used to denote tumors infiltrated by lymphocytes. Tumor-infiltrating lymphocytes (TILs) are immune cells isolated from tumor tissue that contain both positive regulatory immune cells, such as dendritic cells (DCs), CD8 T cells, and NK cells, and negative regulatory immune cells, such as tumor-associated macrophages (TAMs), regulatory T cells (Tregs), myeloid-derived suppressor cells (MDSCs), and tumor-associated neutrophils (TANs).

This concept has led to the development and implementation of the Immunoscore, a robust and standardized scoring system based on the quantification of two lymphocyte populations (CD3 and CD8).

**Immunoscore Classifications:**
- **High Immunoscore Tumors:** High degree of T cell and cytotoxic T cell infiltration.
- **Altered and Immunosuppressed Tumors:** Low presence of T cell and cytotoxic T cell infiltrates, along with the presence of immunosuppressive cells such as myeloid-derived suppressor cells and regulatory T cells.

- **Cold Tumors:** Absence of T cells within the tumor and at its margins, resulting in a low Immunoscore.

This classification is fundamental for understanding the response to immunotherapy and developing new treatment strategies for different cancer types. The efficacy of immune checkpoint inhibitors (ICIs) varies among these types, being more effective in hot tumors. Converting cold or variable tumors into hot ones is a promising area of clinical research aimed at improving therapeutic options for malignant cancers.

**Method for Bioinformatics Analysis**
To determine whether a tumor is hot, cold, or altered/immunosuppressed, analyze the types and proportions of immune cells present, following the characteristics of each tumor type:

**Hot Tumors:**
- **High Infiltration of T and Cytotoxic Cells:** High proportions of CD8+ T cells, NK cells, and activated dendritic cells.
- **High Inflammatory Signature (M1):** Significant presence of M1 macrophages.
- **Other Indicators:** High levels of PD-L1, markers of genomic instability, and pre-existing immune response.

**Cold Tumors:**
- **Low Infiltration of T and Cytotoxic Cells:** Low proportions of CD8+ T cells, NK cells, and dendritic cells.
- **Low Inflammatory Signature (M2):** Predominance of M2 macrophages.
- **Other Indicators:** Low levels of PD-L1 and high activity of immunosuppressive pathways.

**Altered/Immunosuppressed Tumors:**
- **Intermediate Infiltration of T and Cytotoxic Cells:** Moderate levels of CD8+ T cells and NK cells.
- **Presence of Immunosuppressive Cells:** Significant proportions of M2 macrophages, regulatory T cells (Tregs), and myeloid-derived suppressor cells (MDSCs).
- **Other Indicators:** Signs of activation of immunosuppressive pathways.

**Final Classification**
Classify the tumor according to how closely the results match the typical characteristics of each type.

**Example Analysis:** Assume a CIBERSORT analysis reveals the following results for a tumor:
- **CD8+ T Cells:** High proportion
- **NK Cells:** High proportion
- **M1 Macrophages:** High proportion
- **M2 Macrophages:** Low proportion
- **Regulatory T Cells:** Low proportion

Based on these results, the tumor would likely be classified as "hot."
Conversely, if the results showed:
- **CD8+ T Cells:** Low proportion
- **NK Cells:** Low proportion

- **M2 Macrophages:** High proportion
- **Regulatory T Cells:** High proportion

This tumor would likely be classified as "cold."

**METHODOLOGY 2**

For each signature, correlations with the presence of tumor immunophenotype (See Dataset S1H) were calculated. The direction and magnitude of the correlations were considered:

**Hot Tumors:**

- **Gene Overexpression:** High positive correlation with CD8+ T cells, activated NK cells, and M1 macrophages (rho >= Q3). Low or intermediate correlation with M2 macrophages and Tregs (rho between Q1 and Q3).
- **Gene Underexpression:** High negative correlation with CD8+ T cells, activated NK cells, and M1 macrophages (rho <= Q1). Low or intermediate correlation with M2 macrophages and Tregs (rho between Q1 and Q3).
- **Unchanged Gene Expression:** High positive correlation (rho >= Q3).

**Cold Tumors:**

- **Gene Overexpression:** High negative correlation with CD8+ T cells, activated NK cells, and M1 macrophages (rho <= Q1).
- **Gene Underexpression:** High positive correlation with CD8+ T cells, activated NK cells, and M1 macrophages (rho >= Q3).
- **Unchanged Gene Expression:** High negative correlation (rho <= Q1).

**Variable Tumors:**

- **Overexpression:** Intermediate correlation with CD8+ T cells, activated NK cells, and M1 macrophages (rho between Q1 and Q3). Intermediate correlation with M2 macrophages and Tregs (rho between Q2 and Q3).
- **Underexpression:** Intermediate correlation with CD8+ T cells, activated NK cells, and M1 macrophages (rho between Q1 and Q3). Intermediate correlation with M2 macrophages and Tregs (rho between Q2 and Q3).
- **Unchanged:** Intermediate correlation (rho between Q1 and Q3).

**Weights for Immune Cells:** Weights are assigned to different types of immune cells, with CD8+ T cells and NK cells receiving higher weights due to their critical importance in hot tumors.

**Final Classification:** Final classifications are determined based on the weighted scores:

- If the "Hot" score is greater than "Cold" and "Variable," the tumor is classified as "Hot."
- If the "Cold" score is greater than "Hot" and "Variable," the tumor is classified as "Cold."
- If the "Variable" score is greater, the tumor is classified as "Variable."

**Tie Resolution:**

- Tie between "Hot" and "Cold": Classified as "Variable."
- Tie between "Hot" and "Variable": Classified as "Hot."
- Tie between "Cold" and "Variable": Classified as "Cold."

### METHODOLOGY 3
### MULTI-OMIC SIGNATURE RANK METHOD

The Multi-omic Signature Rank Method was developed to evaluate each biomarker signature's clinical utility, specifically within the Cancer Multi-optosis and Multi-omic model. This method systematically ranks signatures by integrating genotypic, phenotypic, and cell death-related features that are predictive of patient outcomes and potential therapeutic responses. Each component within a signature is assigned an integer rank reflecting its relevance, as determined by established biological and clinical insights, ensuring a rigorous, reproducible approach to multi-omic data assessment.

The rank method assigns integer values to each feature category across multiple dimensions, which are subsequently summed to derive a final ranking score for each signature. A higher cumulative rank correlates with greater clinical significance, prioritizing signatures that may contribute valuable information for patient care.

#### Ranking Structure

1. **Genotypic Feature Ranking**: Genotypic features are prioritized by their direct biological impact on cellular functions and cancer progression. This includes:
    - **Protein Expression (7 points)**: The highest priority, as it represents the immediate functional output of genetic information.
    - **Mutations (6 points)**: Key drivers of oncogenesis through alterations in the DNA sequence.
    - **Copy Number Variations (CNVs, 5 points)**: Influence gene dosage without sequence alterations.
    - **miRNA Expression (4 points)**: Post-transcriptional regulators impacting multiple gene targets.
    - **Transcript and mRNA Expression (3 and 2 points)**: Indirect functional markers, providing insights at earlier transcriptional stages.
    - **CpG Methylation (1 point)**: Reflects epigenetic regulation with an indirect influence on gene expression.
2. **Phenotypic Feature Ranking**: Phenotypic features, ranked by their association with treatment response, include:
    - **Tumor Mutational Burden (TMB, 3 points)**: Highly correlated with immunotherapy efficacy.
    - **Microsatellite Instability (MSI, 2 points)**: Linked with genetic instability affecting treatment response.
    - **Tumor Stemness (TSM, 1 point)**: Related to aggressive tumor behavior but with a less direct impact on treatment.
3. **Spearman Correlation Sign**:
    - Positive correlations (P) are ranked higher (2 points), reflecting direct associations, while negative correlations (N) are assigned a lower rank (1 point).
4. **Tumor vs. Non-Tumor Expression (TNC)**:

  - Overexpression and underexpression (2 points) in tumor versus normal tissues are indicative of significant alterations in gene expression patterns, with unchanged expression ranked at 0.

5. **Hazard Risk Code (HRC)** and **Survival Metric Code (SMC)**:
   - Both codes utilize a structured alphanumeric system based on four survival metrics: DSS, DFI, PFI, and OS. The ranks range from 0 to 125 based on hazard values, with protective, neutral, or risky classifications tailored per genotype type.
   - SMC ranks are genotype-dependent, integrating categorical data with survival implications.
6. **Tumor Microenvironment (TMC) and Tumor Lymphocyte Infiltrate Code (TIC)**:
   - TMC values indicate immune characteristics within the tumor microenvironment: anti-tumoral (7), dual (4), pro-tumoral (1), or no significant data (0).
   - TIC values reflect immune cell infiltration levels, ranked as hot (7), variable (4), cold (1), or no significant data (0).
7. **Regulated Cell Death (RCD) Count**:
   - The RCD code ranks signatures by the number and type of regulated cell death processes involved. The more RCD forms engaged, the higher the potential relevance of the signature to cellular processes tied to cancer progression and treatment response.

#### METHODOLOGY 4

#### PDF-Ai-ASSISTED EVIDENCE OF INVOLVEMENT OF SIGNATURE MEMBERS IN THE MULTI-OPTOSIS MODEL

The integration of PDF-Ai tools into large-scale data extraction provides a robust solution for the analysis of complex, multidimensional datasets in biomedical research. In this context, PDF-Ai was deployed to facilitate an in-depth exploration of signature members involved in various regulated cell death (RCD) pathways, immunotherapy targets, and gene-specific roles in cancer-related mechanisms. This approach enabled an exhaustive extraction of relevant information directly from scientific documents, essential to construct the "Multi-Optosis model".

##### Prompting Strategy

The analysis consisted of multiple strategic steps, each designed to retrieve specific information pertinent to different aspects of RCD processes, gene involvement in immunotherapy, and comprehensive gene-pathway associations. Below is an outline of the primary prompting approaches utilized, including specific tasks for each subject area analyze

##### 1. RCD Process Analysis for Cancer Contextualization

This phase explored various RCD processes within the context of cancer. Utilizing a weight-based ranking system, this approach evaluated each RCD form based on six weighted criteria, such as extent of research, direct involvement in therapy, and clinical relevance. An additional layer of categorization sorted each process into programmed, non-programmed, or inflammatory cell death, establishing a foundation for the Multi-Optosis Model's conceptual framework.

**Prompt for searching RCD processes in corpus using PDFAi**

1. Go to the folder: https://myaidrive.com/3wJiT5RAvo34apuQZM37xK/89wn.folder.pdf

2. Find the RCDs processes in the PDFs: Alkaliptosis, Anoikis, Apoptosis, Autophagy, Autosis, Cellular senescence, Cuproptosis, Disulfidptosis, Efferocytosis, Entosis, Erebosis, Ferroptosis, Immunogenic cell death, Lysosome-dependent cell death, Methuosis, Mitochondrial permeability transition, Mitoptosis, Mitotic catastrophe, Necroptosis, Necrosis, NETosis, Oxeiptosis, Paraptosis, Parthanatos and Pyroptosis

3. Make a comprehensive table containing: detailed explanation about the occurrence of overlaps between the processes; description of the main findings related do cancer in the RCDs; ranking of the processes from highest to lowest related to their importance in cancer; criteria used for the ranking; categorize the PCD forms into either Programmed Cell Death, Non-Programmed Cell Death, or Inflammatory Cell Death, reference

4. For each process, assign a weight-based ranking method as described: 1. Extent of Research and Understanding (Weight 1): 25% 2. Direct Involvement in Cancer Therapy (Weight 2): 25% 3. Impact on Cancer Progression and Suppression (Weight 3): 20% 4. Emerging Therapeutic Potential (Weight 4): 15% 5. Biological Significance (Weight 5): 10% 6. Clinical Relevance (Weight 6): 5% These weights can be adjusted based on the specific priorities or insights from experts in the field. Each criterion will be scored on a scale of 1 to 10 for each process. The overall score ( S ) for each process can be calculated using the following equation: \[ S = W_1 \times C_1 + W_2 \times C_2 + W_3 \times C_3 + W_4 \times C_4 + W_5 \times C_5 + W_6 \times C_6 \]; add the criteria and final score to the output table in separated columns; the first column must contain the rank order 1, 2, 3, 4;

5. Add a column with the year of discovery for each process

6. Add a column called "Operational Definition by Lorenzo Galluzzi et al 2018", look up for these definitions in this article, put ND for those processes not described by them

7. For the RCDs not described by Galluzzi et al (2018) [1] and marked as ND, search for "Operational Definition" in the remaining documents, create a new column.

8. Merge the two columns with the Operational Definition.

The generated output enabled a structured comparison of the RCDs, highlighting key processes likely to impact therapeutic outcomes in cancer treatment.

**2. Immunotherapy Genes Target Analysis**

Focusing on genes involved in immunotherapy, the second approach aimed to identify specific gene symbols linked to immunotherapeutic roles. Genes were grouped in sets of ten to manage data flow effectively. The resulting table resumes the essential attributes such as clinical trial relevance, ranking, and overall context. The extraction of data on immunotherapy was tailored to not only capture the therapeutic relevance of each gene but also to systematically rank genes based on their level of investigation and applicability in clinical settings.

**Prompts for analysis of genes involved with immunotherapy using PDFAi**

**Step 1.**

a. Count the number of gene symbols and print the number of genes identified in the uploaded file “Gene_symbols_immynoterapy”.

b. Divide the specified genes into precise batches of ten genes

c. Name the batches as “batch_001”, “batch_002”, etc.

**Step 2**.

Access the folder at https://myaidrive.com/sNbwNifn9zL5XWyGDFXQG4/3iKL.folder.pdf in the folder.

**Step 3.**

Search for each gene symbol of each batch and create a comprehensive table with the following information’s separated in columns:

1. Name of the target; 2. Name of the gene; 3. Description of the target or the gene; 4. Role in the immunotherapy; 5. Evidence for immunotherapy; 6. Clinical trial evidence; 7. Pre-Clinical Trial; 8. Rank based on the trials; 9. Consistent context; 10. Reference with page number and hyperlink for each gene.

**Step 4.**

Save the output for each batch in an Excel file.

**3. Gene Analysis in PDFAi**

The analysis starts with the identification and batch organization of gene symbols, drawn from a file named “Gene_symbols_top_signatures” for subsequent processing. Following this, the genes were systematically divided into smaller, manageable sets (e.g., batches of five genes), facilitating detailed analysis and preventing information overload.

**Prompts for gene analysis in PDFAi**

**Strategy 1.**

Step 1:

a. Count the number of gene symbols and print the number of genes identified in the uploaded file “Gene_symbols_top_signatures”.

b. Divide the specified genes into precise batches of five genes each to ensure a detailed and accurate analysis within processing capabilities.

c. Name the batches as “batch_001”, “batch_002”, etc.

Step 2: Access the folder at https://myaidrive.com/3wJiT5RAvo34apuQZM37xK/89wn.folder.pdf

a. Sequentially analyze each gene for every batch, automatically proceeding from one gene to the next.

b. The Gene-Pathway Summary Table must include the following columns:

i. Gene Name

ii. Description (include only the complete form of the gene's abbreviation)

iii. Pathway Categorization (limit the search to categories such as Alkaliptosis, Anoikis, Apoptosis, Autophagy, Autosis, Cellular senescence, Cuproptosis, Disulfidptosis, Efferocytosis, Entosis, Erebosis, Ferroptosis, Immunogenic cell death, Lysosome-dependent cell death, Methuosis, Mitochondrial permeability transition, Mitoptosis, Mitotic catastrophe, Necroptosis, Necrosis, NETosis, Oxeiptosis, Paraptosis, Parthanatos and Pyroptosis)

iv. Context and Findings: Synthesize information directly obtained or inferred from the specified PDF documents.

v. Document(s) Title(s): The title of the PDF document(s) from which the information was extracted, including the page index.

vi. Hyperlinked URL with Specific Page Reference

Step 3: Ensure all genes are included in the output, even if no relevant information is extracted.

Step 4: Use the specification for Pathway Categorization: The categorization should strictly follow the terminologies used in the documents, or broader categorization can be applied based on a comprehensive understanding when direct mentions are unavailable.

Step 4: Compliance with Criteria for Adequate Information: Each gene must meet the following checklist to consider its dataset complete:

a. Confirmation of the gene’s role in at least one specified Regulated cell death pathway.

b. At least one piece of evidence supporting the gene's involvement in the pathway (e.g., experimental result, literature review).

c. Identify any contradictory information or unresolved questions regarding the gene's function in the pathway.

Step 5. For each batch analysis, please create a R script that will allow saving the output data as an Excel file. Each script for every batch should append the output to the previous Excel file.

**Strategy 2.**

Prompt for analysis

Task 1: Count the total number of valid PDF files in the folder located at https://myaidrive.com/3wJiT5RAvo34apuQZM37xK/89wn.folder.pdf.

Task 2. Title: Extraction of Gene Roles in Anti- and Pro-RCD Mechanisms in Cancer

Description: You have a list of gene symbols in the uploaded file named “Gene_symbols_top_signatures”. Your goal is to extract direct evidence from the provided PDFs regarding whether these gene symbols are implicated in anti-RCD or pro-RCD mechanisms in different cancer types. This involves identifying mentions of these genes and their roles in the specified processes from the scientific literature present in the PDF folder.

Steps to Approach the Task: 2.

1. Identify Relevant Documents:

Search the PDF folder for any documents mentioning the provided gene symbols. Focus the search on keywords related to the following RCD mechanisms (anti and pro): Alkaliptosis, Anoikis, Apoptosis, Autophagy, Autosis, Cellular senescence, Cuproptosis, Disulfidptosis, Efferocytosis, Entosis, Erebosis, Ferroptosis, Immunogenic cell death, Lysosome-dependent cell death, Methuosis, Mitochondrial permeability transition, Mitoptosis, Mitotic catastrophe, Necroptosis, Necrosis, NETosis, Oxeiptosis, Paraptosis, Parthanatos and Pyroptosis.

2. Extract Direct Evidence:

a) Within the identified documents, locate specific sections, sentences, or paragraphs that discuss the roles of these genes in anti-RCD or pro-RCD mechanisms.

b) Extract direct quotes that specifically mention these genes' roles in the RCDs within the context of cancer.
3. Organize the Information:
a) Compile the extracted quotes and their references for each gene symbol.
b) Ensure each quote is properly cited with a clickable URL for the original document, allowing easy verification and further reading.
4. Batch Processing:
a) Split the list of gene symbols into manageable batches (e.g., batches of 10 genes).
b) Process each batch sequentially, appending the results to an existing Excel file.
5. Generate R Code Snippet:
a) Create an R script to compile and save the results in Excel format with the following columns: Gene Symbol, Cancer Type, RCD Involvement (anti- or pro-necrosis), Quote, Document Title, Page, and Link.
b) Each batch analysis should generate a R code that appends the new output data in the previous generated Excel file.

**Reference**:

1. Galluzzi, L., et al., *Molecular mechanisms of cell death: recommendations of the Nomenclature Committee on Cell Death 2018.* Cell Death Differ, 2018. **25**(3): p. 486-541.
2. Cannon, M., et al., *DGIdb 5.0: rebuilding the drug-gene interaction database for precision medicine and drug discovery platforms.* Nucleic Acids Res, 2024. **52**(D1): p. D1227-D1235.

#### Supplementary figures

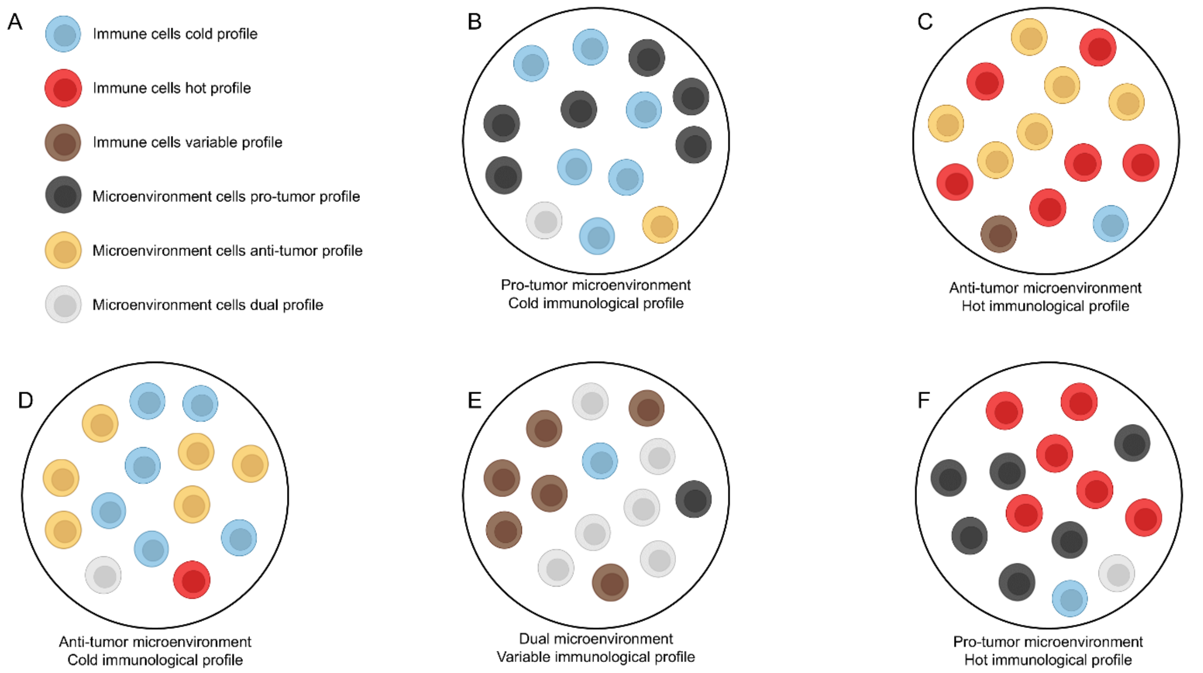


**Figure S1. Categorization of Tumor Microenvironment and Immune Phenotypes**
This cartoon representation supports our system for categorizing multi-omic signatures indicative of various tumor microenvironments and immune phenotypes across five scenarios. (A) Immune and microenvironmental cells are depicted using colors to represent their profiles. The immune cell profiles include "cold", "hot", and variable, while the microenvironment cell profiles include pro-tumor, anti-tumor, and dual. (B) Depicts a scenario with a pro-tumor microenvironment and a "cold" immunological profile, indicating greater resistance to therapy and a poorer prognosis. (C) Shows an anti-tumor microenvironment with a "hot" immunological profile, suggesting better success in immunotherapy and a favorable prognosis. (D) Represents an anti-tumor microenvironment with a "cold" immunological profile, indicating a less aggressive tumor profile but with a weak response to immunotherapy. (E) Illustrates a dual microenvironment with a variable immunological profile, where the tumor exhibits progression and treatment response features. (F) Shows a pro-tumor microenvironment with a "hot" immunological profile, indicating aggressive cancer but with a potential for a good response to immunotherapy.

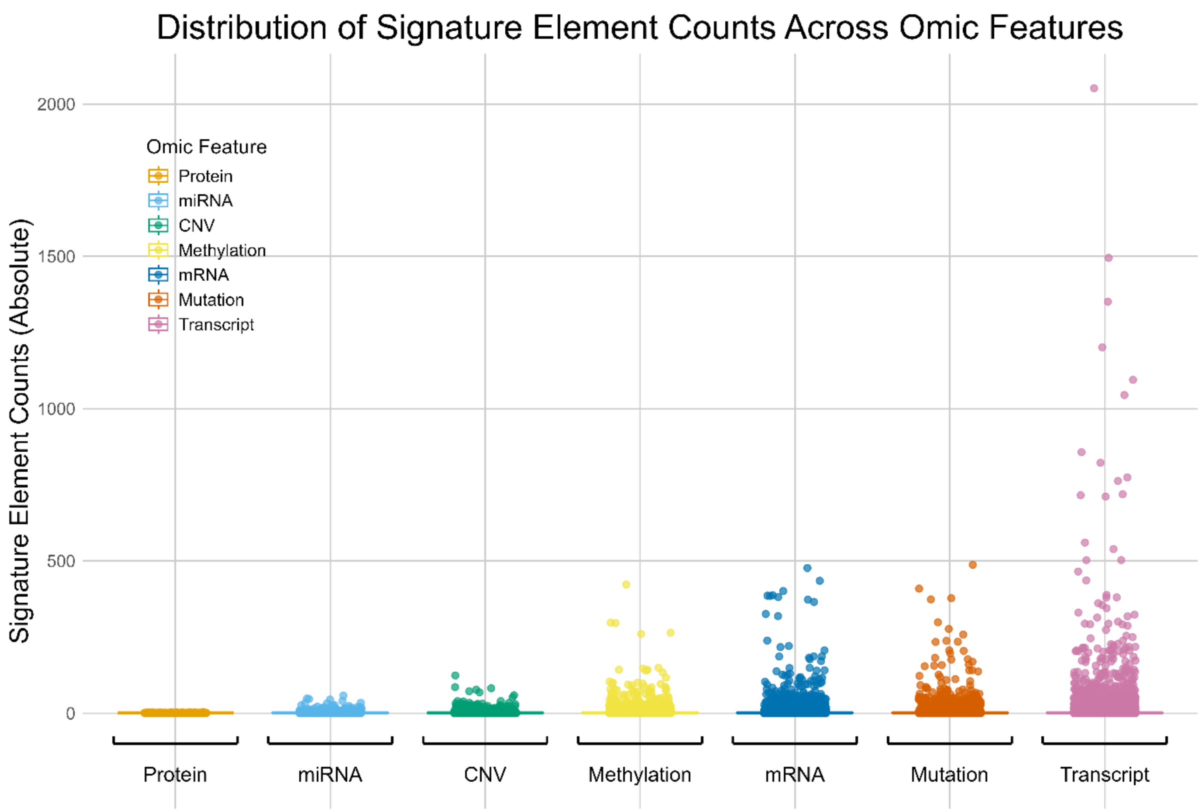


**Figure S2**. Distribution of signature member element counts across genomic features. The element counts are displayed in order of range, from smallest to largest, within each omic feature. Each feature is assigned a distinct color from the Okabe-Ito palette, ensuring clarity and accessibility for color-blind viewers. Individual data points are horizontally jittered to reduce overlap, clearly depicting the distribution and spread of counts within each genomic feature.

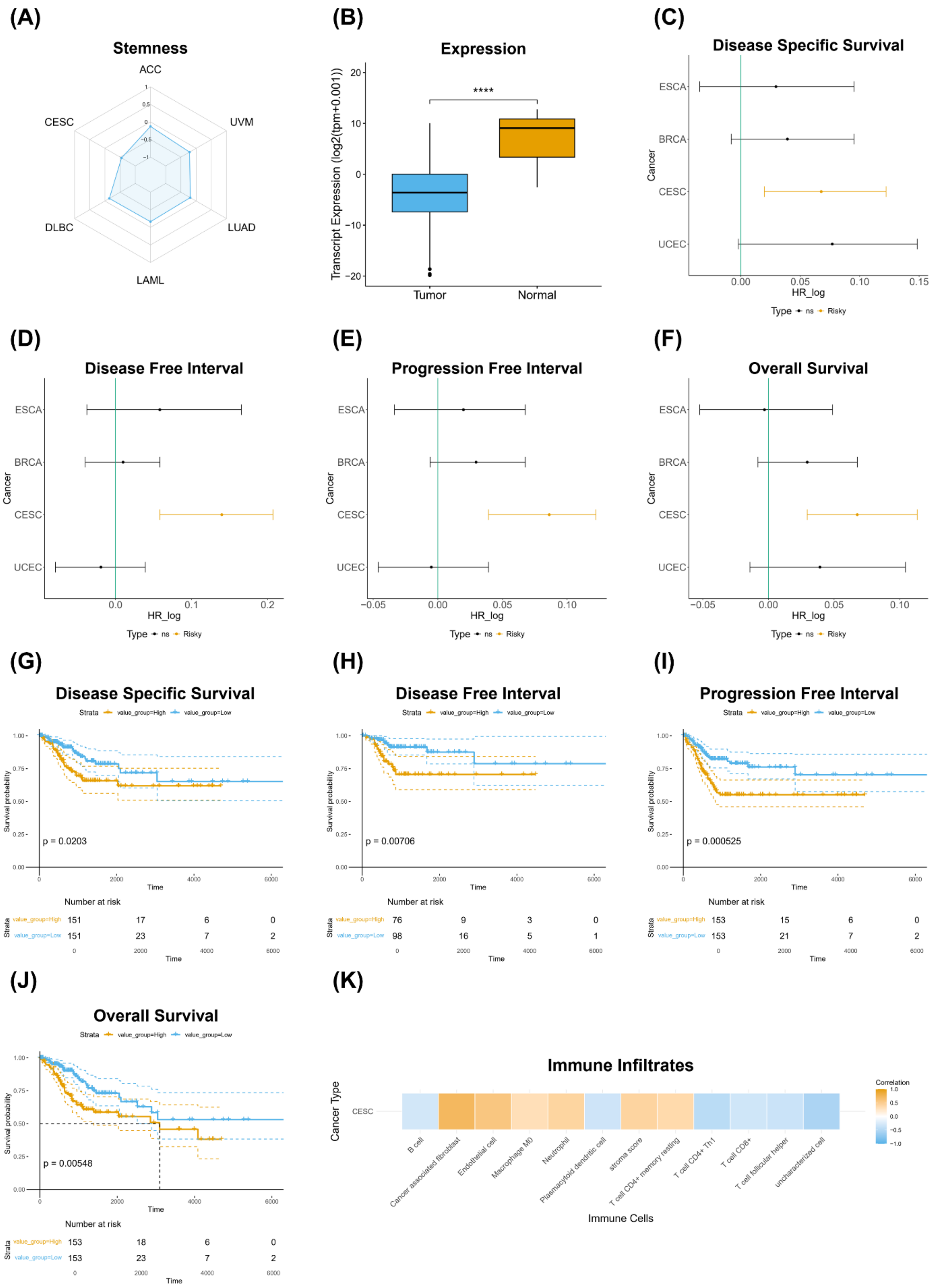


**Figure S3. Phenotypic associations and prognostic significance of the transcript signature CESC-215.5.3.N.2.44.44.1.1.3 in cervical squamous cell carcinoma and endocervical adenocarcinoma (CESC)**. Panel (A) shows a radar plot illustrating the negative correlation between transcript signature expression and TSM across multiple cancer types. Panel (B)

demonstrates significantly lower transcript expression for the transcripts in the signature in tumor samples compared to normal tissue (****$p < 0.0001$). Panels (C-F) present hazard ratio (HR) analyses evaluating the prognostic associations of the transcript signature with clinical outcomes across various cancer types, including (C) Disease-Specific Survival, (D) Disease-Free Interval, (E) Progression-Free Interval, and (F) Overall Survival, where a positive log HR indicates a risk effect of the transcript signature. Panels (G-J) display Kaplan-Meier survival curves for CESC patients stratified by high and low transcript signature expression, with significant survival outcomes for (G) Disease-Specific Survival ($p = 0.0203$), (H) Disease-Free Interval ($p = 0.00706$), (I) Progression-Free Interval ($p = 0.000525$), and (J) Overall Survival ($p = 0.00548$). Panel (K) illustrates the correlation between the transcript signature and immune cell infiltration in CESC, highlighting associations with various immune cell types.

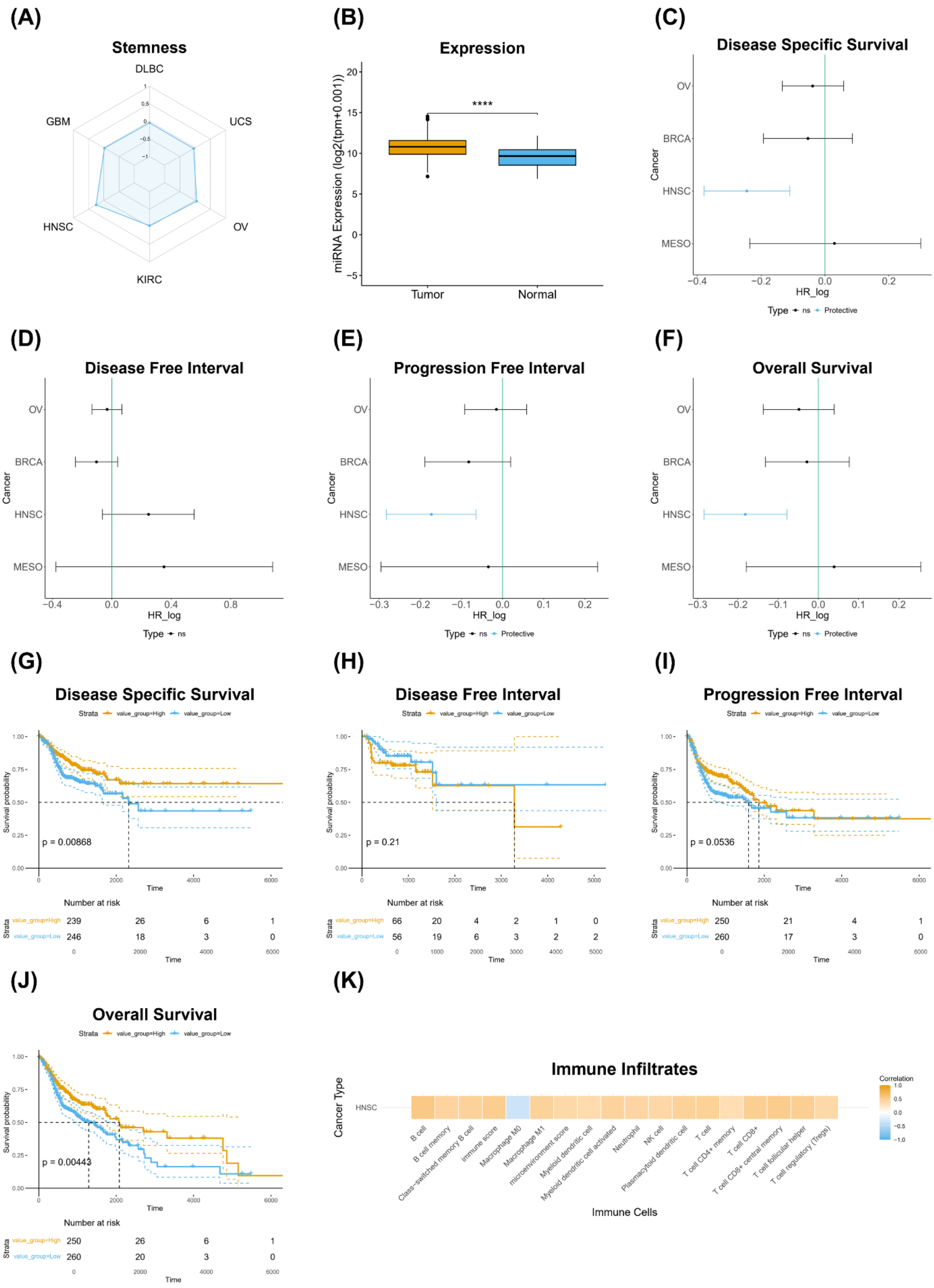


**Figure S4. Phenotypic associations and prognostic significance of the miRNA signature HNSC-1855.4.3.P.3.71.64.1.1.4 in head and neck squamous cell carcinoma (HNSC).** Panel (A) shows a radar plot illustrating the positive correlation between signature miRNA expression and TSM in various cancer types. Panel (B) demonstrates significantly higher miRNA expression for the miRNA signature in tumor samples compared to normal tissue (****$p < 0.0001$). Panels (C-F) present hazard ratio (HR) analyses evaluating the prognostic associations of the miRNA signature with clinical outcomes in multiple cancer types, including (C) Disease-specific survival, (D) Disease-free interval, (E) Progression-free interval, and (F) Overall survival, where a negative log HR indicates a protective effect of signature miRNA expression. Panels (G-J) display Kaplan-Meier survival curves for HNSC patients stratified by high and low miRNA signature expression, with significant survival outcomes for (G) Disease-Specific Survival ($p = 0.00868$), (H) Disease-Free Interval ($p = 0.21$), (I) Progression-Free Interval ($p = 0.0536$), and (J) Overall Survival ($p = 0.00443$). Panel (K) illustrates the correlation between signature miRNA expression and immune cell infiltration in HNSC, highlighting associations with various immune cell types.

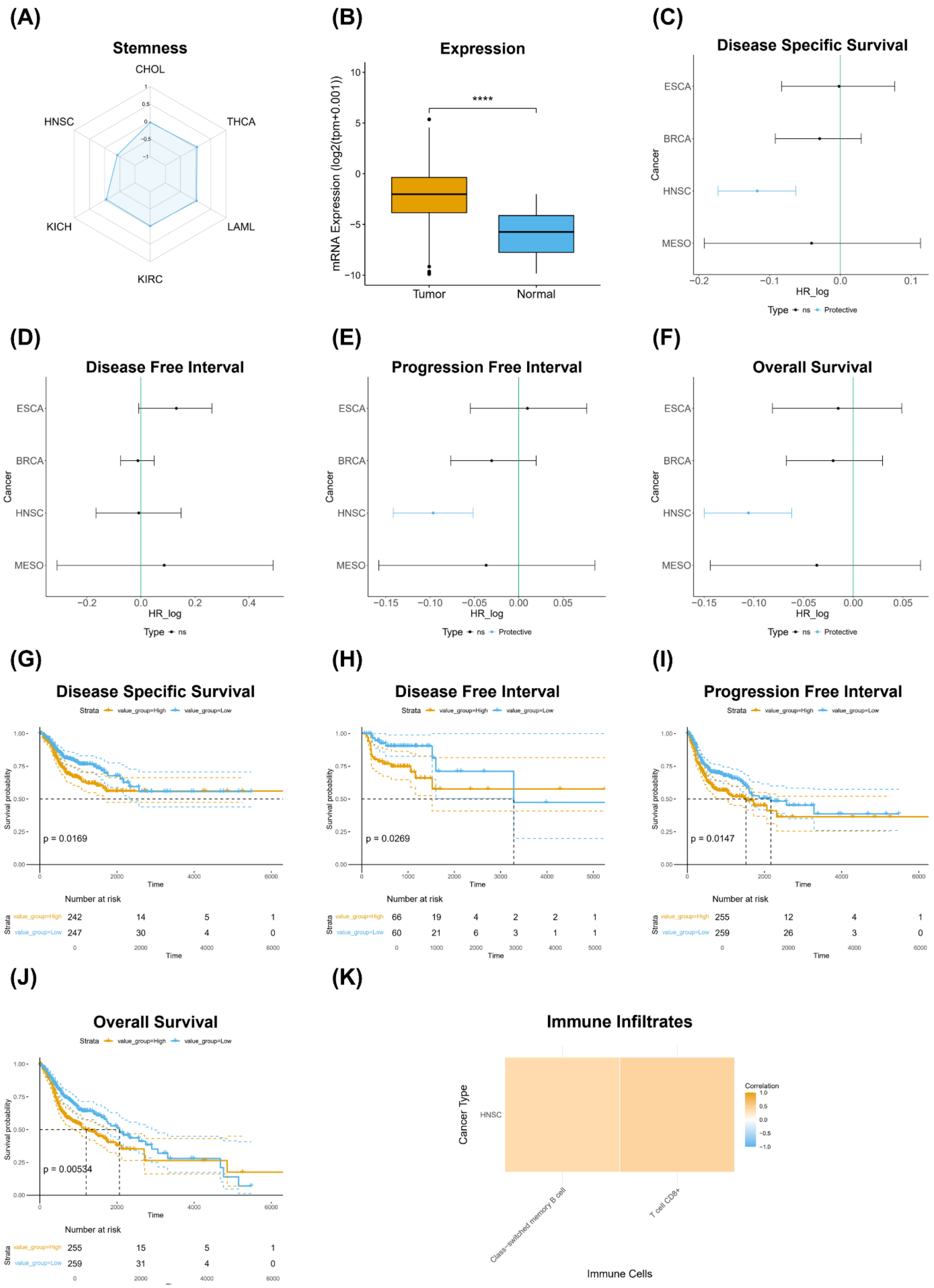


**Figure S5. Phenotypic associations and prognostic significance of the methylation-specific signature HNSC-156.7.3.N.3.71.55.1.1.1 in head and neck squamous cell carcinoma (HNSC).** Panel (A) shows a radar plot illustrating the negative correlation between methylation status of the signature and TSM across multiple cancer types. Panel (B) demonstrates significantly higher mRNA expression for the methylation signature in tumor samples compared to normal tissue (****p < 0.0001). Panels (C-F) present hazard ratio (HR) analyses evaluating the prognostic associations of the mRNA expression of the signature elements (CEBPE + SIRPG) with clinical outcomes in multiple cancer types, including (C) Disease-specific survival, (D) Disease-free interval, (E) Progression-free interval, and (F) Overall survival, where a negative log HR indicates a protective effect. Panels (G-J) display Kaplan-Meier survival curves for HNSC patients stratified by high and low methylation status of the signature, with significant survival outcomes for (G) Disease-Specific Survival (p = 0.0169), (H) Disease-Free Interval (p = 0.0269), (I) Progression-Free Interval (p = 0.0147), and (J) Overall Survival (p = 0.00534). Panel (K) illustrates the correlation between methylation signature mRNA expression and immune cell infiltration in HNSC, highlighting associations with various immune cell types.

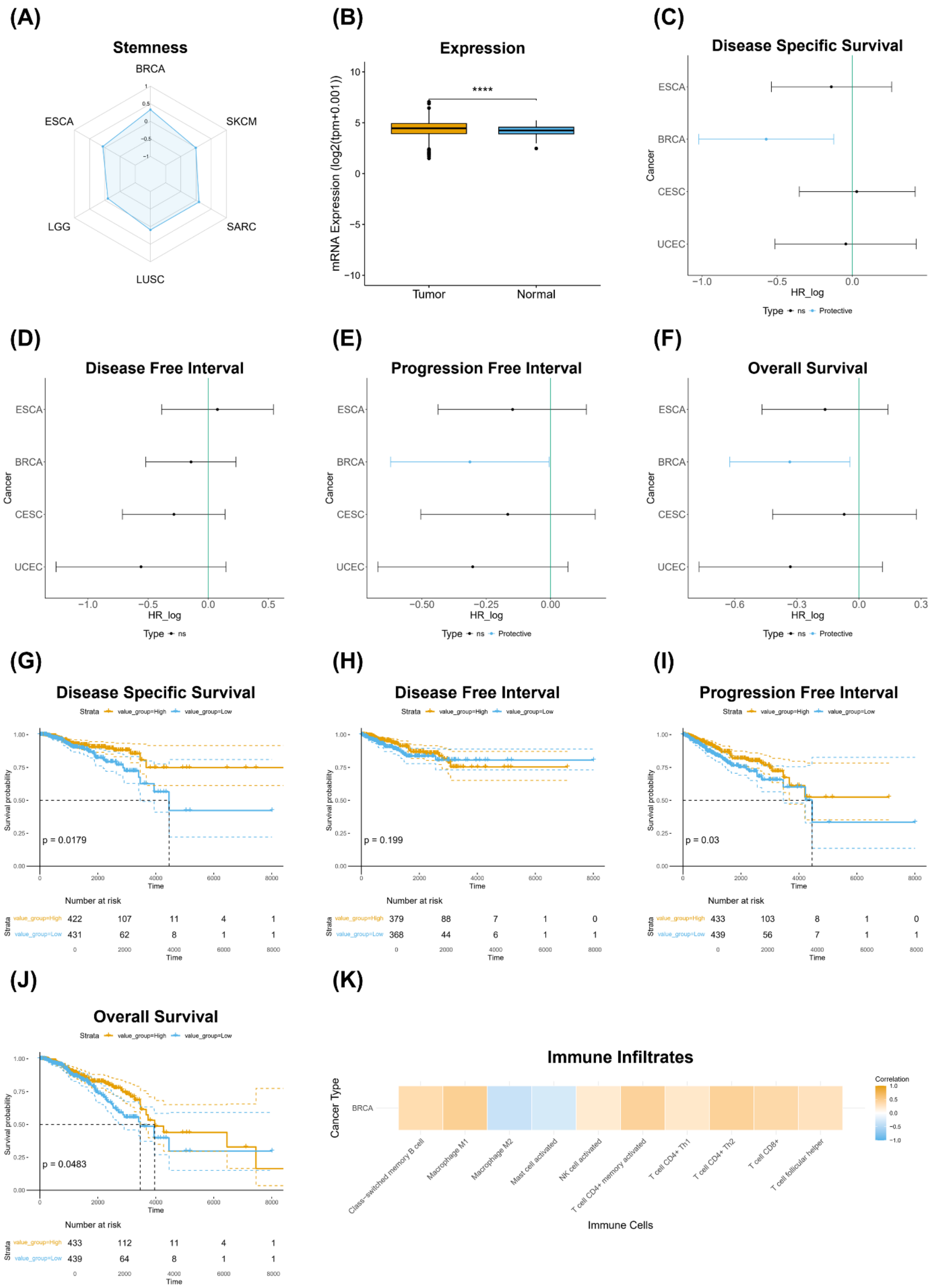


**Figure S6. Phenotypic associations and prognostic significance of the protein signature BRCA-1496.1.3.P.3.71.71.1.1.2 in breast cancer (BRCA)**. Panel (A) shows a radar plot illustrating the positive correlation between protein signature expression and TSM across multiple cancer types. Panel (B) demonstrates significantly higher mRNA expression for the gene encoding the signature element in tumor samples compared to normal tissue (****$p < 0.0001$). Panels (C-F) present hazard ratio (HR) analyses evaluating the prognostic associations of the protein signature with clinical outcomes across various cancer types, including (C) Disease-Specific Survival, (D) Disease-Free Interval, (E) Progression-Free Interval, and (F) Overall Survival, where a negative log HR shows a protective effect of the protein signature. Panels (G-J) display Kaplan-Meier survival curves for BRCA patients stratified by high and low protein signature expression, with significant survival outcomes for (G) Disease-Specific Survival ($p = 0.0179$), (H) Disease-Free Interval ($p = 0.199$), (I) Progression-Free Interval ($p = 0.03$), and (J) Overall Survival ($p = 0.0483$). Panel (K) illustrates the correlation between the protein signature and immune cell infiltration in BRCA, highlighting associations with various immune cell types.

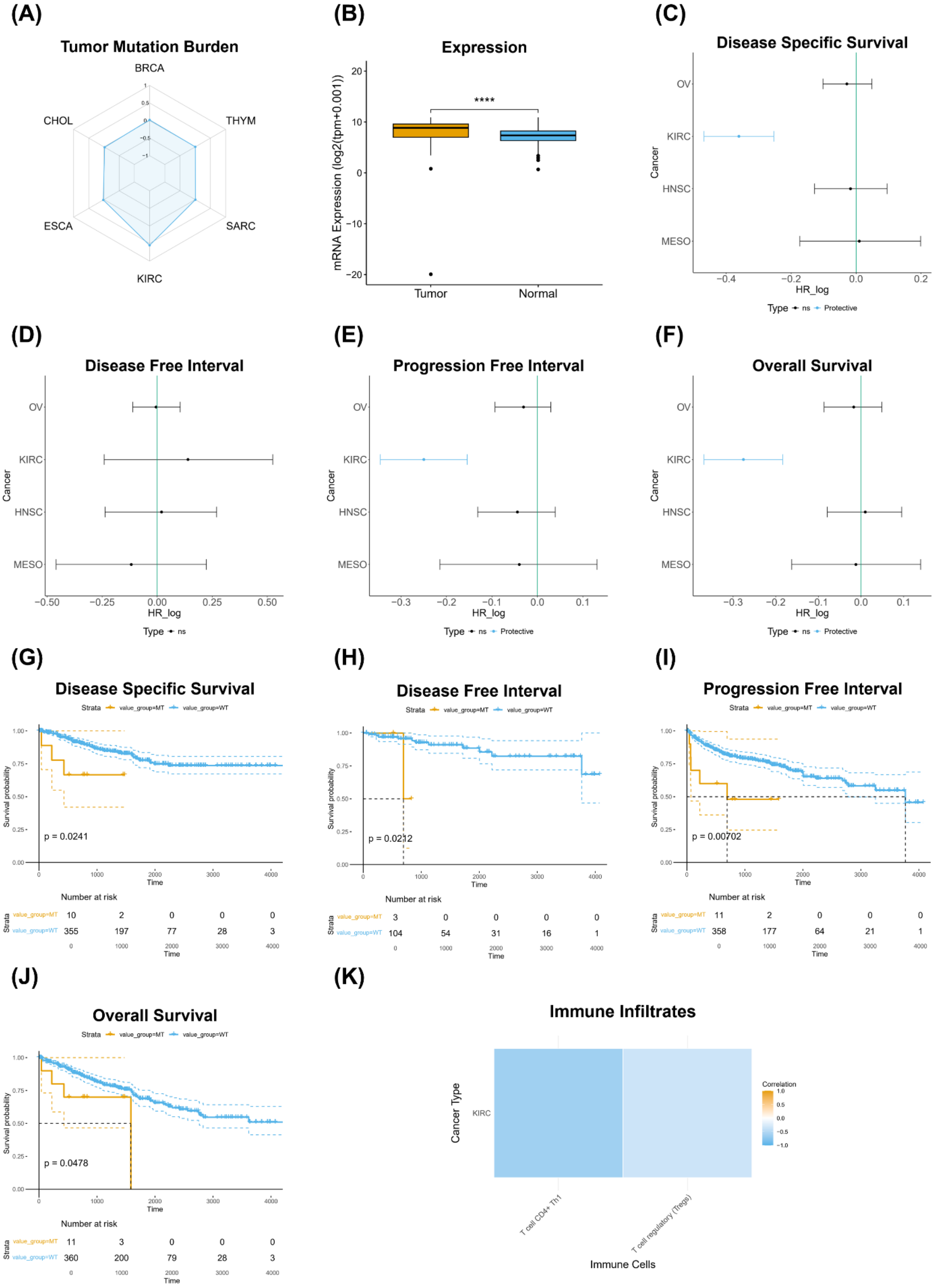


**Figure S7. Phenotypic associations and prognostic significance of the mutation signature KIRC-169.2.1.P.2.71.45.1.1.2 in kidney renal clear cell carcinoma (KIRC)**. Panel (A) shows a radar plot illustrating the positive correlation between mutation signature and TMB in various cancer types. Panel (B) demonstrates significantly higher mRNA expression for the mutation-specific signature in tumor samples compared to normal tissue (****$p < 0.0001$). Panels (C-F) present hazard ratio (HR) analyses evaluating the prognostic associations of mRNA expression of the mutation signature elements with clinical outcomes in multiple cancer types, including (C) Disease-specific survival, (D) Disease-free interval, (E) Progression-free interval, and (F) Overall survival, where a negative log HR indicates a protective effect. Panels (G-J) display Kaplan-Meier survival curves for patients with KIRC stratified by mutated and wildtype signature, with significant survival outcomes for (G) Disease-Specific Survival ($p = 0.0241$), (H) Disease-Free Interval ($p = 0.0212$), (I) Progression-Free Interval ($p = 0.00702$), and (J) Overall Survival ($p = 0.0478$). Panel (K) illustrates the correlation between mRNA expression of the mutation signature and immune cell infiltration in KIRC, highlighting associations with various immune cell types.

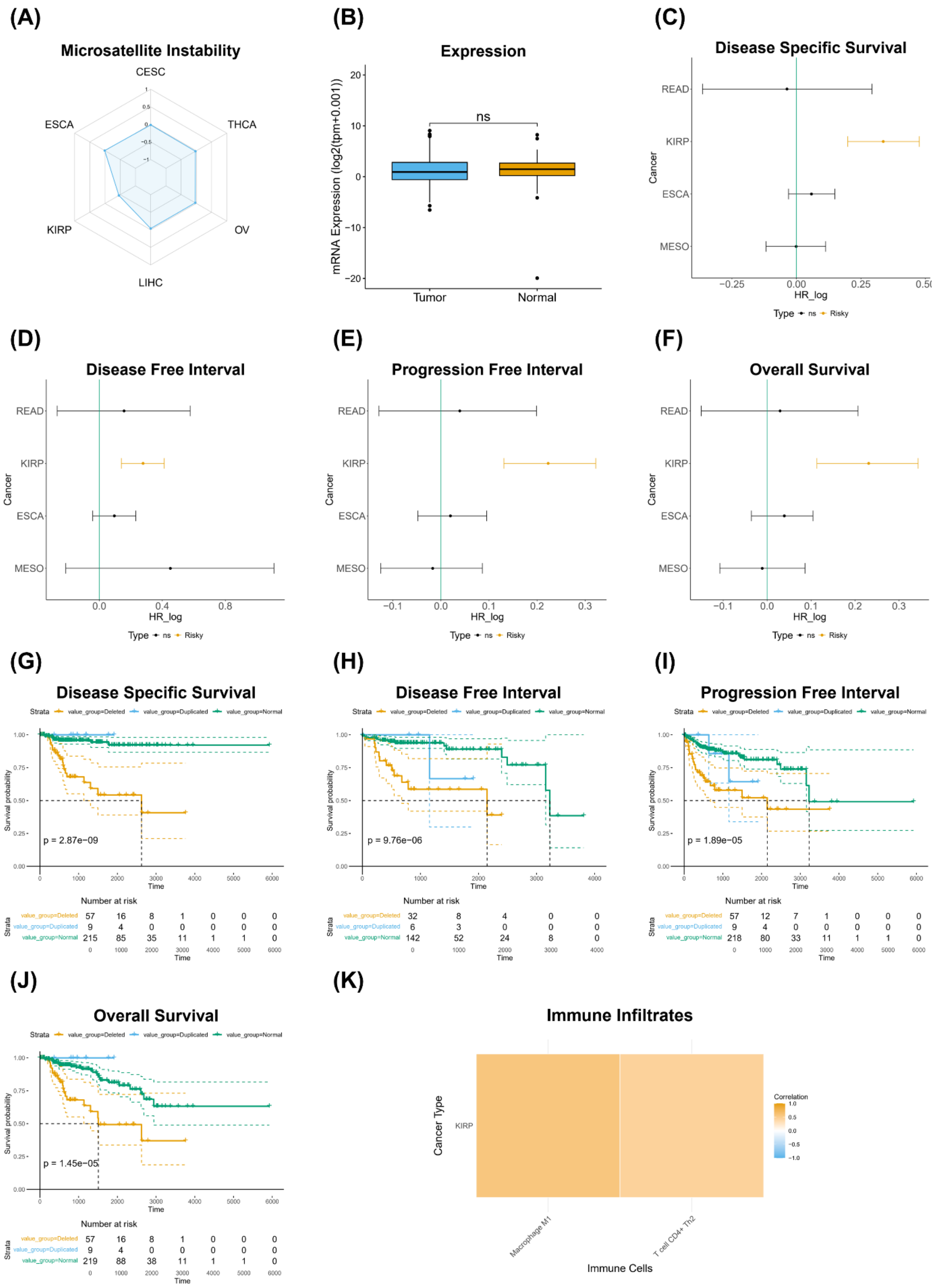


**Figure S8. Phenotypic associations and prognostic significance of the copy number variation signature KIRP-107.3.2.N.1.44.44.1.1.2 in kidney renal papillary cell carcinoma (KIRP)**. Panel (A) shows a radar plot illustrating the negative correlation between CNV signature and MSI in several cancer types. Panel (B) demonstrates unchanged mRNA expression of the CNV signature elements in tumor samples compared to normal tissue. Panels (C-F) present hazard ratio (HR) analyses evaluating the prognostic associations of the mRNA expression of CNV signature elements with clinical outcomes in multiple cancer types, including (C) Disease-specific survival, (D) Disease-free interval, (E) Progression-free interval, and (F) Overall survival, where a positive log HR shows a risk effect. Panels (G-J) display Kaplan-Meier survival curves for KIRP patients stratified by detected, duplicated, and normal CNV, with significant survival outcomes for (G) Disease-Specific Survival (p = 2.87e-09), (H) Disease-Free Interval (p = 9.76e-06), (I) Progression-Free Interval (p = 1.89e-05), and (J) Overall Survival (p = 1.45e-05). Panel (K) illustrates the correlation between the mRNA expression of the CNV signature elements and immune cell infiltration in KIRP patients, highlighting associations with various immune cell types.

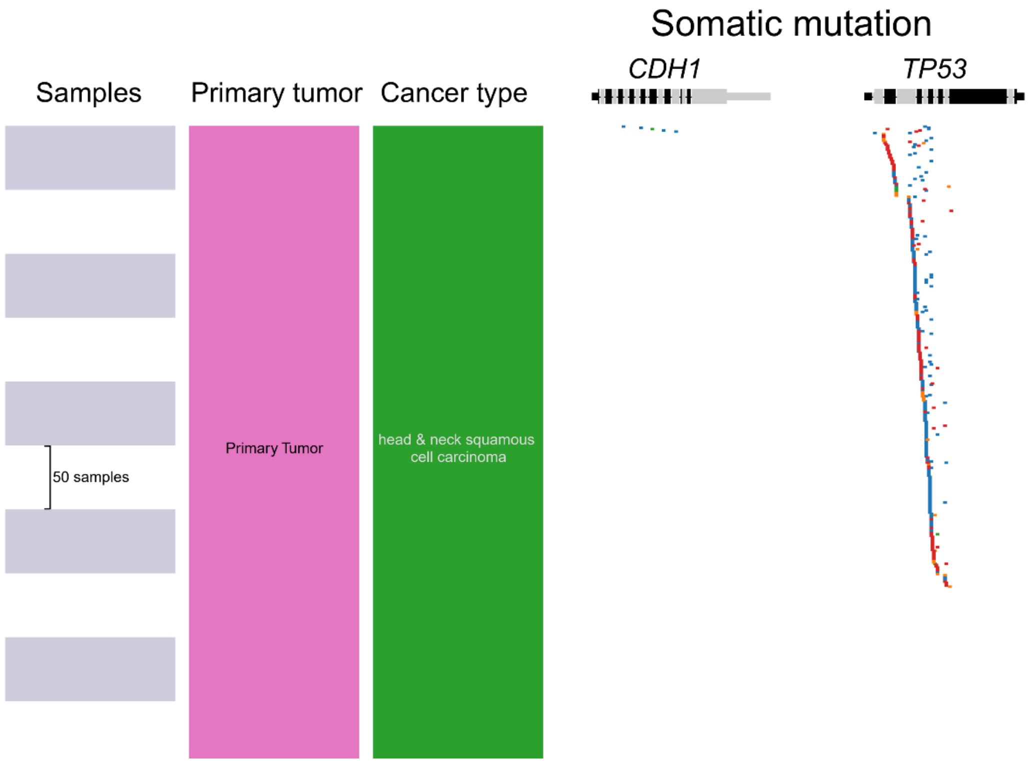


**Figure S9. Distribution of somatic mutations in the *CDH1*, and *TP53* genes across patients with HNSC (head and neck squamous cell carcinoma).** Dots indicate the locations of mutations across each gene's loci, with color coding by mutation type: deleterious (red), missense/in-frame (blue), splice-site (orange), silent (green), and intronic/RNA-associated (gray). Per-patient mutation details are available in Datasets S1T and S1U.

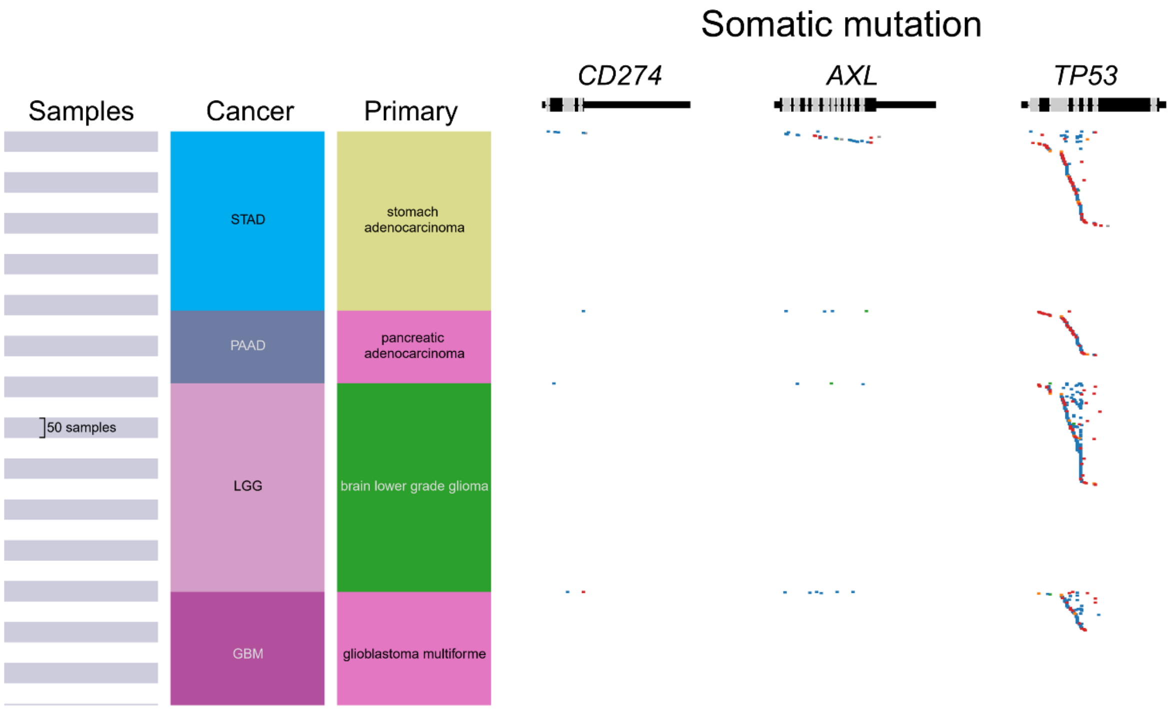


**Figure S10. Distribution of somatic mutations in the *CD274*, *AXL*, and *TP53* genes across patients with STAD (Stomach Adenocarcinoma), PAAD (Pancreatic Adenocarcinoma), GBM (Glioblastoma Multiforme), and LGG (Lower Grade Glioma).** Dots indicate the locations of mutations across each gene's loci, with color coding by mutation type: deleterious (red), missense/in-frame (blue), splice-site (orange), silent (green), and intronic/RNA-associated (gray). Per-patient mutation details are available in Datasets S1X, S1Y, S1Z and S1AA.

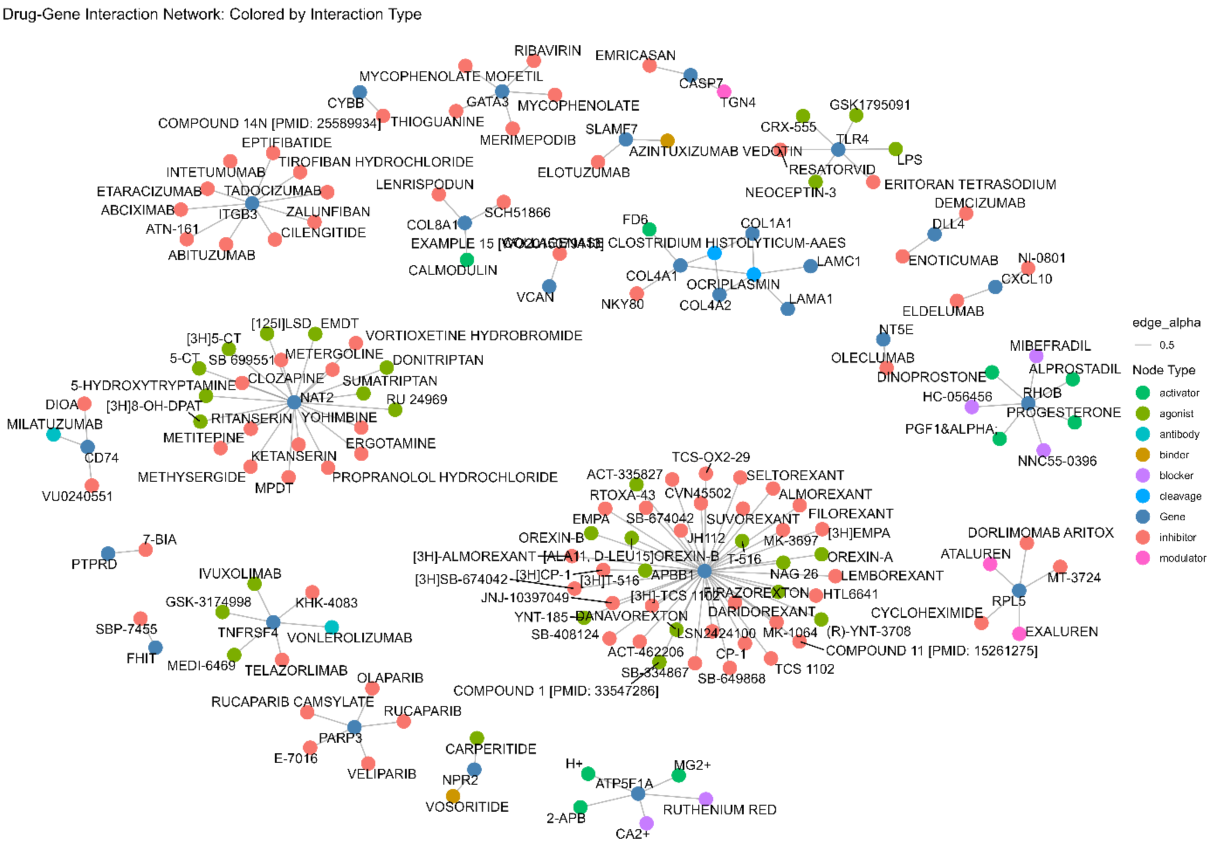


**Figure S11. Drug-Gene Interaction Network with Drug Effect Categorization.** This network visualization depicts drug-gene interactions, where nodes represent genes (steelblue) from top-ranked multi-omic RCD signatures and drugs (colored by interaction type). Edges show interactions between genes and drugs, sourced from the DGIdb database [2]. The drug nodes are color-coded according to their specific interaction effect type, as categorized in the dataset. The network layout follows a force-directed algorithm (Fruchterman-Reingold) to optimize node distribution and reveal interaction patterns. The legend on the right classifies drug interaction categories, highlighting the diversity of drug effects. The interactive plot can be accessed at URL https://github.com/CancerRCD/Supplementary-Material.

**Figure S12. Dynamic Sankey diagram illustrating the comprehensive negative correlations of *COL1A1* and *UMOD* gene isoforms with stemness across specific cancer signatures**
URL – https://github.com/CancerRCD/Supplementary-Material.

**Figure S13. Interactive Proportional Node Network of *MAPK10* transcripts.** This interactive network plot represents the associations of the gene *MAPK10* with various transcripts, phenotypes, and cancer types, highlighting key nodes by their connection frequency. Each node represents a unique entity within the network, color-coded by category: Gene symbol (core gene, in red), Transcript (blue), Phenotype (green), and Cancer type (yellow). The size of each node is proportional to its degree, or the number of connections it holds, with larger nodes indicating more frequent associations. Smooth edges illustrate connections between entities, enabling an intuitive exploration of *MAPK10*'s role in these relationships. Hovering over nodes reveals nearest connections, and the network is fully draggable for customized examination. This visualization was created using R and the visNetwork package, along with dplyr for data manipulation and htmlwidgets for saving the interactive plot as an HTML file.

URL - https://github.com/CancerRCD/Supplementary-Material.
